# Longitudinal Evaluation of Magnetic Resonance Spectroscopy Metabolites as Biomarkers in Huntington’s Disease

**DOI:** 10.1101/2021.10.26.21265516

**Authors:** Alexander J. Lowe, Filipe B. Rodrigues, Marzena Arridge, Eileanoir B. Johnson, Rachael I. Scahill, Lauren M. Byrne, Rosanna Tortelli, Amanda Heslegrave, Henrick Zetterberg, Edward J. Wild

## Abstract

Magnetic resonance spectroscopy (MRS) is a non-invasive method of exploring cerebral metabolism. In Huntington’s disease, altered MRS-determined concentrations of several metabolites have been described; however, findings are often discrepant and longitudinal studies of metabolite trajectory are lacking. MRS metabolites may represent a valuable source of biomarkers, thus their relationship with established biofluid and structural imaging markers of disease progression require further exploration to assess prognostic value and elucidate biochemical pathways associated with neurodegeneration. In a prospective single-site controlled cohort study with standardised collection of CSF, blood, phenotypic and imaging data, we used MRS to evaluate metabolic profiles in the putamen of 56 participants at baseline (15 healthy controls, 15 premanifest and 26 manifest gene expansion carriers) and at 2-year follow-up. Intergroup differences and associations with established measures were assessed cross-sectionally using generalized linear models and partial correlation, controlling for age and CAG repeat length. We report no significant groupwise differences in metabolite concentration but found several metabolites to be associated with measures of disease progression; however, only two relationships were replicated across both time points, with total Creatine (creatine + phosphocreatine) and myo-inositol displaying significant associations with reduced caudate volume. Although relationships were observed between MRS metabolites and biofluid measures, these were not consistent across time points. To further assess prognostic value of the metabolites, we examined whether baseline MRS values, or rate of change, predicted subsequent change in established measures of disease progression. Several associations were found but were inconsistent across known indicators of disease progression. Finally, longitudinal mixed effects models, controlling for age, revealed no significant change in metabolite concentration over time in gene expansion carriers. Altogether, our findings show some interesting cross-sectional associations between select metabolites, namely total creatine and myo-inositol, and markers of disease progression, potentially highlighting the proposed roles of neuroinflammation and metabolic dysfunction in disease pathogenesis. However, the absence of group differences, inconsistency between baseline and follow-up, and lack of clear longitudinal change over two years suggests that MRS metabolites have limited potential as biomarkers in Huntington’s disease.

## Introduction

Huntington’s disease is a neurodegenerative disease characterised by progressive motor, psychiatric and cognitive dysfunction.^1^ Invariably fatal, Huntington’s disease is caused by an autosomal dominant mutation in the *HTT* gene, producing a CAG repeat expansion in the ubiquitously expressed huntingtin protein (HTT).^2^ This mutated pathogenic product (mHTT) causes a wide array of toxicities and disruption of downstream pathways, resulting in neuronal death.^3^ With genetic testing, the development of Huntington’s disease can be accurately predicted; however, there remains a need to discover clinically relevant biomarkers with the ability to detect and quantify pathogenic change, pharmacological target engagement and treatment response.^4^ Due to its non-invasive nature, accessibility and the potential to standardise parameters across multiple sites, neuroimaging is a valuable source of information about progression and prognosis^4^ and has been utilised in Huntington’s disease to explore cross-sectional and longitudinal changes in brain structure, metabolism and activation patterns.^5–12^

Magnetic resonance spectroscopy (MRS) is a non-invasive method of exploring cerebral metabolism, and represents an interesting avenue in biomarker research as neurometabolic alterations may occur prior to the emergence of structural and functional change.^13,14^ The number of quantifiable metabolites depends on several factors including pulse sequence, spectral resolution and signal-to-noise ratio (SNR),^15^ all of which can be influenced by the magnetic field strength, with higher strengths providing increased sensitivity and spectral resolution.^16,17^ In the context of neurodegenerative disease, tNAA (N-acetylaspartate + N-acetylaspartate-glutamate), tCho (phosphocholine + glycophosphocholine), tCre (creatine + phosphocreatine) and myo-inositol (MI) are considered respective biomarkers for neuro-axonal viability and mitochondrial dysfunction,^18,19^ cellular proliferation and neuronal membrane turnover ^20,21^, brain energy metabolism and gliosis,^22^ and astrocytic density.^23^ Due to its relative stability in pathological conditions, creatine (Cr) is often used as an internal reference^24^; however, it is affected in Huntington’s disease, so MRS metabolites may be normalised to unsuppressed water signal, allowing the accurate identification of biochemical change in the brain.^25^

In Huntington’s disease, altered concentrations of several MRS metabolites have been described in both premanifest and manifest patients across multiple brain regions ^26–33^; however, other studies have reported no significant differences in metabolite concentrations when comparing patient cohorts to controls (Table 1).^34,35^ These discrepant findings are likely due, in part, to sample size variations, patient heterogeneity and differences in spatial/spectral resolution. Recent work leveraging 7-tesla MRI^35^ found lower metabolite levels to correspond to poorer clinical, cognitive and behavioural scores, similar to work leveraging the TRACK-HD cohort in which tNAA displayed a significant negative correlation with disease burden score (DBS) across pre- and early manifest patients, further demonstrating its role as a marker of clinical decline.^25^ Longitudinal analyses have produced mixed results thus far, with reduced tNAA and Cho in the putamen, and Cr and MI in the caudate, reported,^36^ whereas other have reported no longitudinal change in metabolite concentration.^36,37^ Importantly, the latter two studies normalised metabolite values to unsuppressed water signal, whilst also benefitting from high SNR and large sample sizes; however, the role of MRS metabolites as prognostic biomarkers remains debatable and warrants further study.

**Table 1.**
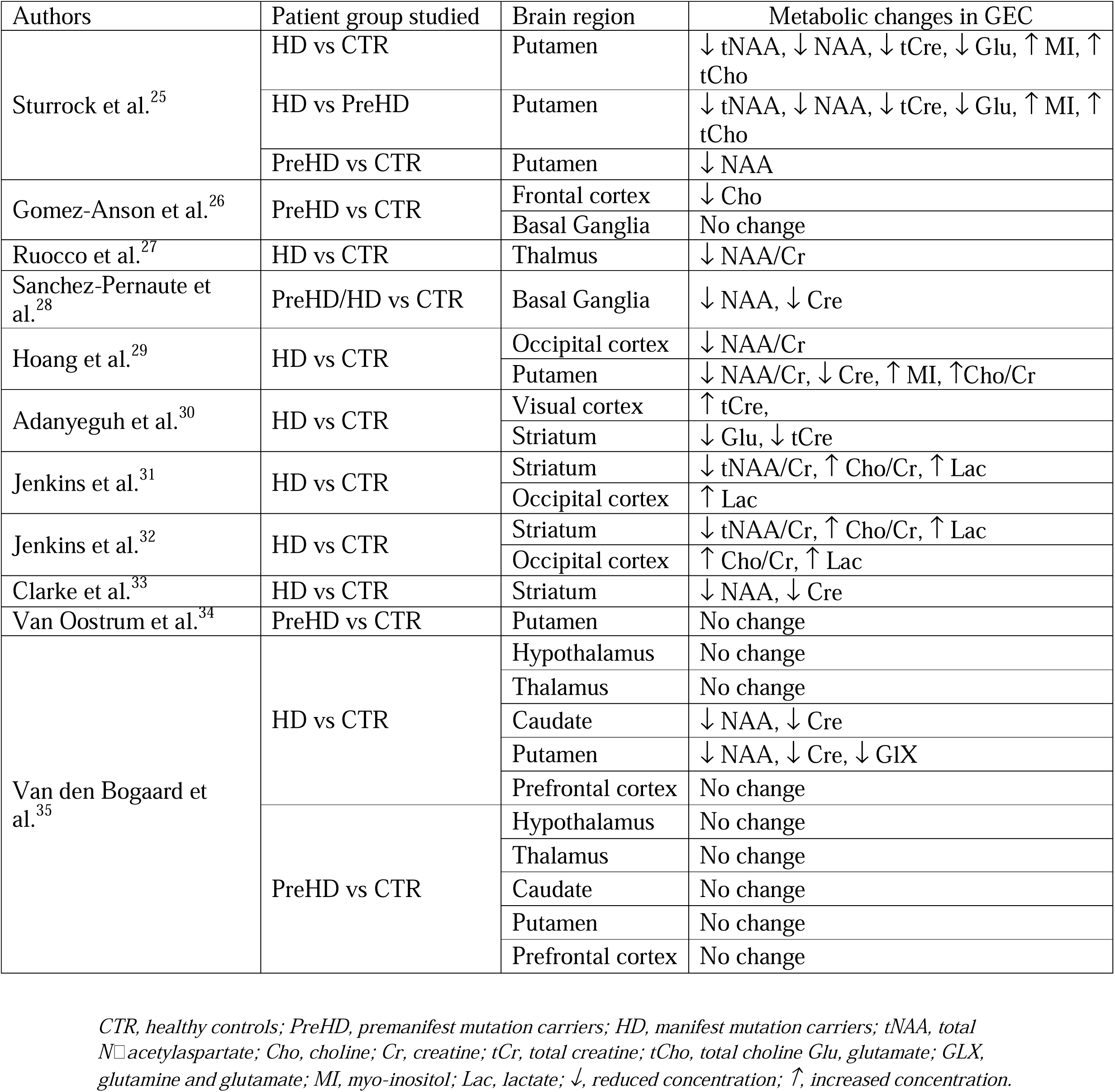
Summary of MRS Studies in HD.

| Authors | Patient group studied | Brain region | Metabolic changes in GEC |
| --- | --- | --- | --- |
| Sturrock et al.<sup>25</sup> | HD vs CTR | Putamen | ↓ tNAA, ↓ NAA, ↓ tCre, ↓ Glu, ↑ MI, ↑ tCho |
|  | HD vs PreHD | Putamen | ↓ tNAA, ↓ NAA, ↓ tCre, ↓ Glu, ↑ MI, ↑ tCho |
|  | PreHD vs CTR | Putamen | ↓ NAA |
| Gomez-Anson et al.<sup>26</sup> | PreHD vs CTR | Frontal cortex | ↓ Cho |
|  |  | Basal Ganglia | No change |
| Ruocco et al.<sup>27</sup> | HD vs CTR | Thalamus | ↓ NAA/Cr |
| Sanchez-Pernaute et al.<sup>28</sup> | PreHD/HD vs CTR | Basal Ganglia | ↓ NAA, ↓ Cre |
| Hoang et al.<sup>29</sup> | HD vs CTR | Occipital cortex | ↓ NAA/Cr |
|  |  | Putamen | ↓ NAA/Cr, ↓ Cre, ↑ MI, ↑ Cho/Cr |
| Adanyeguh et al.<sup>30</sup> | HD vs CTR | Visual cortex | ↑ tCre, |
|  |  | Striatum | ↓ Glu, ↓ tCre |
| Jenkins et al.<sup>31</sup> | HD vs CTR | Striatum | ↓ tNAA/Cr, ↑ Cho/Cr, ↑ Lac |
|  |  | Occipital cortex | ↑ Lac |
| Jenkins et al.<sup>32</sup> | HD vs CTR | Striatum | ↓ tNAA/Cr, ↑ Cho/Cr, ↑ Lac |
|  |  | Occipital cortex | ↑ Cho/Cr, ↑ Lac |
| Clarke et al.<sup>33</sup> | HD vs CTR | Striatum | ↓ NAA, ↓ Cre |
| Van Oostrum et al.<sup>34</sup> | PreHD vs CTR | Putamen | No change |
| Van den Bogaard et al.<sup>35</sup> | HD vs CTR | Hypothalamus | No change |
|  |  | Thalamus | No change |
|  |  | Caudate | ↓ NAA, ↓ Cre |
|  |  | Putamen | ↓ NAA, ↓ Cre, ↓ GIX |
|  |  | Prefrontal cortex | No change |
|  | PreHD vs CTR | Hypothalamus | No change |
|  |  | Thalamus | No change |
|  |  | Caudate | No change |
|  |  | Putamen | No change |
|  |  | Prefrontal cortex | No change |

CTR, healthy controls; PreHD, premanifest mutation carriers; HD, manifest mutation carriers; tNAA, total N-acetylaspartate; Cho, choline; Cr, creatine; tCr, total creatine; tCho, total choline; Glu, glutamate; GLX, glutamine and glutamate; MI, myo-inositol; Lac, lactate; ↓, reduced concentration; ↑, increased concentration.

The relationship between biofluid markers and MRS metabolites requires further exploration, as combining direct and non-invasive quantifications of biochemical alterations could improve the value of both biomarker modalities. The concentration of neurofilament light chain (NfL), measured in CSF and blood, represents axonal damage and is a prognostic biomarker of neurodegeneration.^38–40^ Its relationship with MRS metabolites, particularly tNAA, MI and tCre, warrants additional study but has not previously been examined in Huntington’s disease. In patients with HIV, elevated levels of CSF NfL have been shown to correlate with decreased NAA/Cre in multiple brain regions, indicating compromised neuronal health and stability.^41^ In multiple sclerosis patients, the same inverse relationship has been observed between serum NfL and NAA/Cre at baseline, yet is not present at 12 and 36 months following hematopoietic stem cell transplantation.^42^ Given the elevated concentration of NfL in Huntington’s disease, we hypothesised that an inverse relationship with tNAA and tCre would be present in the putamen of Huntington’s disease patients. Furthermore, mHTT can be accurately quantified in CSF following its release from damaged neurons^43,44^ and displays strong associations with CSF NfL.^39,40,44^ As such, we would expect to observe the same relationships with CSF mHTT. In Alzheimer’s disease, reduced NAA/Cre and increased MI/Cre have been associated with increased p- and t-tau, and decreased CSF amyloid-beta (Aβ42), across several brain regions.^45–47^ The association between MI and tau, another established marker of neurodegeneration,^48^ is thought to be driven by activation of MI-rich astrocytes and microglia.^46^ Additionally, given that neuroinflammation represents a key pathogenic component,^49^ and source of CSF biomarkers,^50^ in Huntington’s disease, we expect to observe positive correlations between MI and all biofluid markers, reflecting the contribution of excessive neuroinflammatory response on disease pathogenesis.

We employed MRS to conduct a cross-sectional and longitudinal neurochemical analysis in the putamen of gene expansion carriers and healthy controls. Using the HD-CSF cohort,^39,40^ a large prospective sample of gene expansion carriers and matched controls with CSF and blood plasma collection and 3T MRI acquisition, we examined the biomarker potential of key MRS metabolites and explored their relationship with several established biofluid markers. We tested the hypothesis that significant metabolic alterations would be observed in the putamen of Huntington’s disease patients, specifically tNAA, MI and tCre, and would correlate with measures of clinical progression and established prognostic biomarkers quantified in CSF and blood.

## Materials and Methods

### Participants

HD-CSF was a prospective single-site study with standardised longitudinal collection of CSF, blood, and phenotypic data (online protocol: 10.5522/04/11828448.v1) from manifest patients (HD), premanifest (PreHD) patients and healthy controls (CTR). Eighty participants were recruited (20 CTR, 20 PreHD and 40 HD) based on a priori sample size calculations to detect cross-sectional and longitudinal differences in CSF mHTT between healthy controls and gene expansion carriers.^44^ 3T MRI scans were optional for HD-CSF participants. The present study used MRS data obtained from 59 participants at baseline and 48 at longitudinal follow-up after 24 months. Manifest Huntington’s disease was defined as Unified Huntington’s Disease Rating Scale (UHDRS)^51^ diagnostic confidence level (DCL) = 4 and CAG repeat length > 36. PreHD had CAG repeat length > 40 and DCL < 4. Healthy controls were contemporaneously recruited, drawn from a population with a similar age to patients, and clinically well, so the risk of incidental neurological diseases was very low. Consent, inclusion and exclusion criteria, clinical assessment, CSF collection and storage were as previously described.^39,52^ Baseline and longitudinal 24-month follow-up samples from HD-CSF have been used for this study.

### Ethical Approval

Ethical approval was given by the London Camberwell St Giles Research Ethics Committee (15/LO/1917), with all participants providing written informed consent prior to enrolment. This study was performed in accordance with the principles of the Declaration of Helsinki, and the International Conference on Harmonization Good Clinical Practice standards.

### Clinical Assessments

Relevant aspects of clinical phenotype were quantified using the UHDRS.^51^ A composite UHDRS (cUHDRS) score was generated for each subject to provide a single measure of motor, cognitive and global functioning decline. This composite score is computed using the following formula (Total Functional Capacity, TFC; Total Motor Score, TMS; Symbol Digit Modality Test, SDMT; Stroop Word Reading, SWR):

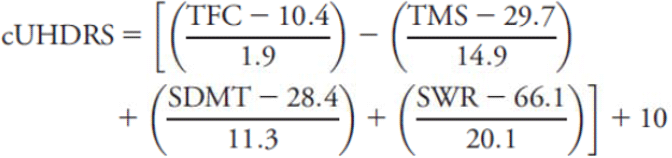

cUHDRS score has been found to display the strongest relationship to Huntington’s disease brain pathology and enhanced sensitivity to clinical change in early manifest disease.^53^ Disease burden score (DBS) was calculated for each gene expansion carrier using the formula [CAG repeat length – 35.5] × age.^54^ DBS estimates cumulative pathology exposure as a function of CAG repeat length and the time exposed to the effects of the expansion and has been shown to predict several features of disease progression including striatal pathology.^11,54^

### Volumetric MRI Acquisition

T1-weighted MRI data were acquired on a 3T Siemens Prisma scanner using a protocol optimized for this study. Images were acquired using a 3D magnetization-prepared 180 degrees radio-frequency pulses and rapid gradient-echo (MPRAGE) sequence with a repetition time (TR) =2000 ms and echo time (TE)=2.05 ms. The protocol had an inversion time of 850 ms, flip angle of 8 degrees, matrix size 256 × 240 mm. 256 coronal partitions were collected to cover the entire brain with a slice thickness of 1·0 mm. Parallel imaging acceleration (GeneRalized Autocalibrating Partial Parallel Acquisition [GRAPPA], acceleration factor [R]=2) was used and 3D distortion correction was applied to all images. Volumetric measures (whole brain, grey matter, white matter and caudate volume) were computed using the previously described methodology^39,40,55^ and adjusted for total intracranial volume (TIV).

### Magnetic Resonance Spectroscopy and LCModel Quantification

All scans were performed using 3T Siemen’s scanner (Prisma VE11C) with 64 channels RF head coil.

For spectroscopy, a single voxel spin echo-based Siemens sequence (svs) was used with the following parameters: echo time TE=30ms; repetition time TR= 2000ms; vector size (number of points in the time domain) = 2048; spectral width = 2400 Hz. Spectra was acquired from a rectangular volume of interest (VOI): 35×10×15 mm3 (Fig. 1). Adjustments included: transmitter gain, receiver gain, shimming (3D Gradient Echo followed by manual adjustments to achieve less than 14 Hz water linewidth), and water suppression. Water suppressed spectrum was acquired with 160 averages with a reference scan (unsuppressed spectrum 4 averages).

**Fig. 1.**
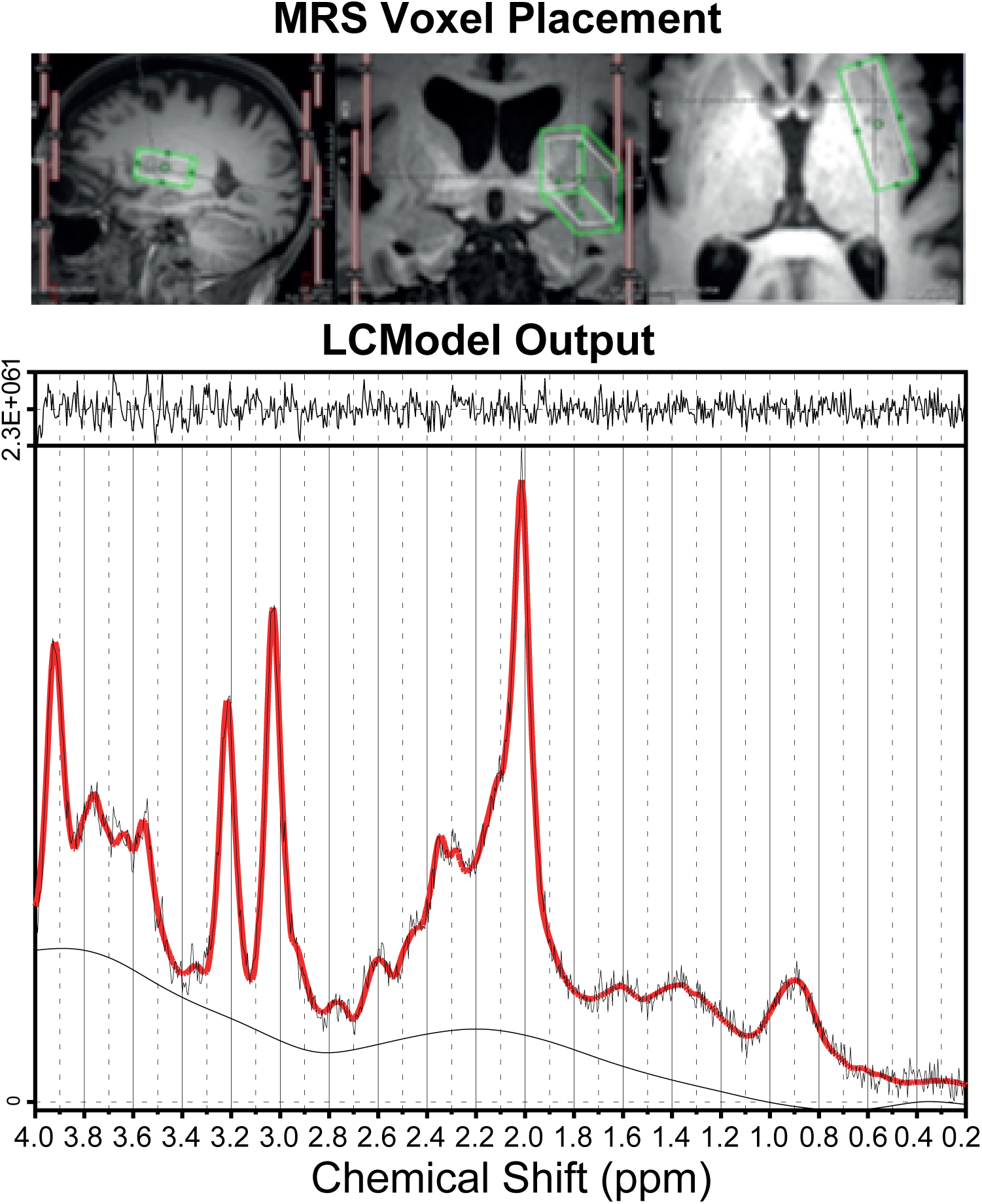
Voxel placement and LCModel output: Example voxel placement in right putamen displayed in all three planes (Top). LCModel output generated from Huntington’s disease mutation carrier (Premanifest) with black and red lines representing the raw spectrum and model fit overlaid on raw data, respectively (Bottom). Output peaks represent specific metabolite concentrations. ppm, parts per million.

LCModel (v6.3-1L) spectra of 18 metabolites were included in the basis data set together with model spectra for macromolecules and lipids. Metabolite levels were estimated using internal water as a reference. The LCModel produces standard deviations (%SD) for each metabolite as a measurement of reliability, with SDs below 20% considered reliable. Only GABA had a mean %SD of >20% at both baseline and follow-up. As a quality control measure, we removed any subject with a %SD >= 100 from the analyses. This was applied to all metabolites, resulting in the removal of subjects from the GABA cohort only (8 from baseline, and 6 from the follow-up cohort; Supplementary Table 1). Spectral quality was assessed individually for all data. Spectra with SNR < 6 were deemed unacceptable for further analysis due to the presence of artefacts, and/or inaccurate fitting of spectra, and excluded.

### Biofluid Collection and Processing

CSF and matched plasma were obtained as previously described.^39,40^ All collections were standardised for time of day after overnight fasting and processed within 30 minutes of collection using standardised equipment. Blood was collected within 10 minutes of CSF and processed to plasma. Biosamples were frozen and stored at -80°C.

### Analyte Quantification

Analytes were quantified as previously described.^39,40^

CSF and plasma NfL were quantified in duplicate using the Neurology 4-Plex B assay on the Simoa HD-1 Analyzer (Quanterix). The limit of detection (LoD) was 0.105 pg/ml, and lower limit of quantification (LLoQ) was 0.500 pg/ml. NfL was above the LLoQ in all samples. The intra-assay coefficients of variation (CV) (calculated as the mean of the CVs for each sample’s duplicate measurements) for CSF NfL and plasma NfL were 5.0% and 3.7%, respectively. The inter-assay CVs (calculated as the mean of the CVs for analogous spiked positive controls provided by the manufacturer and used in each well plate) for CSF NfL and plasma NfL were 2.7% and 8.4%, respectively. CSF mHTT was quantified in triplicate using the 2B7-MW1 immunoassay (SMC Erenna platform, Merck). The LoD was 8 fM, LLoQ was 25 fM and the intra-assay CV was 14.1%. CSF total tau was quantified using the INNOTEST enzyme-linked immunosorbent assay according to the manufacturer’s instructions (Fujirebio, Ghent, Belgium).

All biofluid measures, except CSF mHTT, were log-transformed to meet model assumptions.

### Statistical Analysis

Statistical analysis was performed with Stata IC 15 software (StataCorp, TX, USA). The distributions of all metabolite concentrations were visually assessed using kernel density estimate plots and Q-Q plots. Data transformations were not required to meet model assumptions (Supplementary Fig. 1). Differences in demographic, clinical, cognitive, imaging and biofluid measures were examined using Chi squared tests and generalised linear models (GLMs). Models were not adjusted for age, or CAG length, at this stage.

To reduce the risk of type 1 error, we preselected tNAA, tCre, tCho and MI as primary outcome measures based on the published MRS literature in Huntington’s disease (see introduction). Glutathione (GSH), GABA and GLX were designated as secondary outcome measures. Age and gender were considered potentially confounding variables, thus their relationship with metabolite concentration was examined within controls using Pearson’s correlation and independent samples t-tests. Using linear regression, all metabolites were adjusted for the partial volume effect of cerebrospinal fluid (CSF PVE), with the residuals being used in the subsequent analysis.

To investigate intergroup differences, we constructed two GLMs: one controlling for age; the other for and age and including CAG. The latter was used when comparing premanifest to manifest patients, and the former when comparing healthy controls to premanifest individuals. Additionally, we applied an inverse weighting to the %SD values of each metabolite to compensate for any variations in LCModel output quality. By including both age and CAG as covariates, accurate assessments of associations can be made, independent of known predictors. Due to the exploratory nature of the study, tests were not adjusted for multiple comparisons.

Associations between metabolites and clinical, cognitive, imaging measures, and established biofluid markers were explored cross sectionally using Pearson’s partial correlation controlling for age, and age and CAG, in gene expansion carriers only. DBS is a product of age and CAG, as such, we did not adjust DBS for the effects of these variables. In keeping with our regression analysis, we removed any subject with a %SD >= 100 and applied inverse weighting to the %SD values of the remaining subjects. This process was applied to each metabolite individually. Additionally, we performed unweighted, bootstrapped (1000 repetitions) partial correlations in which bias-corrected and accelerated 95% confidence intervals (95% CI) were calculated for correlation coefficients. Metabolites were deemed to have prognostic potential if a significant relationship was observed across all four correlation models (Inverse weighting controlling for age, and age and CAG; Bootstrapped controlling for age, and age and CAG). Although stringent, we chose this method to allow identification of MRS metabolites that demonstrate the strongest biomarker potential. No adjustments were made for multiplicity.

The cross-sectional statistical analyses outlined above was also applied to the follow-up dataset. We reasoned that the 2 years’ disease progression in all gene expansion carriers might outweigh the loss of power from participant dropout.

Annualised rate of change for each MRS metabolite and clinical, cognitive, and imaging measures were computed by subtracting baseline from follow-up values and dividing by the time between visits in years. Intergroup differences and correlations were examined using the methods outlined above. Only those subjects with data at both baseline and follow-up were included in this analysis.

To study longitudinal trajectories of the metabolites, we used mixed effects models with age as a fixed effect, and random effects for participant (intercept) and age (slope), generated independently for controls and mutation carriers. All available data points were used in this analysis.

### Data Availability

The data that support the findings of this study are available on request from the corresponding author, EJW. The data are not publicly available due to their containing information that could compromise the privacy of research participants.

## Results

### Participant Characteristics

At baseline our cohort consisted of 15 healthy controls and 44 Huntington’s gene expansion carriers, of whom 15 had premanifest and 29 manifest Huntington’s disease. 3 manifest participants had MRS with an SNR value of <6 so were deemed to be of poor quality and removed. Groups were equally matched for gender (χ^2^ = 0.002, *p* = 0.99) and CAG repeat length (among gene expansion carriers), but as expected, displayed significant differences in clinical, cognitive, imaging and biofluid measures (Table 2). A significant difference in age was observed with manifest patients being significantly older than premanifest patients due to being more advanced in their disease course.

**Table 2.** Participant Group Demographics at Baseline.

|  | CTR |  | PreHD |  | HD |  | Model<br>*p*-value | CTR vs<br>PreHD<br>*p*-value | PreHD<br>vs HD<br>*p*-value |
| --- | --- | --- | --- | --- | --- | --- | --- | --- | --- |
|  | n | Mean ± SD | n | Mean ± SD | n | Mean ± SD |  |  |  |
| Demographics |  |  |  |  |  |  |  |  |  |
| Age (Years) | 15 | 49.2 ± 10.5 | 15 | 42.6 ± 11.8 | 26 | 55.7 ± 9.2 | 0.001 | 0.08 | <0.001 |
| Sex (M/F) | 15 | 8/7 | 15 | 8/7 | 26 | 14/12 | 0.99 | N/A | N/A |
| CAG | N/A | N/A | 15 | 42.1 ± 1.6 | 26 | 42.5 ± 1.8 | 0.48 | N/A | 0.48 |
| Clinical Scores |  |  |  |  |  |  |  |  |  |
| cUHDRS | 15 | 17.5 ± 1.4 | 15 | 18.0 ± 1.2 | 26 | 11.1 ± 3.7 | <0.001 | 0.62 | <0.001 |
| DBS | N/A | N/A | 15 | 275.4 ± 69.2 | 26 | 382.5 ± 84.5 | <0.001 | N/A | <0.001 |
| TFC | 15 | 13.0 ± 0.0 | 15 | 13.0 ± 0.0 | 26 | 9.8 ± 2.7 | <0.001 | 1.00 | <0.001 |
| TMS | 15 | 1.8 ± 1.5 | 15 | 2.5 ± 2.7 | 26 | 32.8 ± 16.6 | <0.001 | 0.86 | <0.001 |
| Cognitive Scores |  |  |  |  |  |  |  |  |  |
| SDMT | 15 | 50.9 ± 10.2 | 15 | 55.5 ± 9.9 | 26 | 30.0 ± 13.4 | <0.001 | 0.29 | <0.001 |
| SWR | 15 | 101.4 ± 17.9 | 15 | 105.0 ± 13.1 | 26 | 64.8 ± 23.5 | <0.001 | 0.62 | <0.001 |
| VFC | 15 | 23.9 ± 4.3 | 15 | 23.7 ± 3.0 | 26 | 15.3 ± 6.1 | <0.001 | 0.91 | <0.001 |
| SCN | 15 | 75.1 ± 13.2 | 15 | 81.5 ± 11.6 | 26 | 48.3 ± 17.0 | <0.001 | 0.24 | <0.001 |
| Imaging Measures (mL, adjusted for total intracranial volume) |  |  |  |  |  |  |  |  |  |
| Whole brain | 15 | 1195.0 ± 56.4 | 15 | 1187.5 ± 51.3 | 25 | 1062.2 ± 73.7 | <0.001 | 0.75 | <0.001 |
| Caudate volume | 12 | 7.1 ± 0.8 | 14 | 6.1 ± 1.2 | 24 | 3.9 ± 1.1 | <0.001 | 0.02 | <0.001 |
| Grey matter | 15 | 705.2 ± 53.0 | 15 | 709.5 ± 48.0 | 26 | 602.8 ± 59.4 | <0.001 | 0.83 | <0.001 |
| White matter | 15 | 438.5 ± 34.3 | 15 | 430.4 ± 29.1 | 26 | 386.0 ± 36.3 | <0.001 | 0.52 | <0.001 |
| Biofluid Measures (log pg/ml, unless stated otherwise) |  |  |  |  |  |  |  |  |  |
| CSF NfL | 15 | 5.9 ± 0.6 | 15 | 6.8 ± 0.8 | 26 | 7.7 ± 0.4 | <0.001 | <0.001 | <0.001 |
| CSF mHTT (fM) | N/A | N/A | 15 | 35.0 ± 21.1 | 26 | 50.3 ± 22.8 | <0.05 | N/A | <0.05 |
| CSF Tau | 15 | 4.2 ± 0.3 | 15 | 4.3 ± 0.3 | 26 | 4.5 ± 0.4 | <0.05 | 0.62 | <0.05 |
| Plasma NfL | 15 | 1.9 ± 0.4 | 15 | 2.4 ± 0.6 | 26 | 3.3 ± 0.4 | <0.001 | <0.01 | <0.001 |
| Plasma Tau | 15 | 1.7 ± 0.4 | 15 | 1.6 ± 0.4 | 26 | 1.6 ± 0.5 | 0.81 | 0.71 | 0.81 |

Intergroup differences were assessed using general linear models and Pearson's chi squared test (Gender). *P*-values are not adjusted for multiple comparisons. Models do not control for age or CAG repeat length. CTR, healthy controls; PreHD, premanifest mutation carriers; HD, manifest mutation carriers; cUHDRS, composite Unified Huntington's Disease Rating Scale; DBS, Disease Burden Score; TFC, Total Functional Capacity; TMS, Total Motor Score; SDMT, Symbol Digit Modalities Test; SWR, Stroop Word Reading Test; VFC, Verbal Fluency Categorical; SCN, Stroop Colour Naming; NfL, Neurofilament light; mHTT, mutant Huntingtin; NA, not applicable.

Our follow-up cohort was smaller, consisting of 12 healthy controls and 36 gene expansion carriers (13 premanifest and 23 manifest), but largely similar in terms of demographics. No subjects were removed due to poor SNR value. Relationships that were significant in the baseline sample were also significant in the follow-up cohort (Supplementary Table 2).

When analysing GABA, 8 subjects were removed at baseline and 6 at follow-up due to high %SD values (≥100), resulting in a smaller sample size (n=48 at baseline and 42 at follow-up) for this metabolite.

### Analysis of Metabolite Group Differences

Baseline analysis in healthy controls revealed MI to be significantly associated with age (*r* = 0.64, *p* = 0.01) and tCho to display significant gender differences (mean difference = -0.11, *p* = 0.04). Therefore, in addition to age, gender was included as a covariate in all subsequent baseline analysis of tCho. In the follow-up cohort, no significant relationships were observed, and gender was not controlled for when analysing tCho (Supplementary Table 3).

At baseline, we found no significant differences in metabolite levels between groups when controlling for the effects of age, and age and CAG. However, we observed trends towards reduced tNAA and tCre as disease progresses (Fig. 2; Table 3). Analysis in the follow-up cohort produced similar results, although tCre displayed significantly lower concentration in manifest, compared to premanifest, patients (*p* = 0.02). This relationship did not reach statistical significance when controlling for age and CAG repeat length (Supplementary Table 4; Supplementary Fig. 2).

**Fig. 2:**
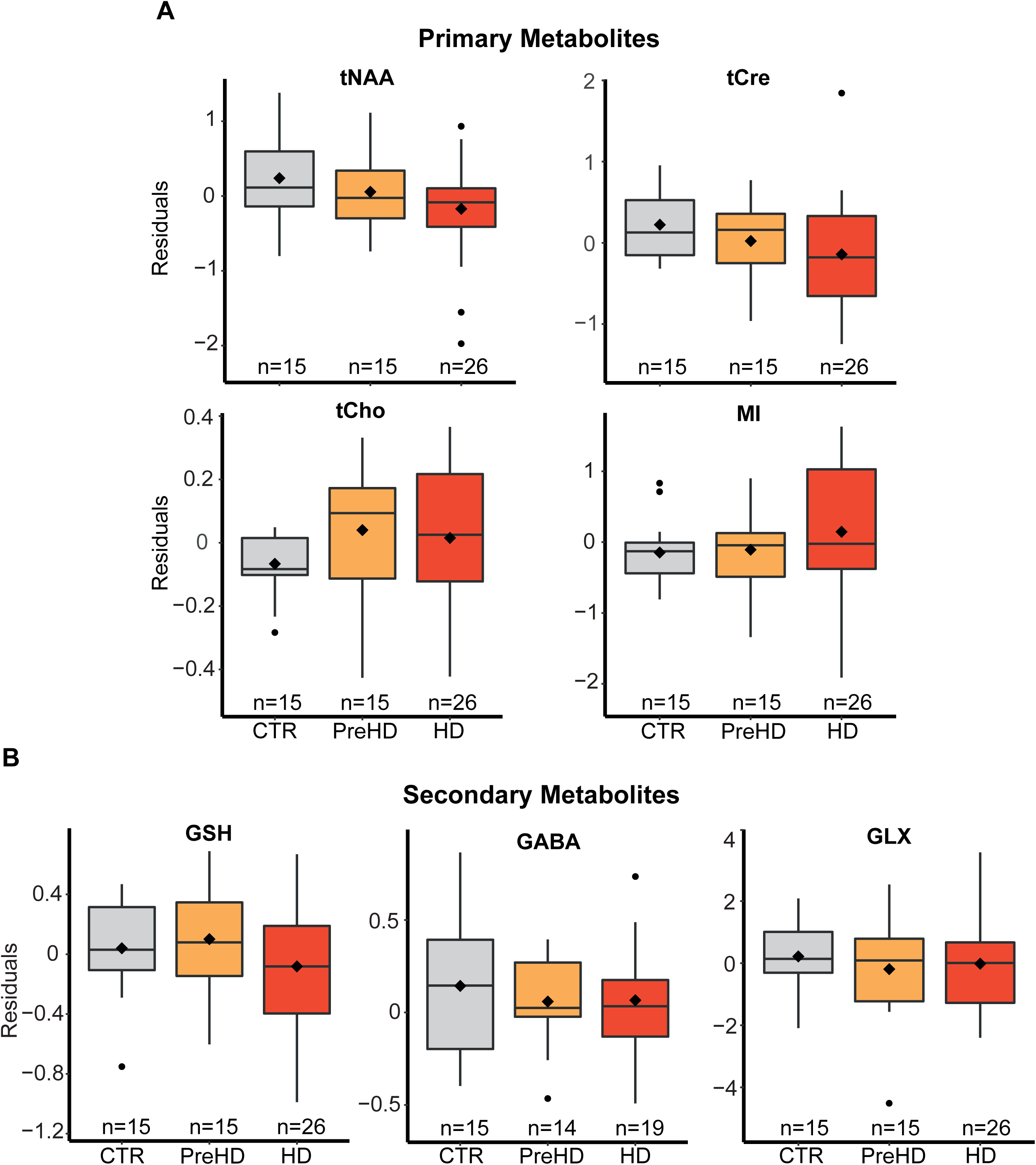
Group differences in metabolite concentration. No significant differences were observed in primary (**A**) or secondary (**B**) metabolites between controls, premanifest and manifest patients. Group membership main effect p-values are displayed in Table 3. When controlling for age and CAG, no significant group differences were observed. Residual values are displayed after controlling for CSF PVE only. Diamonds represent mean values. Tests were not corrected for multiple comparisons.

**Table 3.**
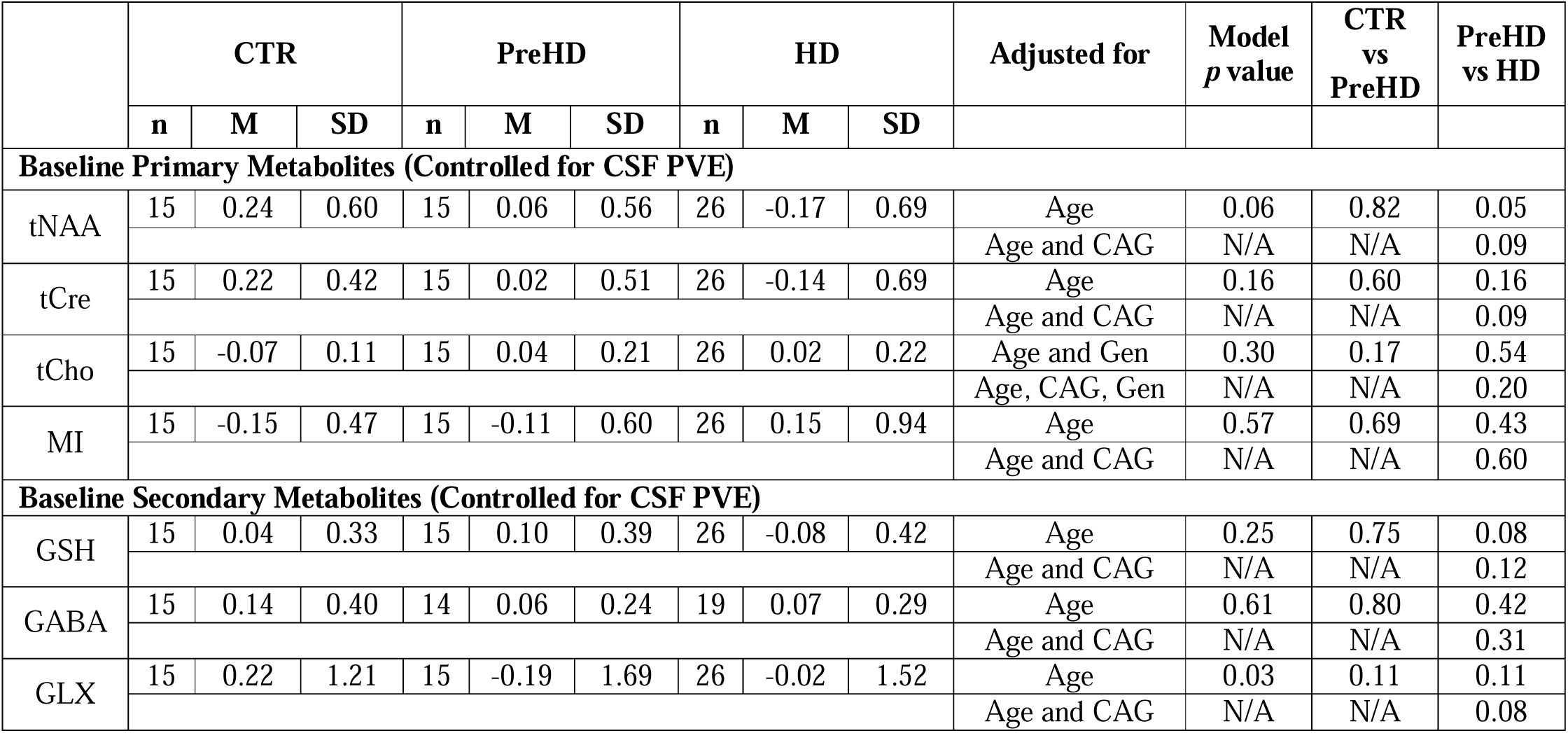
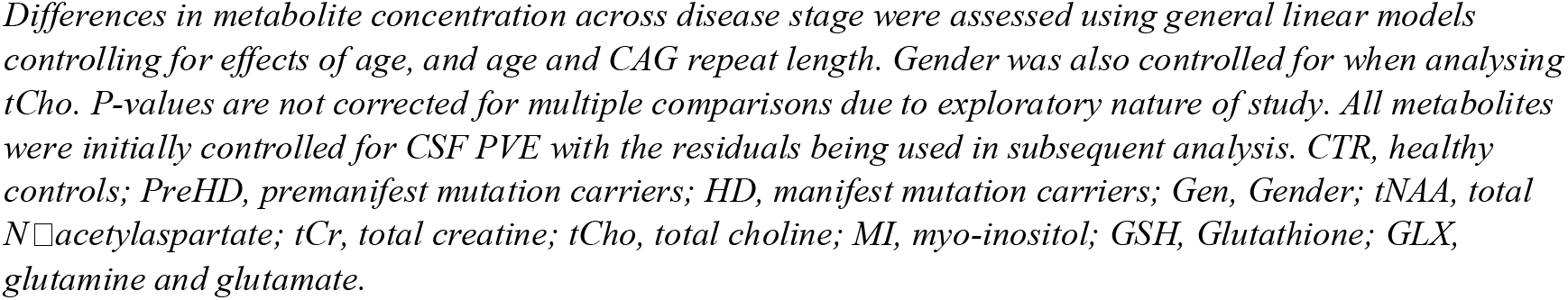
Intergroup Differences in Primary and Secondary Metabolites at Baseline.

|  | CTR |  |  | PreHD |  |  | HD |  |  | Adjusted for | Model<br>*p* value | CTR<br>vs<br>PreHD | PreHD<br>vs HD |
| --- | --- | --- | --- | --- | --- | --- | --- | --- | --- | --- | --- | --- | --- |
|  | n | M | SD | n | M | SD | n | M | SD |  |  |  |  |
| Baseline Primary Metabolites (Controlled for CSF PVE) |  |  |  |  |  |  |  |  |  |  |  |  |  |
| tNAA | 15 | 0.24 | 0.60 | 15 | 0.06 | 0.56 | 26 | -0.17 | 0.69 | Age | 0.06 | 0.82 | 0.05 |
|  |  |  |  |  |  |  |  |  |  | Age and CAG | N/A | N/A | 0.09 |
| tCre | 15 | 0.22 | 0.42 | 15 | 0.02 | 0.51 | 26 | -0.14 | 0.69 | Age | 0.16 | 0.60 | 0.16 |
|  |  |  |  |  |  |  |  |  |  | Age and CAG | N/A | N/A | 0.09 |
| tCho | 15 | -0.07 | 0.11 | 15 | 0.04 | 0.21 | 26 | 0.02 | 0.22 | Age and Gen | 0.30 | 0.17 | 0.54 |
|  |  |  |  |  |  |  |  |  |  | Age, CAG, Gen | N/A | N/A | 0.20 |
| MI | 15 | -0.15 | 0.47 | 15 | -0.11 | 0.60 | 26 | 0.15 | 0.94 | Age | 0.57 | 0.69 | 0.43 |
|  |  |  |  |  |  |  |  |  |  | Age and CAG | N/A | N/A | 0.60 |
| Baseline Secondary Metabolites (Controlled for CSF PVE) |  |  |  |  |  |  |  |  |  |  |  |  |  |
| GSH | 15 | 0.04 | 0.33 | 15 | 0.10 | 0.39 | 26 | -0.08 | 0.42 | Age | 0.25 | 0.75 | 0.08 |
|  |  |  |  |  |  |  |  |  |  | Age and CAG | N/A | N/A | 0.12 |
| GABA | 15 | 0.14 | 0.40 | 14 | 0.06 | 0.24 | 19 | 0.07 | 0.29 | Age | 0.61 | 0.80 | 0.42 |
|  |  |  |  |  |  |  |  |  |  | Age and CAG | N/A | N/A | 0.31 |
| GLX | 15 | 0.22 | 1.21 | 15 | -0.19 | 1.69 | 26 | -0.02 | 1.52 | Age | 0.03 | 0.11 | 0.11 |
|  |  |  |  |  |  |  |  |  |  | Age and CAG | N/A | N/A | 0.08 |

Differences in metabolite concentration across disease stage were assessed using general linear models controlling for effects of age, and age and CAG repeat length. Gender was also controlled for when analysing tCho. P-values are not corrected for multiple comparisons due to exploratory nature of study. All metabolites were initially controlled for CSF PVE with the residuals being used in subsequent analysis. CTR, healthy controls; PreHD, premanifest mutation carriers; HD, manifest mutation carriers; Gen, Gender; tNAA, total N-acetylaspartate; tCr, total creatine; tCho, total choline; MI, myo-inositol; GSH, Glutathione; GLX, glutamine and glutamate.

### Correlation Analysis of Metabolites and measures of Disease Progression

When controlling for age, we found MI to display a strong negative correlation with cUHDRS, in both the weighted and bootstrapped analysis. When additionally controlling for CAG repeat length, this relationship no longer achieved statistical significance (Fig. 3; S5 Supplementary Table 5; Supplementary Fig. 3).

**Fig. 3:**
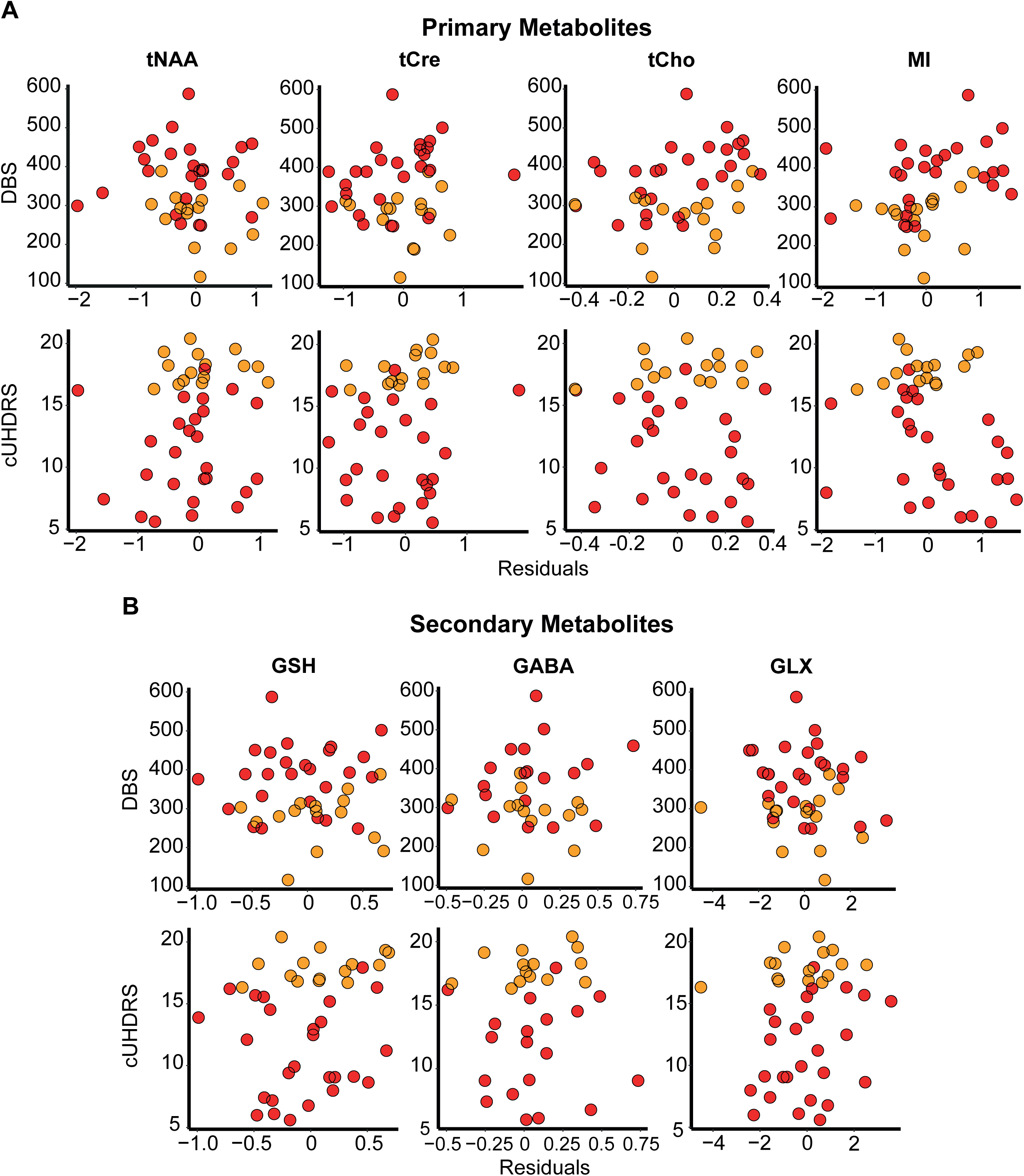
Associations between metabolites and measures of clinical progression (baseline cohort). When controlling for age, MI displayed a significant negative association with cUHDRS. When additionally controlling for CAG, the correlation did not achieve statistical significance (S5 Table). Gender was included in the model when analysing tCho. Values displayed are controlled for CSF PVE only. Red and orange datapoints indicate manifest and premanifest patients, respectively.

In the follow-up cohort, we found reduced levels of tCre and GLX to be significantly associated with a reduction in cUHDRS scores, indicative of a worsening clinical phenotype. The relationships remained significant across all 4 correlation models (tCre, *r* ≥ 0.41, *P* ≤ 0.01; GLX, *r* ≥ 0.36, *P* ≤ 0.03) (*S5 Table*).

There were no statistically significant associations between any metabolite and DBS.

### Correlation Analysis of Metabolites and Cognitive and Imaging Measures

When controlling for age, we observed significant relationships between several metabolites and cognitive and imaging measures in the baseline cohort (Fig. 4; Supplementary Table 5; Supplementary Fig. 3). Most notably, increased MI was significantly associated with a worsening clinical picture, cognitive decline, and volumetric reductions. When controlling for both age and CAG repeat length, many of the observed relationships did not reach statistical significance. However, several relationships survived across all models, with reduced levels of tNAA being associated with reduced caudate volume (*r* ≥ 0.29, *P* ≤ 0.04) and SCN score (*r* ≥ 0.30, *P* ≤ 0.04), and reduced tCre displaying a positive relationship with caudate volume only (*r* ≥ 0.34, *P* ≤ 0.02). Furthermore, the negative association between MI and caudate volume (*r* ≤ -0.32, *P* ≤ 0.05) remained, further highlighting the relationship between increased neuroinflammatory response and neurodegenerative processes in Huntington’s disease.

**Fig. 4:**
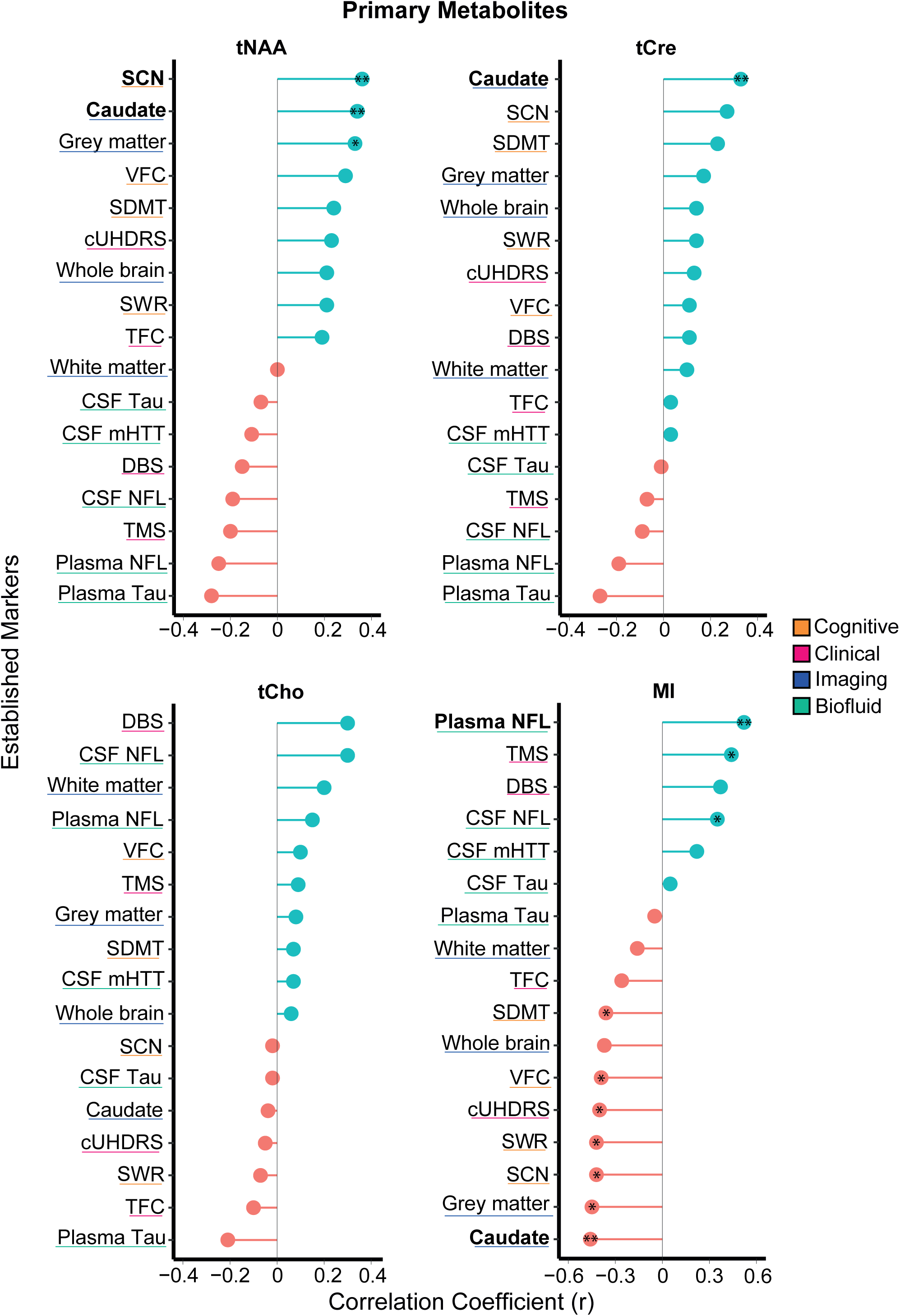
Associations between primary metabolites and clinical, imaging and biofluid measures (baseline cohort). Correlation coefficients were generated using Pearson’s partial correlation controlling for age and bootstrapped with 1000 repetitions. Gender was included in the model when analysing tCho. Single (*) and double stars (**) represent statistical significance (p < 0.05) when controlling for age, and age and CAG repeat length, respectively, in both the weighted and bootstrapped analyses. Bold text also indicates significance across all 4 correlation models.

In the follow-up cohort, only two relationships observed at baseline were replicated, with tCre (*r* ≥ 0.41, *P* ≤ 0.01) and MI (*r* ≤ -0.35, *P* ≤ 0.04) continuing to display significant correlations with caudate volume across all four models. Additionally, tCre, GLX and MI all displayed significant correlations with multiple measures; however, due to the exploratory nature of this study and resulting lack of multiplicity testing, these results should be interpreted with caution (Supplementary Table 5; Supplementary Fig. 4).

### Correlation Analysis of Metabolites and Established Biofluid Biomarkers

When assessing the relationships between metabolites and the established biofluid markers; CSF NfL, CSF mHTT, CSF tau, plasma NfL and plasma tau, only MI displayed significant associations across all 4 models in the baseline cohort. When controlling for age, strong positive correlations were observed with both CSF and plasma NfL in both the weighted and bootstrapped analyses; however, when additionally controlling for CAG repeat length, only the relationship with plasma NfL remained significant across both models (*r* ≥ 0.39, *P* ≤ 0.02), further reflecting the contribution of excessive neuroinflammatory response on disease pathogenesis (Fig. 4; Supplementary Table 5; Supplementary Fig. 3).

Cross-sectional analysis in the follow-up cohort did not replicate any of the findings observed at baseline. Additional relationships were revealed however, with GLX and tCho displaying significant inverse correlations with mHTT (*r* ≤ -0.47, *P* ≤ 0.01) and CSF tau (*r* ≤ -0.38, *P* ≤ 0.02), respectively. Most notably, negative associations between tCre and multiple biofluid markers were observed across all models (mHTT, *r* ≤ -0.47, *P* ≤ 0.01; CSF tau, *r* ≤ -0.30, *P* ≤ 0.04; CSF NfL, *r* ≤ -0.51, *P* ≤ 0.01; Plasma NfL, *r* ≤ -0.51, *P* ≤ 0.01) (Supplementary Table 5; Supplementary Fig. 4). Due to the lack of multiplicity testing, we cannot rule out false positives, thus further validation is required.

### Longitudinal Analysis of Metabolites

Out of the 56 subjects at baseline, 42 had MRS data available at 24-month follow-up. 9 subjects were removed due to high %SD values in GABA, resulting in a smaller sample size (n=33) for this metabolite.

The rate of change did not differ across disease stage for any of the primary metabolites. For the secondary metabolites, we found manifest patients to display a greater rate of change in GLX compared to premanifest patients (Table 4, Fig. 5).

**Table 4.** Annual Rate of Change in all MRS Metabolites.

|  | CTR |  |  | PreHD |  |  | HD |  |  | Adjusted for | Model<br>*p* value | CTR<br>vs<br>PreHD | PreHD<br>vs HD |
| --- | --- | --- | --- | --- | --- | --- | --- | --- | --- | --- | --- | --- | --- |
|  | n | M | SD | n | M | SD | n | M | SD |  |  |  |  |
| Annual Rate of Change in Primary Metabolites (per annum) |  |  |  |  |  |  |  |  |  |  |  |  |  |
| tNAA | 11 | 0.17 | 0.45 | 12 | -0.04 | 0.44 | 19 | -0.05 | 0.42 | Age | 0.60 | 0.30 | 0.79 |
|  |  |  |  |  |  |  |  |  |  | Age and CAG | N/A | N/A | 0.49 |
| tCre | 11 | 0.23 | 0.40 | 12 | 0.07 | 0.40 | 19 | 0.02 | 0.36 | Age | 0.15 | 0.73 | 0.21 |
|  |  |  |  |  |  |  |  |  |  | Age and CAG | N/A | N/A | 0.23 |
| tCho | 11 | 0.09 | 0.19 | 12 | 0.02 | 0.15 | 19 | 0.02 | 0.13 | Age | 0.02 | 0.81 | 0.16 |
|  |  |  |  |  |  |  |  |  |  | Age and CAG | N/A | N/A | 0.11 |
| MI | 11 | -0.01 | 0.41 | 12 | 0.30 | 0.21 | 19 | 0.03 | 0.50 | Age | 0.97 | 0.75 | 0.72 |
|  |  |  |  |  |  |  |  |  |  | Age and CAG | N/A | N/A | 0.81 |
| Annual Rate of Change in Secondary Metabolites (per annum) |  |  |  |  |  |  |  |  |  |  |  |  |  |
| GSH | 11 | 0.14 | 0.54 | 12 | -0.07 | 0.41 | 19 | 0.09 | 0.41 | Age | 0.35 | 0.16 | 0.22 |
|  |  |  |  |  |  |  |  |  |  | Age and CAG | N/A | N/A | 0.17 |
| GABA | 11 | 0.15 | 0.41 | 11 | -0.00 | 0.23 | 11 | -0.07 | 0.25 | Age | 0.81 | 0.61 | 0.72 |
|  |  |  |  |  |  |  |  |  |  | Age and CAG | N/A | N/A | 0.62 |
| GLX | 11 | 0.15 | 1.06 | 12 | 0.68 | 1.59 | 19 | 0.03 | 1.10 | Age | 0.25 | 0.16 | 0.05 |
|  |  |  |  |  |  |  |  |  |  | Age and CAG | N/A | N/A | **0.02** |

*Differences in annual rate of change across disease stage were assessed using general linear models controlling for effects of age, and age and CAG repeat length. P-values are not corrected for multiple comparisons due to exploratory nature of study. CTR, healthy controls; PreHD, premanifest mutation carriers; HD, manifest mutation carriers; tNAA, total N-acetylaspartate; tCr, total creatine; tCho, total choline; MI, myo-inositol; GSH, Glutathione; GLX, glutamine and glutamate.*

**Fig. 5:**
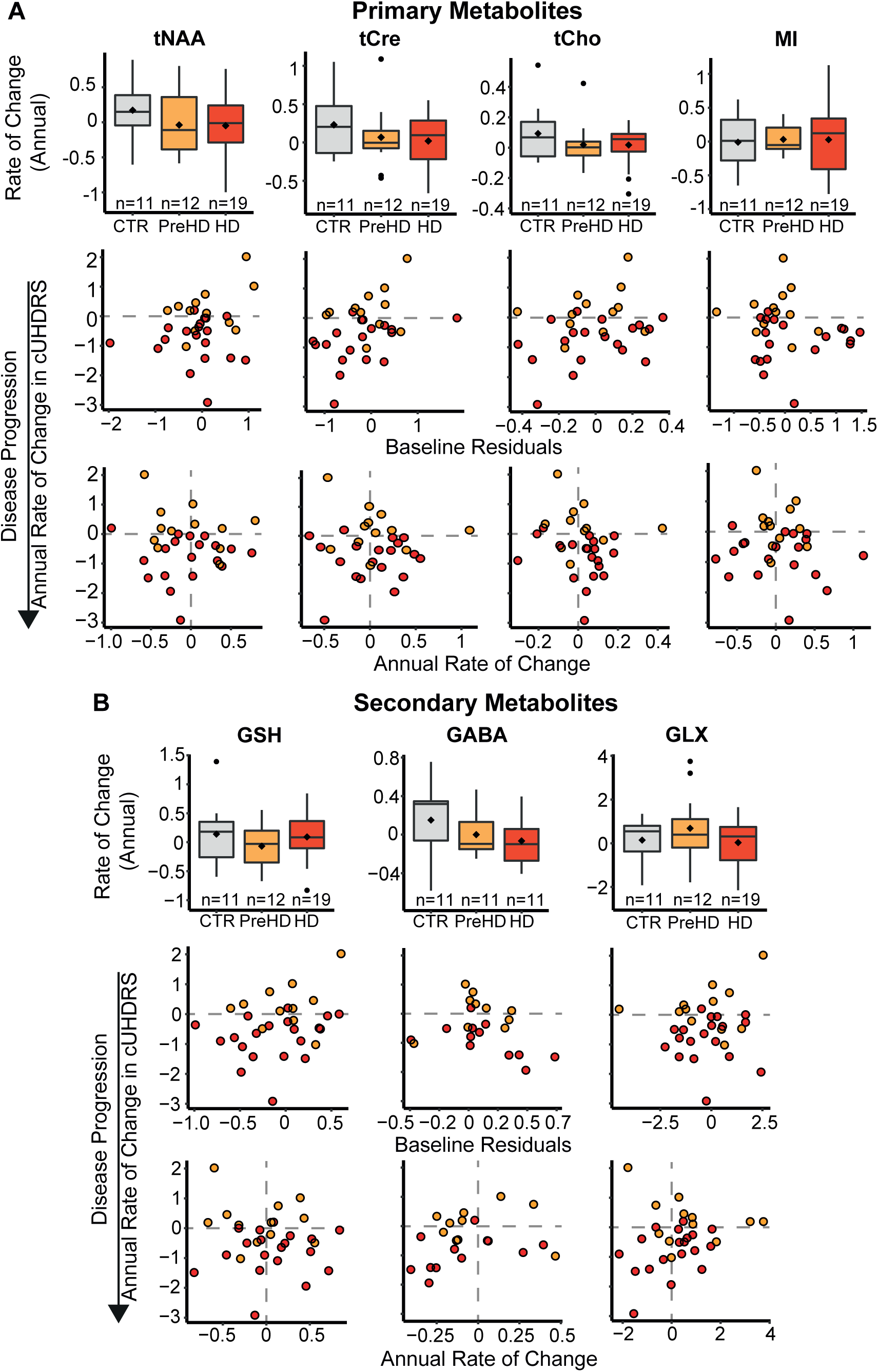
Metabolite rate of change and longitudinal associations with disease progression. Annualised rate of change in MRS metabolites is displayed across disease stage for both primary (**A**) and secondary (**B**) metabolites (Top Panels). Model outputs are displayed in Table 3. Associations between baseline values (Middle Panels) or annualised rate of change (Bottom Panels) in each metabolite, with annual rate of change in cUHDRS are also displayed. Dashed horizontal lines represent no change in cUHDRS, negative values indicate disease progression. Dash vertical lines represent no change in metabolites. Data points are controlled for CSF PVE only. Pearson’s partial correlation coefficient controlling for age, and age and CAG, and bias-corrected bootstrapped confidence intervals are displayed in Supplementary Table 6 and 7. Red, orange, and grey datapoints indicate manifest patients, premanifest patients, and healthy controls, respectively.

To assess prognostic value of the metabolites, we examined if baseline values predicted subsequent change in established measures of disease progression. When controlling for all covariates, we found tCre to display a significant positive correlation with change in cUHDRS (*r* ≥ 0.36, *P* ≤ 0.03), indicating significant predictive power independent of the core genetic mutation (Fig. 5, Supplementary Table 6). Several additional relationships were also observed, most notably with MRI measures; however, only two remained significant across all 4 correlation models, with MI significantly predicting decline in white matter volume (*r* ≤ -0.44, *P* ≤ 0.02) and GSH associating with change in whole brain volume (*r* ≥ 0.36, *P* ≤ 0.04) (Supplementary Fig. 5, Supplementary Table 6).

To assess if rate of change in metabolites provided additional prognostic behaviour beyond that observed using baseline values, we correlated metabolite rate of change, with the rate of change in markers of disease progression (Fig. 5). Findings were limited, with tCre displaying significant correlations with grey matter volume (*r* ≥ 0.38, *P* ≤ 0.02) and GLX displaying relationships with grey matter volume (r ≥ 0.39, *P* ≤ 0.04) and TMS (*r* ≤ -0.36, *P* ≤ 0.03), across all models (Supplementary Fig. 6, Supplementary Table 7).

Longitudinal trajectories of each metabolite within individuals are displayed in Fig. 6. Longitudinal mixed effects models, controlling for age, revealed no significant change in metabolite concentration over time in Huntington’s disease mutation carriers.

**Fig. 6:**
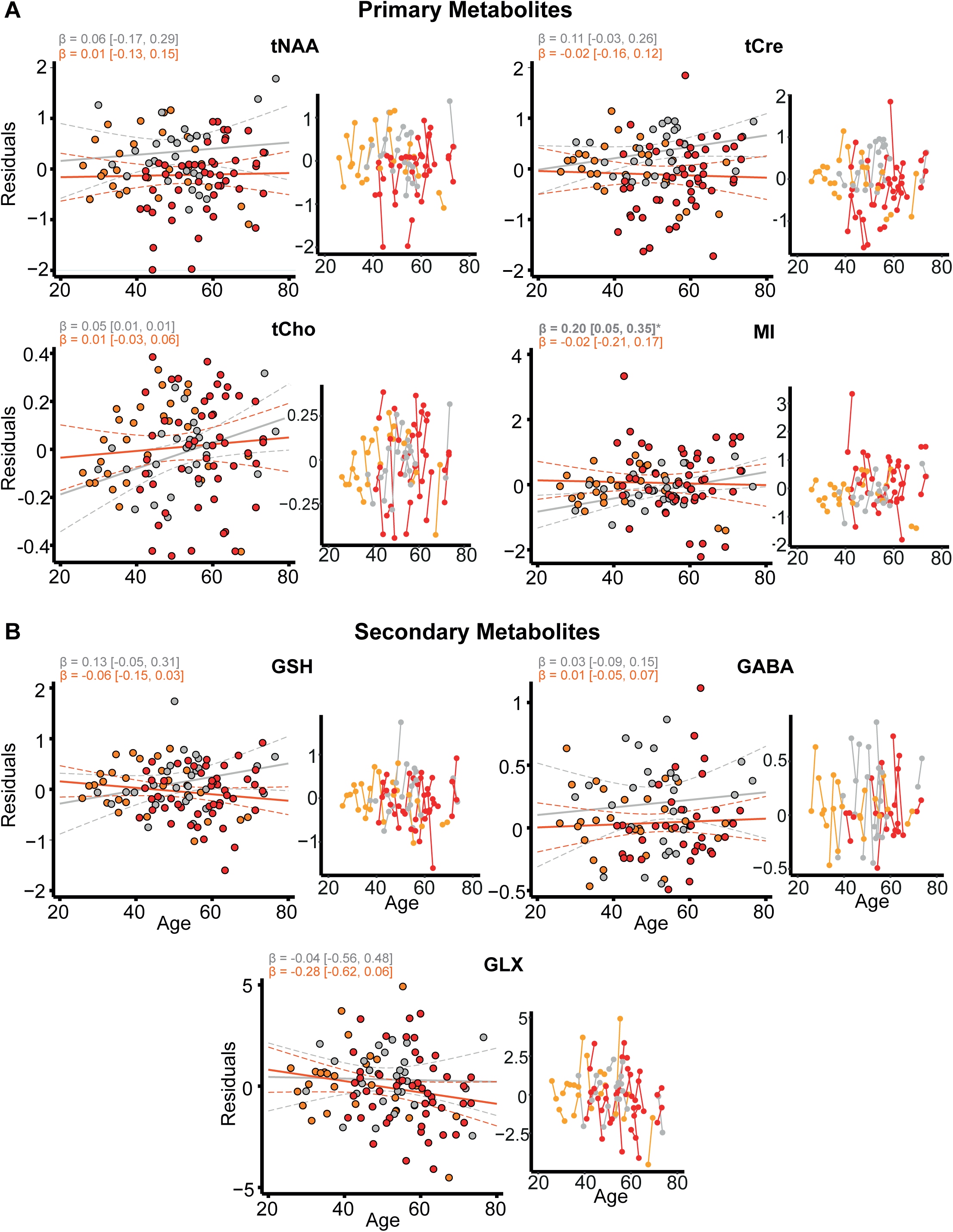
Longitudinal analysis of MRS metabolites. Longitudinal trajectories (Left Panel) of all metabolites were studied controlling for age (controls), and age and CAG (mutation carriers). Model (solid lines) and 95% (dashed lines) confidence intervals were generated from generalized mixed-effects models. Bold text and ‘*’ indicate significance at p < 0.05. Beta values and 95% confidence intervals have been multiplied by 10 to show change/10yrs. Individual participant trajectories are displayed in the right panels with connected dots representing the same participant. Red, orange, and grey datapoints indicate manifest patients, premanifest patients, and healthy controls, respectively.

## Discussion

In this study, we employed 3T magnetic resonance spectroscopy to successfully quantify seven metabolites in the putamen of Huntington’s disease patients and healthy controls. We specified the most prominent metabolites in the ^1^H spectrum – tNAA, tCre, tCho and MI – as primary metabolites and included lesser studied metabolites – GABA, GLX and GSH – as secondary metabolites. In keeping with previous work, metabolites were normalised to unsuppressed water signal,^25,28,37,56^ allowing for increased accuracy when identifying changes in brain biochemistry^56^ and additionally controlled for CSF partial volume effect. Using general linear models and correlation analysis, we assessed their potential as prognostic and diagnostic biomarkers, both cross sectionally and longitudinally, by exploring their relationships with established markers of disease progression, cognitive decline, and brain atrophy. Furthermore, we studied the relationship between MRS metabolites and several biomarkers derived from CSF and plasma, including NfL and the pathogenic protein, mHTT. To our knowledge, such relationships have not been explored in Huntington’s disease.

When controlling for age and CAG, we observed no discernible group differences in any metabolite concentration across both time points. This finding contrasts with those of Sturrock et al.,^25,37^ who employed a similar methodology in a larger cohort and found tNAA concentration to be reduced, suggestive of neuronal dysfunction, in manifest compared to premanifest patients, and between premanifest patients and healthy controls across time points. In our baseline cohort, we observed non-significantly (p=0.05) reduced tNAA in manifest compared to premanifest patients; this was not replicated at the follow-up timepoint, likely due to lack of study power. Furthermore, Sturrock et al. found MI to be increased in manifest compared with premanifest patients at baseline, 12- and 24-month follow-up. Our MI results did not support this and may reflect methodological differences between the studies, specifically our adjustment for the effects of CSF partial volume effects, and inclusion of both age and CAG repeat length as covariates in all models. However, in keeping with Sturrock et al’s. findings, we did not observe any significant differences in MI concentration between premanifest patients and controls, and tCre was found to be significantly reduced in manifest compared to premanifest patients in the follow-up cohort; however, this relationship did not survive when additionally controlling for CAG repeat length. Reduced creatine has been consistently demonstrated in Huntington’s disease patients^25,28,35^ and may demonstrate diagnostic potential but will require further study in larger samples.

Our study did not find any significant group differences when comparing premanifest patients to healthy controls. This supports earlier work,^26,34^ in addition to an exploratory study leveraging 7T MRI,^35^ in which concentrations of creatine, choline, MI, tNAA, GLX and lactate did not differ in the putamen of premanifest patients and controls, in addition to 4 other distinct brain regions. Our findings may reflect the fact that our premanifest group were clinically well, demonstrating no significant differences in clinical, cognitive, or imaging measurements compared with healthy controls, whereas other studies may have included premanifest patients closer to clinical onset or with prodromal disease.

In our cross-sectional correlation analysis, MI was significantly associated with caudate volume in both baseline and follow-up cohorts, and plasma NfL at baseline only. To our knowledge, this represents the first data in Huntington’s disease patients relating non-invasive MRS measures to an established biofluid marker of disease progression. MI reflects astrocytic density, while NfL reflects neuro-axonal injury from any mechanism.^57–59^ This association perhaps reflects astrocytic involvement in neuroinflammation or in compensating for neurodegeneration.^60–63^ However, this finding was not replicated in the follow-up cohort and will require further study to better elucidate the relationship between the two measures.

We also observed tCre to be significantly associated with caudate volume at baseline and follow up, representing the second correlation to be replicated across both time points. At follow up, it was significantly associated with measures of disease progression and cognitive decline, which is in keeping with previous findings.^28,35^ The strongest relationships were seen with cUHDRS, TMS, grey matter volume and SDMT, with reduced tCre indicative of a more severe disease phenotype. Additionally, we found tCre was negatively associated with CSF and plasma NfL, CSF mHTT and CSF tau in the follow-up cohort. Reduced GLX was associated with multiple markers, including CSF mHTT, in the follow up cohort only. Previous work has shown reduced GLX in the putamen of Huntington’s disease patients and its associations with worse performance on the SDMT.^35^ The lack of multiplicity testing means we cannot rule out false positives in this study and the lack of consistency between both cross sectional correlational analyses should be acknowledged; however, these results lend support to the notion that creatine concentration may reflect disease activity in a meaningful way, concordant with many other disease measures, and independently of known predictors, and provides additional evidence for reduced GLX being indicative of a worsening clinical phenotype.

Like Sturrock et al.,^37^ we observed no longitudinal change in any metabolites in Huntington’s disease patients over 24 months. However, we did find that baseline values of tCre significantly predicted subsequent change in cUHDRS, a composite clinical measure sensitive to clinical change.^53^ We also found the rate of change in tCre to predict change in grey matter volume. Both relationships remained significant when controlling for age and CAG. While this predictive potential is of interest, it must be considered in the context of many statistical tests and should therefore be considered exploratory or hypothesis-generating.

We also observed a significant relationship between rate of change in GLX and change in TMS, concordant with our cross-sectional findings showing reduced GLX to be associated with worse cognitive performance. However, given that no relationships were observed with baseline GLX values, this result should be interpreted with caution. Furthermore, we found baseline MI values to associate with annualised rate of change in neuroimaging markers, but only the relationship with white matter volume remained significant across all models. This relationship lends support to earlier work highlighting the link between inflammation and myelin breakdown in manifest patients,^64^ and demonstrates MI’s potential as a marker of axonal degeneration. Interestingly, we also observed a significant relationship between reduced baseline GSH and larger rate of change in whole brain volume. GSH is a major antioxidant known to be dysregulated in Huntington’s disease^65^ and given that glial cell activation has been linked to increased reactive oxygen species (ROS) production,^66^ this finding could represent a cyclic cascade of events whereby increased ROS production due to neuroinflammation is insufficiently buffered by GSH, resulting in oxidative stress and mitochondrial dysfunction, further driving inflammatory pathways and contributing to the neuropathological hallmarks of the disease.

This study is not without its limitations. Due to the exploratory nature of the study, we chose not to adjust our analyses for multiple comparisons. In doing so, we cannot rule out the influence of false positives on our findings. Our decision to adopt more rigorous methodologies also increases the chance of type 2 (false negative) error but lends greater credibility to our findings overall. Although our results provide some evidence supporting the prognostic potential of specific MRS metabolites, there was a lack of consistency between time-points, with the association between tCre and caudate volume being the only relationship to meet all pre-defined tests at baseline and follow up. Consequently, further validation is required in a larger sample. Although HD-CSF is a high-quality longitudinal cohort with biofluid collection and MRI imaging, the sample was principally designed to study manifest Huntingtin’s disease. Previous MRS studies often compared manifest patients, or a combination of manifest and premanifest patients, directly to controls, explored different brain regions and in some cases, normalised values to metabolites thought to be affected in Huntington’s disease^28,30–32^; thus our results may not be directly comparable to earlier work. As a means of quality control, we also excluded some participants based on SNR and %SD values, resulting in a smaller sample size, and reducing the generalisability of the findings. The longitudinal nature of this study is also limited by the small number of available time points. Future studies should aim to incorporate additional time points to help better characterise the longitudinal trajectory of metabolites and improve the models designed to inform on clinical prognosis.

In conclusion, we found no groupwise differences in MRS metabolite concentration when comparing manifest to premanifest Huntington’s disease patients, and premanifest patients to healthy controls. This does not exclude the role of MRS-detectable metabolic dysfunctions in disease pathology, only that their use a state biomarker is limited. We found interesting cross-sectional associations between multiple metabolites, namely tCre, MI and GLX, and markers of disease progression, highlighting the proposed roles of neuroinflammation and metabolic dysfunction in Huntington’s disease pathogenesis, but the inconsistent findings between timepoints and with rigorous statistical modelling suggests these changes, too will have limited biomarker potential. We provide the first evidence, to our knowledge, of an association between MRS metabolites and established CSF biomarkers in gene expansion carriers and, although no longitudinal change in metabolite concentration was observed, we found tCre, MI and GLX to significantly predict change in measures of disease progression, independent of existing predictors. The potential of non-invasive MRS measurements of brain metabolic activity to monitor the progression of Huntington’s disease or the response to therapeutic interventions warrants directed study of these hypotheses in larger longitudinal imaging cohorts linked to biofluid collection, such as the nascent Image-Clarity study, which will add advanced imaging modalities to the large, multi-site HDClarity CSF collection initiative.^67^

## Abbreviations

Cho: choline
Cr: creatine
CTR: healthy controls
cUHDRS: composite Unified Huntington’s Disease Rating Scale
DBS: disease burden score
DCL: diagnostic confidence level
Glu: glutamate
Glx: glutamine + glutamate
GSH: Glutathione
HD: manifest mutation carriers
Lac: Lactate
LLoQ: lower limit of quantification
LoD: limit of detection
mHTT: mutant Huntingtin
MI: myo-inositol
MRS: magnetic resonance spectroscopy
NfL: neurofilament light chain
PreHD: premanifest mutation carriers
PVE: partial volume effect
ROS: reactive oxygen species.
SD: standard deviation
SDMT: Symbol Digit Modality Test
SNR: signal-to-noise ratio
SWR: Stroop Word Reading
tCho: total choline (phosphocholine + glycophosphocholine)
tCre: total creatine (creatine + phosphocreatine)
TFC: Total Functional Capacity
TIV: total intracranial volume
TMS: Total Motor Score
tNAA: total N-acetylaspartate (N-acetylaspartate + N-acetylaspartate-glutamate)
VFC: Verbal Fluency (Categorical)

## Acknowledgements

We would like to thank all the participants from the Huntington’s disease community who donated samples and gave their time to take part in this study.

