## Supplementary Files for "Longitudinal Evaluation of Magnetic Resonance Spectroscopy Metabolites as Biomarkers in Huntington’s Disease"

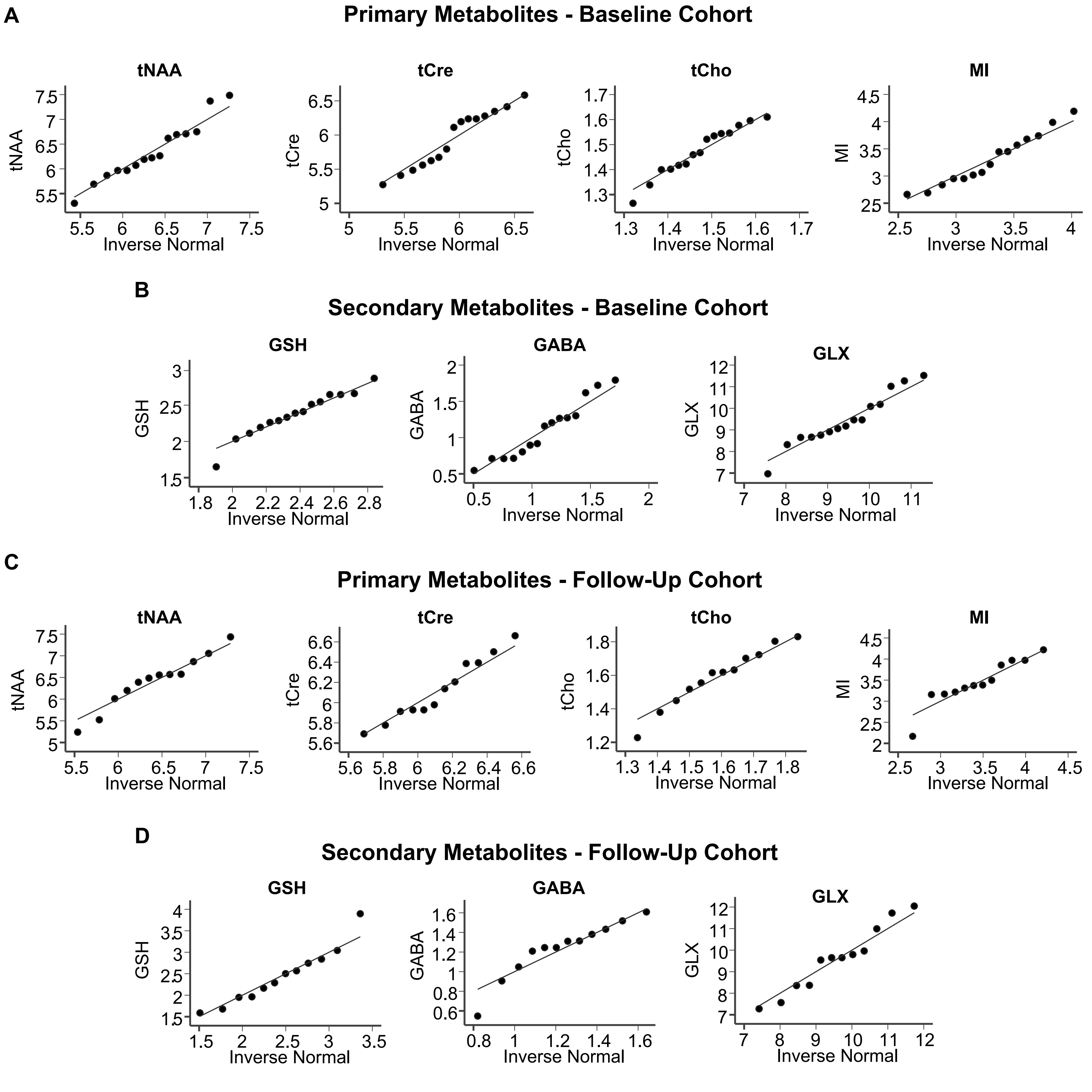


**Supplementary Fig. 1: Q-Q plots for All Metabolites in both Baseline and Follow-Up Cohort**. Q-Q plots obtained from controls only. Neither the primary, nor secondary, metabolites required data transformation to achieve a suitable distribution.

**<u>Supplementary Table 1: MRS Metabolite Quality Control across both Timepoints.</u>**

| Baseline Cohort | % SD > 100 (n) | % SD (Mean) | % SD (Max) | SD > 20% (% of n) |
|---|---|---|---|---|
| **Primary Metabolites** | | | | |
| tNAA | 0 | 4.8 | 10 | 0 |
| tCre | 0 | 3.4 | 6 | 0 |
| tCho | 0 | 4.1 | 7 | 0 |
| MI | 0 | 6.5 | 16 | 0 |
| **Secondary Metabolites** | | | | |
| GSH | 0 | 10.9 | 21 | 0.56 |
| GABA | 8 | 47.7 | 99 | 100 |
| GLX | 0 | 7.9 | 17 | 0 |

| Follow-Up Cohort | % SD > 100 (n) | % SD (Mean) | % SD (Max) | SD > 20% (% of n) |
|---|---|---|---|---|
| **Primary Metabolites** | | | | |
| tNAA | 0 | 5.1 | 7 | 0 |
| tCre | 0 | 3.5 | 7 | 0 |
| tCho | 0 | 4.2 | 9 | 0 |
| MI | 0 | 6.2 | 13 | 0 |
| **Secondary Metabolites** | | | | |
| GSH | 0 | 12.2 | 39 | 1.9 |
| GABA | 6 | 45.8 | 99 | 100 |
| GLX | 0 | 7.9 | 16 | 0 |

*Metabolite measurements with a %SD of >= 100 were removed from all analysis. GABA was the only metabolite with %SD values of >= 100, resulting in the removal of 8 and 6 participants from the baseline and follow-up cohort, respectively. For the remaining metabolites, mean %SD values were <20, indicating high reliability. tNAA, total N-acetylaspartate; tCr, total creatine; tCho, total choline; MI, myo-inositol; GSH, Glutathione; GLX, glutamine and glutamate.*

**Supplementary Table 2. Participant Group Demographics at Follow-Up.**

| | CTR | | PreHD | | HD | | Model *p*-value | CTR vs PreHD *p*-value | PreHD vs HD *p*-value |
|---|---|---|---|---|---|---|---|---|---|
| | n | Mean ± SD | n | Mean ± SD | n | Mean ± SD | | | |
| **Demographics** | | | | | | | | | |
| Age (Years) | 12 | 55.5 ± 10.7 | 13 | 44.7 ± 12.3 | 23 | 58.7 ± 9.0 | 0.001 | 0.01 | <0.001 |
| Sex (M/F) | 12 | 8/4 | 13 | 7/6 | 23 | 11/12 | 0.57 | N/A | N/A |
| **Clinical Scores** | | | | | | | | | |
| cUHDRS | 12 | 17.4 ± 1.4 | 13 | 18.3 ± 1.7 | 22 | 9.7 ± 4.7 | <0.001 | 0.49 | <0.001 |
| DBS | N/A | N/A | 13 | 280.4 ± 64.4 | 23 | 403.6 ± 98.2 | <0.001 | N/A | <0.001 |
| CAG | N/A | N/A | 13 | 42 ± 1.7 | 23 | 42.5 ± 1.9 | 0.41 | N/A | 0.41 |
| TFC | 12 | 13 ± 0 | 13 | 12.9 ± 0.3 | 23 | 8.4 ± 3.7 | <0.001 | 0.94 | <0.001 |
| TMS | 12 | 1.9 ± 1.8 | 13 | 3.3 ± 2.9 | 23 | 40.5 ± 25.0 | <0.001 | 0.84 | <0.001 |
| **Cognitive Scores** | | | | | | | | | |
| SDMT | 12 | 51.3 ± 10.1 | 13 | 58.7 ± 10.7 | 22 | 26.8 ± 14.9 | <0.001 | 0.15 | <0.001 |
| SWR | 12 | 98.2 ± 16.5 | 13 | 107.9 ± 21.6 | 22 | 57.2 ± 26.1 | <0.001 | 0.29 | <0.001 |
| VF | 12 | 25 ± 4.9 | 13 | 23.9 ± 5.0 | 22 | 14.0 ± 6.2 | <0.001 | 0.63 | <0.001 |
| SCN | 12 | 76.2 ± 12.4 | 13 | 80.8 ± 12.2 | 22 | 43.9 ± 19.7 | <0.001 | 0.48 | <0.001 |
| **Imaging Measures (*adjusted for total intracranial volume (ml))** | | | | | | | | | |
| Whole brain | 11 | 1191.0 ± 53.1 | 11 | 1176.8 ± 56.3 | 22 | 1048.2 ± 78.4 | <0.001 | 0.62 | <0.001 |
| Caudate volume | 11 | 6.9 ± 0.7 | 12 | 6.1 ± 1.0 | 22 | 3.6 ± 1.2 | <0.001 | 0.09 | <0.001 |
| Grey matter | 10 | 709.9 ± 43.7 | 12 | 714.1 ± 47.9 | 21 | 592.8 ± 66.2 | <0.001 | 0.86 | <0.001 |
| White matter | 10 | 429.4 ± 32.8 | 12 | 419.2 ± 24.3 | 21 | 378.4 ± 40.4 | <0.001 | 0.50 | 0.002 |
| **Biofluid Measures (log pg/ml, unless stated otherwise)** | | | | | | | | | |
| CSF NfL | 11 | 6.3 ± 0.7 | 13 | 6.9 ± 0.7 | 22 | 8.0 ± 0.5 | <0.001 | <0.05 | <0.001 |
| CSF mHTT (pM) | N/A | N/A | 12 | 34.6 ± 12.2 | 22 | 60.2 ± 29.8 | <0.01 | N/A | <0.01 |
| CSF Tau | 11 | 4.4 ± 0.3 | 13 | 4.4 ± 0.3 | 22 | 4.9 ± 0.4 | <0.001 | 0.86 | <0.01 |
| Plasma NfL | 12 | 2.2 ± 0.5 | 12 | 2.5 ± 0.6 | 23 | 3.4 ± 0.4 | <0.001 | 0.25 | <0.001 |
| Plasma Tau | 12 | 1.9 ± 0.3 | 13 | 2.1 ± 0.2 | 23 | 2.1 ± 0.3 | 0.15 | 0.08 | 0.85 |

*Intergroup differences were assessed using general linear models and Pearson's chi squared test (Gender). P-values are not adjusted for multiple comparisons. Models do not control for age or CAG repeat length. CTR, healthy controls; PreHD, premanifest mutation carriers; HD, manifest mutation carriers; cUHDRS, composite Unified Huntington's Disease Rating Scale; DBS, Disease Burden Score; CAG, CAG triplet repeat count; TFC, Total Functional Capacity; TMS, Total Motor Score; SDMT, Symbol Digit Modalities Test; SWR, Stroop Word Reading Test; VFC, Verbal Fluency Categorical; SCN, Stroop Colour Naming; mHTT, mutant Huntingtin; NA, not applicable.*

### Supplementary Table 3: Assessments for Confounding Variables in All Metabolites

| Baseline Cohort | Age | | Gender | |
|---|---|---|---|---|
| | ***r*** | ***p* value** | ***t*** | ***p* value** |
| **Primary Metabolites (Controlled for CSF PVE)** | | | | |
| tNAA | 0.09 | 0.74 | 0.70 | 0.50 |
| tCre | -0.01 | 0.98 | -1.89 | 0.08 |
| tCho | 0.17 | 0.54 | **-2.31** | **0.04** |
| MI | **0.64** | **0.01** | -1.55 | 0.15 |
| **Secondary Metabolites (Controlled for CSF PVE)** | | | | |
| GSH | 0.48 | 0.07 | -0.65 | 0.53 |
| GABA | -0.02 | 0.95 | 0.07 | 0.95 |
| GLX | -0.26 | 0.34 | -2.11 | 0.06 |

| Follow-Up Cohort | Age | | Gender | |
|---|---|---|---|---|
| | ***r*** | ***p* value** | ***t*** | ***p* value** |
| **Primary Metabolites (Controlled for CSF PVE)** | | | | |
| tNAA | 0.22 | 0.49 | 0.03 | 0.97 |
| tCre | 0.52 | 0.09 | -0.78 | 0.45 |
| tCho | 0.47 | 0.13 | 0.64 | 0.53 |
| MI | 0.37 | 0.24 | 0.37 | 0.72 |
| **Secondary Metabolites (Controlled for CSF PVE)** | | | | |
| GSH | 0.08 | 0.80 | -0.42 | 0.69 |
| GABA | 0.15 | 0.64 | 1.59 | 0.14 |
| GLX | 0.08 | 0.80 | -0.16 | 0.88 |

*Values are Pearson's r and t-test statistic. Bold indicates significance at the $p<0.05$ level. All metabolites were controlled for CSF PVE with resulting residuals used in the analysis. tNAA, total N-acetylaspartate; tCr, total creatine; tCho, total choline; MI, myo-inositol; GSH, Glutathione; GLX, glutamine and glutamate*

<u>**Supplementary Table 4: Intergroup Differences in All Metabolites at Follow-Up**</u>

| | **CTR** | | | **PreHD** | | | **HD** | | | **Adjusted for** | **Model *p* value** | **CTR vs PreHD** | **PreHD vs HD** |
|---|---|---|---|---|---|---|---|---|---|---|---|---|---|
| | **n** | **M** | **SD** | **n** | **M** | **SD** | **n** | **M** | **SD** | | | | |
| **Follow-Up Primary Metabolites (Controlled for CSF PVE)** | | | | | | | | | | | | | |
| tNAA | 12 | 0.44 | 0.66 | 13 | 0.09 | 0.63 | 23 | -0.28 | 0.68 | Age | 0.03 | 0.40 | 0.09 |
| | | | | | | | | | | Age and CAG | N/A | N/A | 0.05 |
| tCre | 12 | 0.50 | 0.39 | 13 | 0.08 | 0.47 | 23 | -0.30 | 0.72 | Age | **0.002** | 0.24 | **0.02** |
| | | | | | | | | | | Age and CAG | N/A | N/A | 0.23 |
| tCho | 12 | 0.05 | 0.17 | 13 | 0.01 | 0.15 | 23 | -0.03 | 0.24 | Age | 0.10 | 0.93 | 0.09 |
| | | | | | | | | | | Age and CAG | N/A | N/A | 0.25 |
| MI | 12 | -0.22 | 0.51 | 13 | -0.10 | 0.62 | 23 | 0.17 | 1.15 | Age | 0.98 | 0.98 | 0.74 |
| | | | | | | | | | | Age and CAG | N/A | N/A | 0.57 |
| **Follow-Up Secondary Metabolites (Controlled for CSF PVE).** | | | | | | | | | | | | | |
| GSH | 12 | 0.26 | 0.67 | 13 | -0.06 | 0.54 | 23 | -0.10 | 0.60 | Age | 0.07 | 0.07 | 0.88 |
| | | | | | | | | | | Age and CAG | N/A | N/A | 0.23 |
| GABA | 12 | 0.27 | 0.29 | 13 | 0.05 | 0.28 | 17 | 0.02 | 0.34 | Age | 0.30 | 0.20 | 0.75 |
| | | | | | | | | | | Age and CAG | N/A | N/A | 0.88 |
| GLX | 12 | 0.36 | 1.64 | 13 | 0.54 | 1.91 | 23 | -0.49 | 2.02 | Age | 0.21 | 0.96 | 0.18 |
| | | | | | | | | | | Age and CAG | N/A | N/A | 0.43 |

*Differences in metabolite concentration across disease stage were assessed using general linear models controlling for effects of age, and age and CAG repeat length. P-values are not corrected for multiple comparisons due to exploratory nature of study. All metabolites were controlled for CSF PVE with the residuals being used in subsequent analysis. CTR, healthy controls; PreHD, premanifest patients; HD, manifest patients; tNAA, total N-acetylaspartate; tCr, total creatine; tCho, total choline; MI, myo-inositol; GSH, Glutathione; GLX, glutamine and glutamate.*

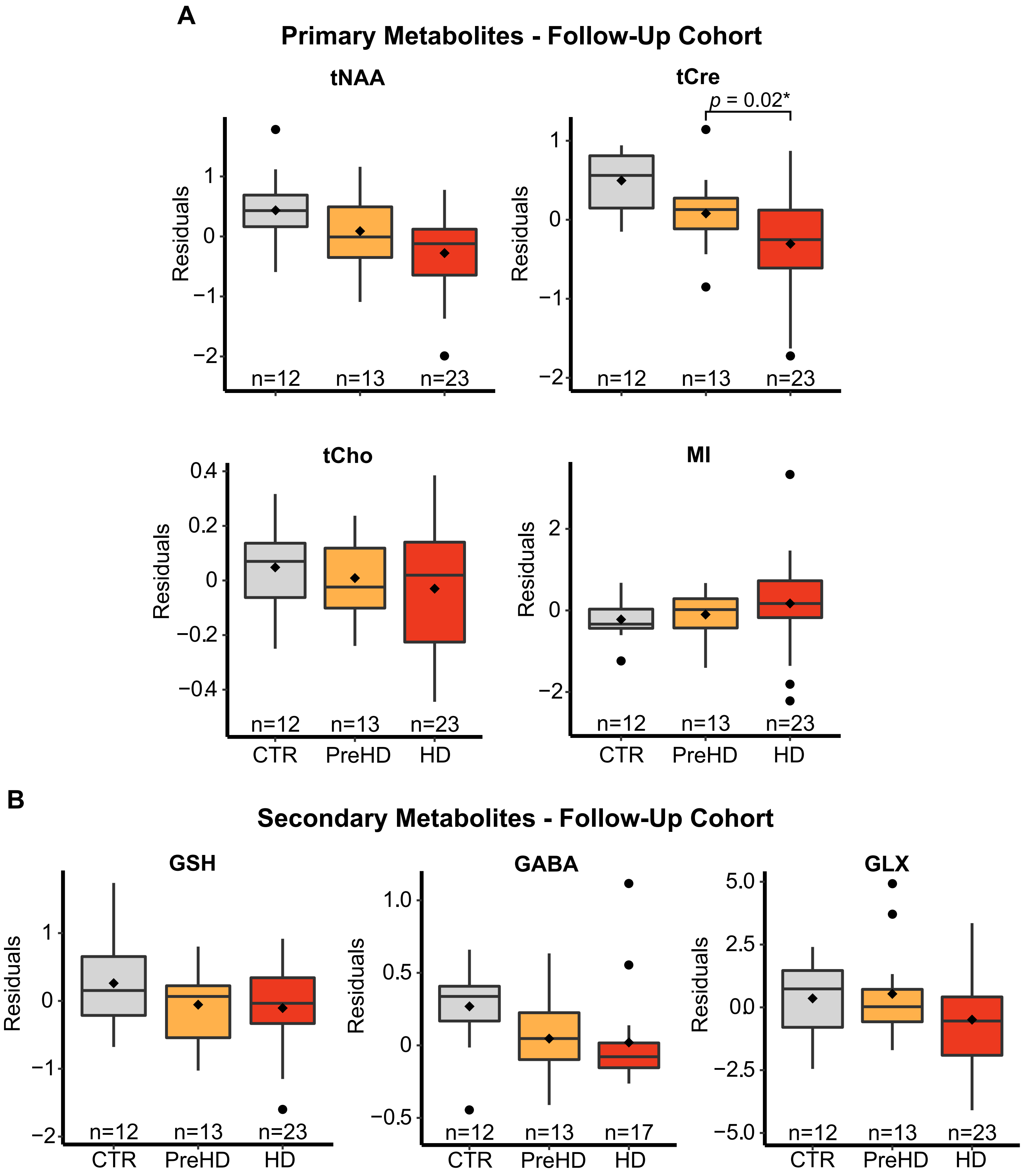


**Supplementary Fig. 2: Intergroup Differences in All Metabolites at Follow-Up (Boxplots)**. Box plots displaying group differences in metabolite concentration. Group membership main effect p-values are displayed in S4 Table. When controlling for age, manifest patients had significantly lower tCre concentration compared with premanifest patients. When additionally controlling for CAG, no significant group differences were observed. Residual values are displayed after controlling for CSF PVE only. Diamonds represent mean values. Tests were not corrected for multiple comparisons

**Supplementary Table 5 – Correlations between Metabolites and Clinical, Cognitive, Imaging and Biofluid Measures in Baseline and Follow-Up Cohorts.**

| **Baseline** | | **Age-adjusted** | | | | | **Age and CAG-adjusted** | | | | |
|---|---|---|---|---|---|---|---|---|---|---|---|
| **tNAA** | | **Inverse weighted** | | **Bootstrapped** | | | **Inverse weighted** | | **Bootstrapped** | | |
| **Measures** | **n** | **r** | ***p* value** | **r** | **95 % CIs** | ***p* value** | **r** | ***p* value** | **r** | **95 % CIs** | ***p* value** |
| cUHDRS | 41 | 0.22 | 0.17 | 0.23 | -0.09, 0.52 | 0.16 | 0.16 | 0.32 | 0.12 | -0.14, 0.49 | 0.43 |
| DBS | 41 | -0.09 | 0.57 | -0.15 | -0.39, 0.11 | 0.26 | 0.07 | 0.66 | 0.03 | -0.24, 0.25 | 0.80 |
| TFC | 41 | 0.18 | 0.27 | 0.19 | -0.15, 0.47 | 0.24 | 0.11 | 0.49 | 0.09 | -0.23, 0.41 | 0.59 |
| TMS | 41 | -0.21 | 0.19 | -0.20 | -0.50, 0.12 | 0.22 | -0.15 | 0.36 | -0.09 | -0.46, 0.18 | 0.58 |
| SDMT | 41 | 0.22 | 0.16 | 0.24 | -0.07, 0.52 | 0.13 | 0.17 | 0.29 | 0.15 | -0.11, 0.49 | 0.33 |
| SCN | 41 | 0.35 | **0.03** | 0.36 | 0.09, 0.62 | **0.01** | 0.32 | **0.04** | 0.30 | 0.07, 0.61 | **0.03** |
| VFC | 41 | 0.31 | 0.05 | 0.29 | 0.00, 0.56 | **0.04** | 0.28 | 0.07 | 0.21 | -0.04, 0.53 | 0.15 |
| SWR | 41 | 0.20 | 0.21 | 0.21 | -0.08, 0.49 | 0.16 | 0.13 | 0.40 | 0.10 | -0.10, 0.45 | 0.46 |
| Whole brain | 40 | 0.27 | 0.10 | 0.21 | -0.08, 0.44 | 0.14 | 0.13 | 0.41 | 0.08 | -0.18, 0.36 | 0.59 |
| Caudate | 38 | 0.36 | **0.03** | 0.34 | 0.06, 0.61 | **0.02** | 0.35 | **0.03** | 0.29 | 0.03, 0.59 | **0.04** |
| White matter | 41 | 0.10 | 0.54 | 0.00 | -0.27, 0.29 | 0.99 | 0.07 | 0.68 | -0.04 | -0.33, 0.23 | 0.77 |
| Grey matter | 41 | 0.41 | **0.01** | 0.33 | -0.03, 0.56 | **0.03** | 0.39 | **0.01** | 0.27 | -0.14, 0.58 | 0.16 |
| CSF NfL (log) | 41 | -0.15 | 0.34 | -0.19 | -0.43, 0.04 | 0.12 | -0.07 | 0.65 | -0.06 | -0.29, 0.18 | 0.60 |
| CSF Tau (log) | 41 | -0.03 | 0.87 | -0.07 | -0.42, 0.24 | 0.66 | 0.00 | 0.99 | -0.04 | -0.35, 0.26 | 0.77 |
| CSF mHTT | 41 | -0.11 | 0.48 | -0.11 | -0.36, 0.16 | 0.42 | -0.03 | 0.87 | 0.04 | -0.32, 0.34 | 0.80 |
| Plasma NfL (log) | 41 | -0.25 | 0.12 | -0.25 | -0.57, 0.08 | 0.13 | -0.21 | 0.20 | -0.15 | -0.52, 0.09 | 0.34 |
| Plasma Tau (log) | 41 | -0.28 | 0.07 | -0.28 | -0.53, -0.05 | **0.02** | -0.26 | 0.09 | -0.27 | -0.50, -0.04 | **0.03** |

| **Follow-Up** | | **Age-adjusted** | | | | | **Age and CAG-adjusted** | | | | |
|---|---|---|---|---|---|---|---|---|---|---|---|
| **tNAA** | | **Inverse weighted** | | **Bootstrapped** | | | **Inverse weighted** | | **Bootstrapped** | | |
| **Measures** | **n** | **r** | ***p* value** | **r** | **95 % CIs** | ***p* value** | **r** | ***p* value** | **r** | **95 % CIs** | ***p* value** |
| cUHDRS | 35 | 0.16 | 0.35 | 0.20 | -0.07, 0.45 | 0.13 | 0.36 | **0.03** | 0.40 | 0.13, 0.60 | **0.001** |
| DBS | 36 | 0.11 | 0.53 | 0.07 | -0.19, 0.35 | 0.60 | 0.03 | 0.85 | -0.01 | -0.26, 0.26 | 0.97 |
| TFC | 36 | 0.04 | 0.82 | 0.09 | -0.22, 0.36 | 0.55 | 0.14 | 0.43 | 0.20 | -0.09, 0.45 | 0.15 |
| TMS | 36 | -0.16 | 0.36 | -0.20 | -0.44, 0.07 | 0.12 | -0.31 | 0.07 | -0.35 | -0.59, -0.07 | **0.007** |
| SDMT | 35 | 0.21 | 0.22 | 0.25 | -0.06, 0.53 | 0.09 | 0.38 | **0.03** | 0.41 | 0.14, 0.65 | **0.002** |
| SCN | 35 | 0.18 | 0.30 | 0.20 | -0.08, 0.45 | 0.15 | 0.37 | **0.03** | 0.39 | 0.10, 0.61 | **0.002** |
| VFC | 35 | 0.04 | 0.81 | 0.05 | -0.21, 0.30 | 0.69 | 0.15 | 0.38 | 0.16 | -0.16, 0.45 | 0.28 |
| SWR | 35 | 0.21 | 0.23 | 0.22 | -0.05, 0.44 | 0.08 | 0.38 | **0.02** | 0.39 | 0.15, 0.59 | **<0.001** |
| Whole brain | 34 | 0.09 | 0.60 | 0.10 | -0.21, 0.35 | 0.49 | 0.16 | 0.37 | 0.17 | -0.10, 0.44 | 0.21 |
| Caudate | 34 | 0.18 | 0.30 | 0.17 | -0.11, 0.41 | 0.19 | 0.32 | 0.06 | 0.32 | 0.06, 0.54 | **0.009** |
| White matter | 33 | -0.06 | 0.76 | -0.04 | -0.29, 0.22 | 0.79 | -0.02 | 0.90 | 0.00 | -0.25, 0.26 | 0.99 |
| Grey matter | 33 | 0.17 | 0.35 | 0.18 | -0.15, 0.48 | 0.26 | 0.24 | 0.18 | 0.27 | -0.05, 0.58 | 0.11 |
| CSF NfL (log) | 35 | -0.04 | 0.83 | -0.04 | -0.33, 0.25 | 0.78 | -0.16 | 0.35 | -0.18 | -0.45, 0.18 | 0.26 |
| CSF Tau (log) | 35 | 0.16 | 0.36 | 0.15 | -0.11, 0.39 | 0.25 | 0.13 | 0.45 | 0.12 | -0.13, 0.39 | 0.36 |
| CSF mHTT | 34 | 0.00 | 0.99 | -0.03 | -0.31, 0.27 | 0.84 | -0.08 | 0.64 | -0.10 | -0.40, 0.20 | 0.53 |
| Plasma NfL (log) | 36 | -0.10 | 0.54 | -0.10 | -0.38, 0.17 | 0.48 | -0.26 | 0.13 | -0.26 | -0.52, 0.10 | 0.09 |
| Plasma Tau (log) | 36 | 0.05 | 0.77 | 0.03 | -0.31, 0.38 | 0.87 | 0.03 | 0.87 | 0.01 | -0.35, 0.34 | 0.95 |

| Baseline | | Age-adjusted | | | | | Age and CAG-adjusted | | | | |
|---|---|---|---|---|---|---|---|---|---|---|---|
| **tCre** | | **Inverse weighted** | | **Bootstrapped** | | | **Inverse weighted** | | **Bootstrapped** | | |
| **Measures** | **n** | **r** | ***p* value** | **r** | **95 % CIs** | ***p* value** | **r** | ***p* value** | **r** | **95 % CIs** | ***p* value** |
| cUHDRS | 41 | 0.14 | 0.40 | 0.13 | -0.19, 0.42 | 0.39 | 0.26 | 0.10 | 0.25 | -0.04, 0.53 | 0.09 |
| DBS | 41 | 0.13 | 0.41 | 0.11 | -0.11, 0.31 | 0.30 | 0.06 | 0.71 | 0.04 | -0.19, 0.26 | 0.74 |
| TFC | 41 | -0.01 | 0.95 | 0.03 | -0.23, 0.33 | 0.85 | 0.05 | 0.77 | 0.10 | -0.19, 0.43 | 0.54 |
| TMS | 41 | -0.09 | 0.57 | -0.07 | -0.36, 0.25 | 0.66 | -0.19 | 0.24 | -0.16 | -0.45, 0.15 | 0.28 |
| SDMT | 41 | 0.23 | 0.16 | 0.23 | -0.10, 0.50 | 0.15 | 0.34 | **0.03** | 0.34 | 0.03, 0.59 | **0.02** |
| SCN | 41 | 0.28 | 0.08 | 0.27 | -0.07, 0.53 | 0.09 | 0.41 | **0.01** | 0.40 | 0.07, 0.63 | **0.004** |
| VFC | 41 | 0.13 | 0.41 | 0.11 | -0.17, 0.39 | 0.47 | 0.27 | 0.08 | 0.24 | -0.05, 0.48 | 0.08 |
| SWR | 41 | 0.15 | 0.34 | 0.14 | -0.20, 0.41 | 0.36 | 0.28 | 0.08 | 0.25 | -0.06, 0.50 | 0.07 |
| Whole brain | 40 | 0.22 | 0.18 | 0.14 | -0.16, 0.41 | 0.32 | 0.26 | 0.10 | 0.19 | -0.07, 0.43 | 0.14 |
| Caudate | 38 | 0.38 | **0.02** | 0.31 | 0.01, 0.57 | **0.03** | 0.50 | **0.001** | 0.42 | 0.11, 0.63 | **0.001** |
| White matter | 41 | 0.14 | 0.38 | 0.10 | -0.17, 0.33 | 0.46 | 0.16 | 0.31 | 0.11 | -0.14, 0.34 | 0.36 |
| Grey matter | 41 | 0.27 | 0.09 | 0.17 | -0.16, 0.43 | 0.28 | 0.36 | **0.02** | 0.25 | -0.11, 0.50 | 0.10 |
| CSF NfL (log) | 41 | -0.16 | 0.33 | -0.09 | -0.39, 0.20 | 0.54 | -0.29 | 0.07 | -0.21 | -0.47, 0.19 | 0.20 |
| CSF Tau (log) | 41 | -0.05 | 0.75 | -0.01 | -0.36, 0.51 | 0.98 | -0.07 | 0.68 | -0.02 | -0.37, 0.52 | 0.93 |
| CSF mHTT | 41 | -0.06 | 0.72 | 0.03 | -0.30, 0.40 | 0.88 | -0.15 | 0.37 | -0.04 | -0.40, 0.45 | 0.84 |
| Plasma NfL (log) | 41 | -0.23 | 0.15 | -0.19 | -0.47, 0.10 | 0.21 | -0.40 | **0.01** | -0.34 | -0.57, -0.09 | **0.007** |
| Plasma Tau (log) | 41 | -0.22 | 0.18 | -0.27 | -0.51, 0.03 | 0.06 | -0.22 | 0.16 | -0.28 | -0.52, 0.01 | 0.05 |

| **Follow-Up** | | **Age-adjusted** | | | | | **Age and CAG-adjusted** | | | | |
|---|---|---|---|---|---|---|---|---|---|---|---|
| **tCre** | | **Inverse weighted** | | **Bootstrapped** | | | **Inverse weighted** | | **Bootstrapped** | | |
| **Measures** | **n** | **r** | ***p* value** | **r** | **95 % CIs** | ***p* value** | **r** | ***p* value** | **r** | **95 % CIs** | ***p* value** |
| cUHDRS | 35 | 0.50 | **0.002** | 0.45 | 0.07, 0.67 | **0.003** | 0.43 | **0.01** | 0.41 | 0.12, 0.63 | **0.001** |
| DBS | 36 | -0.32 | 0.06 | -0.26 | -0.60, 0.06 | 0.14 | -0.12 | 0.48 | -0.05 | -0.42, 0.29 | 0.81 |
| TFC | 36 | 0.32 | 0.06 | 0.32 | -0.02, 0.62 | 0.05 | 0.16 | 0.37 | 0.22 | -0.06, 0.51 | 0.13 |
| TMS | 36 | -0.54 | **<0.001** | -0.48 | -0.73, -0.17 | **0.001** | -0.48 | **0.003** | -0.44 | -0.68, -0.16 | **0.001** |
| SDMT | 35 | 0.48 | **0.004** | 0.42 | 0.10, 0.62 | **0.001** | 0.39 | **0.02** | 0.36 | 0.10, 0.57 | **0.002** |
| SCN | 35 | 0.52 | **0.001** | 0.47 | 0.13, 0.68 | **0.001** | 0.46 | **0.005** | 0.44 | 0.17, 0.66 | **0.001** |
| VFC | 35 | 0.36 | **0.03** | 0.30 | -0.01, 0.53 | **0.03** | 0.22 | 0.19 | 0.21 | -0.12, 0.48 | 0.18 |
| SWR | 35 | 0.48 | **0.003** | 0.43 | 0.01, 0.66 | **0.005** | 0.40 | **0.02** | 0.38 | 0.02, 0.63 | **0.01** |
| Whole brain | 34 | 0.34 | 0.05 | 0.31 | -0.00, 0.70 | 0.09 | 0.35 | 0.05 | 0.30 | -0.04, 0.68 | 0.11 |
| Caudate | 34 | 0.47 | **0.006** | 0.43 | 0.16, 0.72 | **0.003** | 0.54 | **<0.001** | 0.49 | 0.28, 0.75 | **<0.001** |
| White matter | 33 | 0.24 | 0.19 | 0.19 | -0.26, 0.55 | 0.37 | 0.22 | 0.22 | 0.17 | -0.31, 0.54 | 0.43 |
| Grey matter | 33 | 0.40 | **0.02** | 0.38 | 0.11, 0.64 | **0.007** | 0.40 | **0.02** | 0.38 | 0.08, 0.63 | **0.007** |
| CSF NfL (log) | 35 | -0.59 | **<0.001** | -0.51 | -0.72, -0.17 | **<0.001** | -0.58 | **<0.001** | -0.51 | -0.71, -0.18 | **<0.001** |
| CSF Tau (log) | 35 | -0.45 | **0.007** | -0.37 | -0.65, -0.06 | **0.02** | -0.35 | **0.04** | -0.30 | -0.58, -0.01 | **0.04** |
| CSF mHTT | 34 | -0.53 | **0.001** | -0.48 | -0.75, -0.16 | **0.002** | -0.48 | **0.004** | -0.47 | -0.74, -0.17 | **0.002** |
| Plasma NfL (log) | 36 | -0.58 | **<0.001** | -0.51 | -0.74, -0.17 | **0.001** | -0.57 | **0.003** | -0.52 | -0.75, -0.27 | **<0.001** |
| Plasma Tau (log) | 36 | -0.09 | 0.59 | -0.10 | -0.40, 0.17 | 0.50 | -0.01 | 0.97 | -0.05 | -0.32, 0.21 | 0.74 |

| **Baseline** | | **Age and Gender-adjusted** | | | | | **Age, Gender and CAG-adjusted** | | | | |
|---|---|---|---|---|---|---|---|---|---|---|---|
| **tCho** | | **Inverse weighted** | | **Bootstrapped** | | | **Inverse weighted** | | **Bootstrapped** | | |
| **Measures** | **n** | **r** | ***p* value** | **r** | **95 % CIs** | ***p* value** | **r** | ***p* value** | **r** | **95 % CIs** | ***p* value** |
| cUHDRS | 41 | -0.07 | 0.66 | -0.05 | -0.39, 0.29 | 0.79 | 0.17 | 0.30 | 0.18 | -0.12, 0.46 | 0.22 |
| DBS | 41 | 0.30 | 0.06 | 0.30 | 0.05, 0.50 | **0.007** | 0.08 | 0.60 | 0.07 | -0.21, 0.30 | 0.56 |
| TFC | 41 | -0.15 | 0.34 | -0.10 | -0.42, 0.26 | 0.55 | 0.02 | 0.92 | 0.08 | -0.22, 0.41 | 0.63 |
| TMS | 41 | 0.12 | 0.47 | 0.09 | -0.25, 0.40 | 0.56 | -0.09 | 0.57 | -0.11 | -0.39, 0.20 | 0.47 |
| SDMT | 41 | 0.05 | 0.78 | 0.07 | -0.30, 0.38 | 0.71 | 0.27 | 0.08 | 0.28 | -0.03, 0.52 | **0.04** |
| SCN | 41 | -0.02 | 0.92 | -0.01 | -0.33, 0.29 | 0.93 | 0.20 | 0.21 | 0.20 | -0.07, 0.44 | 0.13 |
| VFC | 41 | 0.10 | 0.54 | 0.10 | -0.22, 0.37 | 0.50 | 0.45 | **0.004** | 0.43 | 0.20, 0.60 | **<0.001** |
| SWR | 41 | -0.08 | 0.64 | -0.07 | -0.37, 0.25 | 0.64 | 0.15 | 0.34 | 0.14 | -0.13, 0.42 | 0.30 |
| Whole brain | 40 | 0.11 | 0.51 | 0.06 | -0.24, 0.37 | 0.72 | 0.26 | 0.11 | 0.20 | -0.05, 0.53 | 0.16 |
| Caudate | 38 | 0.02 | 0.89 | -0.03 | -0.34, 0.31 | 0.86 | 0.20 | 0.23 | 0.13 | -0.20, 0.46 | 0.45 |
| White matter | 41 | 0.23 | 0.15 | 0.20 | -0.11, 0.46 | 0.17 | 0.32 | **0.04** | 0.28 | -0.00, 0.51 | **0.04** |
| Grey matter | 41 | 0.18 | 0.27 | 0.08 | -0.32, 0.40 | 0.68 | 0.38 | **0.01** | 0.26 | -0.16, 0.57 | 0.15 |
| CSF NfL (log) | 41 | 0.26 | 0.10 | 0.30 | 0.01, 0.52 | **0.02** | 0.10 | 0.54 | 0.15 | -0.08, 0.41 | 0.25 |
| CSF Tau (log) | 41 | -0.04 | 0.83 | -0.01 | -0.29, 0.36 | 0.96 | -0.09 | 0.56 | -0.05 | -0.31, 0.36 | 0.74 |
| CSF mHTT | 41 | 0.01 | 0.95 | 0.07 | -0.27, 0.39 | 0.66 | -0.24 | 0.14 | -0.15 | -0.47, 0.24 | 0.42 |
| Plasma NfL (log) | 41 | 0.15 | 0.34 | 0.15 | -0.17, 0.42 | 0.32 | -0.06 | 0.70 | -0.05 | -0.36, 0.25 | **0.73** |
| Plasma Tau (log) | 41 | -0.17 | 0.29 | -0.21 | -0.45, 0.05 | 0.09 | -0.20 | 0.21 | -0.26 | -0.47, 0.01 | **0.04** |

| **Follow-Up** | | **Age and Gender-adjusted** | | | | | **Age, Gender and CAG-adjusted** | | | | |
|---|---|---|---|---|---|---|---|---|---|---|---|
| **tCho** | | **Inverse weighted** | | **Bootstrapped** | | | **Inverse weighted** | | **Bootstrapped** | | |
| **Measures** | **n** | **r** | ***p* value** | **r** | **95 % CIs** | ***p* value** | **r** | ***p* value** | **r** | **95 % CIs** | ***p* value** |
| cUHDRS | 35 | 0.20 | 0.24 | 0.11 | -0.27, 0.45 | 0.53 | 0.15 | 0.40 | 0.11 | -0.24, 0.41 | 0.49 |
| DBS | 36 | -0.17 | 0.32 | -0.06 | -0.51, 0.27 | 0.78 | -0.12 | 0.48 | -0.01 | -0.47, 0.32 | 0.98 |
| TFC | 36 | -0.06 | 0.72 | -0.11 | -0.43, 0.25 | 0.53 | -0.21 | 0.21 | -0.19 | -0.45, 0.09 | 0.18 |
| TMS | 36 | -0.25 | 0.15 | -0.15 | -0.45, 0.23 | 0.38 | -0.21 | 0.22 | -0.15 | -0.41, 0.21 | 0.32 |
| SDMT | 35 | 0.28 | 0.10 | 0.19 | -0.20, 0.49 | 0.28 | 0.24 | 0.15 | 0.20 | -0.14, 0.50 | 0.21 |
| SCN | 35 | 0.27 | 0.12 | 0.17 | -0.17, 0.49 | 0.31 | 0.24 | 0.17 | 0.20 | -0.10, 0.53 | 0.23 |
| VFC | 35 | 0.16 | 0.35 | 0.10 | -0.18, 0.41 | 0.47 | 0.09 | 0.60 | 0.10 | -0.20, 0.42 | 0.53 |
| SWR | 35 | 0.25 | 0.14 | 0.18 | -0.19, 0.48 | 0.30 | 0.21 | 0.22 | 0.19 | -0.09, 0.48 | 0.20 |
| Whole brain | 34 | 0.11 | 0.54 | 0.08 | -0.30, 0.38 | 0.66 | 0.19 | 0.28 | 0.16 | -0.24, 0.46 | 0.38 |
| Caudate | 34 | 0.07 | 0.71 | 0.03 | -0.28, 0.44 | 0.86 | 0.19 | 0.27 | 0.15 | -0.16, 0.54 | 0.38 |
| White matter | 33 | 0.16 | 0.39 | 0.13 | -0.21, 0.47 | 0.46 | 0.21 | 0.23 | 0.18 | -0.16, 0.53 | 0.30 |
| Grey matter | 33 | 0.09 | 0.62 | 0.06 | -0.31, 0.37 | 0.72 | 0.17 | 0.34 | 0.14 | -0.24, 0.43 | 0.42 |
| CSF NfL (log) | 35 | -0.42 | **0.01** | -0.33 | -0.61, 0.05 | 0.05 | -0.48 | **0.003** | -0.43 | -0.66, -0.11 | **0.002** |
| CSF Tau (log) | 35 | -0.41 | **0.02** | -0.40 | -0.65, -0.13 | **0.003** | -0.38 | **0.02** | -0.40 | -0.61, -0.12 | **0.001** |
| CSF mHTT | 34 | -0.39 | **0.02** | -0.34 | -0.64, 0.01 | 0.05 | -0.40 | **0.02** | -0.39 | -0.62, 0.08 | **0.006** |
| Plasma NfL (log) | 36 | -0.36 | **0.03** | -0.24 | -0.59, 0.13 | 0.20 | -0.40 | **0.02** | -0.30 | -0.60, 0.01 | 0.06 |
| Plasma Tau (log) | 36 | 0.06 | 0.71 | 0.09 | -0.23, 0.36 | 0.55 | 0.10 | 0.54 | 0.11 | -0.18, 0.37 | 0.44 |

| **Baseline** | | **Age-adjusted** | | | | | **Age and CAG-adjusted** | | | | |
|---|---|---|---|---|---|---|---|---|---|---|---|
| **MI** | | **Inverse weighted** | | **Bootstrapped** | | | **Inverse weighted** | | **Bootstrapped** | | |
| **Measures** | **n** | **r** | ***p* value** | **r** | **95 % CIs** | ***p* value** | **r** | ***p* value** | **r** | **95 % CIs** | ***p* value** |
| cUHDRS | 41 | -0.35 | **0.02** | -0.40 | -0.61, -0.13 | **0.001** | -0.15 | 0.34 | -0.22 | -0.53, 0.06 | 0.14 |
| DBS | 41 | 0.26 | 0.10 | 0.37 | 0.03, 0.58 | **0.005** | -0.04 | 0.79 | 0.05 | -0.26, 0.31 | 0.73 |
| TFC | 41 | -0.22 | 0.17 | -0.26 | -0.52, -0.01 | 0.05 | -0.01 | 0.97 | -0.06 | -0.37, 0.21 | 0.70 |
| TMS | 41 | 0.34 | **0.03** | 0.44 | 0.12, 0.66 | **0.001** | 0.15 | 0.36 | 0.28 | -0.11, 0.58 | 0.11 |
| SDMT | 41 | -0.32 | **0.04** | -0.36 | -0.59, -0.02 | **0.009** | -0.14 | 0.39 | -0.18 | -0.47, 0.09 | 0.20 |
| SCN | 41 | -0.36 | **0.02** | -0.42 | -0.64, -0.17 | **<0.001** | -0.19 | 0.24 | -0.26 | -0.58, 0.04 | 0.10 |
| VFC | 41 | -0.33 | **0.04** | -0.39 | -0.62, -0.09 | **0.004** | -0.11 | 0.50 | -0.19 | -0.49, 0.11 | 0.23 |
| SWR | 41 | -0.40 | **0.01** | -0.42 | -0.64, -0.20 | **<0.001** | -0.22 | 0.16 | -0.24 | -0.56, 0.00 | 0.08 |
| Whole brain | 40 | -0.26 | 0.11 | -0.37 | -0.57, -0.10 | **0.002** | -0.12 | 0.45 | -0.23 | -0.49, 0.10 | 0.11 |
| Caudate | 38 | -0.44 | **0.005** | -0.47 | -0.68, -0.22 | **<0.001** | -0.32 | **0.05** | -0.34 | -0.62, -0.07 | **0.02** |
| White matter | 41 | -0.06 | 0.72 | -0.16 | -0.40, 0.16 | 0.26 | -0.01 | 0.97 | -0.09 | -0.35, 0.23 | 0.54 |
| Grey matter | 41 | -0.33 | **0.03** | -0.46 | -0.64, -0.20 | **<0.001** | -0.19 | 0.23 | -0.33 | -0.55, -0.05 | **0.008** |
| CSF NfL (log) | 41 | 0.35 | **0.03** | 0.35 | 0.12, 0.57 | **0.003** | 0.13 | 0.42 | 0.13 | -0.15, 0.42 | 0.37 |
| CSF Tau (log) | 41 | 0.08 | 0.61 | 0.05 | -0.35, 0.45 | 0.80 | -0.00 | 0.98 | -0.00 | -0.37, 0.36 | 0.98 |
| CSF mHTT | 41 | 0.13 | 0.42 | 0.22 | -0.23, 0.48 | 0.21 | -0.16 | 0.32 | -0.04 | -0.46, 0.26 | 0.78 |
| Plasma NfL (log) | 41 | 0.52 | **<0.001** | 0.52 | 0.21, 0.73 | **<0.001** | 0.39 | **0.01** | 0.39 | 0.04, 0.67 | **0.02** |
| Plasma Tau (log) | 41 | 0.04 | 0.79 | -0.05 | -0.40, 0.33 | 0.79 | 0.03 | 0.85 | -0.09 | -0.45, 0.31 | 0.64 |

| Follow-Up | | Age-adjusted | | | | | Age and CAG-adjusted | | | | |
|---|---|---|---|---|---|---|---|---|---|---|---|
| MI | | Inverse weighted | | Bootstrapped | | | Inverse weighted | | Bootstrapped | | |
| **Measures** | **n** | **r** | ***p* value** | **r** | **95 % CIs** | ***p* value** | **r** | ***p* value** | **r** | **95 % CIs** | ***p* value** |
| cUHDRS | 35 | -0.17 | 0.32 | -0.23 | -0.48, 0.07 | 0.10 | -0.23 | 0.19 | -0.24 | -0.50, -0.02 | 0.06 |
| DBS | 35 | -0.06 | 0.74 | 0.03 | -0.23, 0.24 | 0.78 | -0.12 | 0.50 | -0.03 | -0.28, 0.22 | 0.82 |
| TFC | 36 | -0.16 | 0.36 | -0.22 | -0.50, 0.16 | 0.21 | -0.19 | 0.27 | -0.22 | -0.48, 0.15 | 0.16 |
| TMS | 36 | 0.16 | 0.36 | 0.18 | -0.08, 0.41 | 0.15 | 0.20 | 0.25 | 0.18 | -0.10, 0.44 | 0.21 |
| SDMT | 35 | -0.09 | 0.62 | -0.15 | -0.41, 0.13 | 0.29 | -0.09 | 0.60 | -0.13 | -0.38, 0.10 | 0.32 |
| SCN | 35 | -0.22 | 0.20 | -0.27 | -0.54, -0.02 | **0.04** | -0.30 | 0.08 | -0.29 | -0.49, -0.08 | **0.006** |
| VFC | 35 | -0.13 | 0.47 | -0.19 | -0.41, 0.11 | 0.15 | -0.15 | 0.40 | -0.18 | -0.40, 0.04 | 0.12 |
| SWR | 35 | -0.23 | 0.18 | -0.27 | -0.48, -0.02 | **0.02** | -0.30 | 0.08 | -0.29 | -0.48, -0.07 | **0.008** |
| Whole brain | 34 | -0.25 | 0.15 | -0.31 | -0.55, 0.01 | **0.03** | -0.26 | 0.14 | -0.30 | -0.51, 0.05 | **0.02** |
| Caudate | 34 | -0.35 | **0.04** | -0.39 | -0.67, -0.14 | **0.006** | -0.43 | **0.01** | -0.42 | -0.62, -0.18 | **<0.001** |
| White matter | 33 | 0.04 | 0.83 | -0.06 | -0.33, 0.26 | 0.67 | 0.06 | 0.75 | -0.03 | -0.35, 0.23 | 0.83 |
| Grey matter | 33 | -0.42 | **0.02** | -0.46 | -0.67, -0.15 | **<0.001** | -0.44 | **0.01** | -0.47 | -0.66, 0.16 | **<0.001** |
| CSF NfL (log) | 35 | 0.17 | 0.34 | 0.17 | -0.10, 0.52 | 0.28 | 0.14 | 0.41 | 0.12 | -0.12, 0.36 | 0.34 |
| CSF Tau (log) | 35 | 0.17 | 0.32 | 0.14 | -0.16, 0.40 | 0.32 | 0.14 | 0.41 | 0.10 | -0.18, 0.32 | 0.44 |
| CSF mHTT | 34 | 0.10 | 0.59 | 0.13 | -0.15, 0.45 | 0.39 | 0.00 | 0.99 | 0.02 | -0.23, 0.24 | 0.89 |
| Plasma NfL (log) | 36 | 0.14 | 0.42 | 0.18 | -0.10, 0.53 | 0.26 | 0.19 | 0.27 | 0.20 | -0.13, 0.46 | 0.18 |
| Plasma Tau (log) | 36 | 0.21 | 0.21 | 0.19 | -0.21, 0.39 | 0.14 | 0.21 | 0.21 | 0.18 | -0.17, 0.42 | 0.19 |

| **Baseline** | | **Age-adjusted** | | | | | **Age and CAG-adjusted** | | | | |
|---|---|---|---|---|---|---|---|---|---|---|---|
| **GSH** | | **Inverse weighted** | | **Bootstrapped** | | | **Inverse weighted** | | **Bootstrapped** | | |
| **Measures** | **n** | **r** | ***p* value** | **r** | **95 % CIs** | ***p* value** | **r** | ***p* value** | **r** | **95 % CIs** | ***p* value** |
| cUHDRS | 41 | 0.08 | 0.60 | 0.15 | -0.13, 0.42 | 0.27 | 0.14 | 0.37 | 0.21 | -0.09, 0.48 | 0.15 |
| DBS | 41 | 0.04 | 0.82 | 0.02 | -0.29, 0.35 | 0.91 | 0.04 | 0.83 | 0.02 | -0.29, 0.35 | 0.92 |
| TFC | 41 | 0.09 | 0.58 | 0.16 | -0.09, 0.43 | 0.21 | 0.13 | 0.40 | 0.20 | -0.08, 0.48 | 0.16 |
| TMS | 41 | -0.00 | 0.99 | -0.05 | -0.30, 0.24 | 0.72 | -0.03 | 0.86 | -0.07 | -0.36, 0.22 | 0.66 |
| SDMT | 41 | 0.11 | 0.49 | 0.19 | -0.12, 0.48 | 0.22 | 0.16 | 0.31 | 0.23 | -0.11, 0.49 | 0.13 |
| SCN | 41 | 0.17 | 0.29 | 0.22 | -0.02, 0.46 | 0.08 | 0.24 | 0.13 | 0.27 | 0.02, 0.48 | **0.02** |
| VFC | 41 | 0.09 | 0.58 | 0.12 | -0.18, 0.40 | 0.44 | 0.16 | 0.33 | 0.16 | -0.13, 0.42 | 0.25 |
| SWR | 41 | 0.10 | 0.52 | 0.17 | -0.08, 0.42 | 0.20 | 0.17 | 0.30 | 0.22 | -0.04, 0.48 | 0.09 |
| Whole brain | 41 | 0.22 | 0.17 | 0.23 | -0.08, 0.51 | 0.12 | 0.24 | 0.14 | 0.25 | -0.04, 0.52 | 0.08 |
| Caudate | 38 | 0.30 | 0.06 | 0.32 | 0.03, 0.56 | **0.02** | 0.36 | **0.03** | 0.35 | 0.07, 0.58 | **0.007** |
| White matter | 41 | 0.16 | 0.33 | 0.18 | -0.08, 0.43 | 0.18 | 0.17 | 0.30 | 0.18 | -0.06, 0.45 | 0.17 |
| Grey matter | 41 | 0.24 | 0.12 | 0.19 | -0.15, 0.46 | 0.23 | 0.29 | 0.06 | 0.22 | -0.13, 0.48 | 0.14 |
| CSF NfL (log) | 41 | -0.07 | 0.65 | -0.05 | -0.37, 0.24 | 0.77 | -0.13 | 0.43 | -0.07 | -0.42, 0.21 | 0.68 |
| CSF Tau (log) | 41 | -0.05 | 0.74 | -0.02 | -0.38, 0.32 | 0.92 | -0.06 | 0.72 | -0.02 | -0.37, 0.32 | 0.92 |
| CSF mHTT | 41 | 0.06 | 0.72 | 0.10 | -0.20, 0.40 | 0.51 | 0.05 | 0.76 | 0.13 | -0.18, 0.46 | 0.41 |
| Plasma NfL (log) | 41 | -0.15 | 0.35 | -0.14 | -0.43, 0.14 | 0.35 | -0.14 | 0.14 | -0.19 | -0.48, 0.09 | 0.17 |
| Plasma Tau (log) | 41 | -0.22 | 0.17 | -0.30 | -0.54, 0.06 | **0.04** | -0.22 | 0.16 | -0.31 | -0.54, 0.07 | **0.04** |

| Follow Up | | Age-adjusted | | | | | Age and CAG-adjusted | | | | |
|---|---|---|---|---|---|---|---|---|---|---|---|
| **GSH** | | **Inverse weighted** | | **Bootstrapped** | | | **Inverse weighted** | | **Bootstrapped** | | |
| **Measures** | **n** | **r** | ***p* value** | **r** | **95 % CIs** | ***p* value** | **r** | ***p* value** | **r** | **95 % CIs** | ***p* value** |
| cUHDRS | 35 | 0.23 | 0.19 | 0.23 | -0.14, 0.52 | 0.17 | 0.04 | 0.81 | 0.08 | -0.23, 0.35 | 0.61 |
| DBS | 36 | -0.34 | **0.04** | -0.27 | -0.57, 0.01 | 0.08 | -0.16 | 0.34 | -0.07 | -0.42, 0.23 | 0.69 |
| TFC | 36 | 0.21 | 0.22 | 0.18 | -0.16, 0.48 | 0.26 | 0.07 | 0.69 | 0.04 | -0.23, 0.36 | 0.79 |
| TMS | 36 | -0.26 | 0.12 | -0.28 | -0.58, 0.04 | 0.09 | -0.14 | 0.43 | -0.17 | -0.41, 0.11 | 0.22 |
| SDMT | 35 | 0.25 | 0.15 | 0.26 | -0.09, 0.54 | 0.12 | 0.10 | 0.57 | 0.12 | -0.17, 0.43 | 0.42 |
| SCN | 35 | 0.12 | 0.49 | 0.15 | -0.22, 0.49 | 0.42 | -0.12 | 0.51 | -0.05 | -0.37, 0.30 | 0.79 |
| VFC | 35 | 0.19 | 0.27 | 0.19 | -0.22, 0.55 | 0.34 | 0.01 | 0.94 | 0.03 | -0.37, 0.39 | 0.88 |
| SWR | 35 | 0.07 | 0.68 | 0.11 | -0.28, 0.43 | 0.54 | -0.17 | 0.33 | -0.09 | -0.39, 0.25 | 0.60 |
| Whole brain | 34 | 0.03 | 0.86 | 0.19 | -0.08, 0.50 | 0.22 | -0.01 | 0.94 | 0.15 | -0.14, 0.46 | 0.31 |
| Caudate | 34 | -0.01 | 0.93 | 0.11 | -0.24, 0.40 | 0.37 | -0.11 | 0.55 | 0.05 | -0.30, 0.31 | 0.76 |
| White matter | 33 | 0.26 | 0.15 | 0.22 | -0.16, 0.52 | 0.22 | 0.24 | 0.18 | 0.19 | -0.13, 0.49 | 0.25 |
| Grey matter | 33 | -0.07 | 0.71 | 0.17 | -0.18, 0.53 | 0.35 | -0.12 | 0.52 | 0.12 | -0.25, 0.49 | 0.50 |
| CSF NfL (log) | 35 | -0.26 | 0.13 | -0.28 | -0.59, 0.08 | 0.11 | -0.20 | 0.25 | -0.18 | -0.45, 0.13 | 0.21 |
| CSF Tau (log) | 35 | -0.19 | 0.27 | -0.19 | -0.51, 0.15 | 0.27 | -0.11 | 0.53 | -0.11 | -0.40, 0.24 | 0.48 |
| CSF mHTT | 34 | -0.31 | 0.08 | -0.35 | -0.66, 0.04 | 0.05 | -0.25 | 0.16 | -0.28 | -0.57, 0.04 | 0.08 |
| Plasma NfL (log) | 36 | -0.24 | 0.16 | -0.26 | -0.52, 0.03 | 0.08 | -0.10 | 0.58 | -0.12 | -0.39, 0.14 | 0.37 |
| Plasma Tau (log) | 36 | -0.18 | 0.30 | -0.20 | -0.47, 0.08 | 0.14 | -0.12 | 0.48 | -0.16 | -0.42, 0.11 | 0.23 |

| **Baseline** | | **Age-adjusted** | | | | | **Age and CAG-adjusted** | | | | |
|---|---|---|---|---|---|---|---|---|---|---|---|
| **GABA** | | **Inverse weighted** | | **Bootstrapped** | | | **Inverse weighted** | | **Bootstrapped** | | |
| **Measures** | **n** | **r** | ***p* value** | **r** | **95 % CIs** | ***p* value** | **r** | ***p* value** | **r** | **95 % CIs** | ***p* value** |
| cUHDRS | 33 | 0.12 | 0.50 | 0.14 | -0.24, 0.44 | 0.42 | 0.11 | 0.55 | 0.10 | -0.19, 0.41 | 0.49 |
| DBS | 33 | 0.02 | 0.92 | -0.01 | -0.31, 0.32 | 0.93 | 0.09 | 0.62 | 0.06 | -0.25, 0.37 | 0.72 |
| TFC | 33 | 0.01 | 0.97 | 0.05 | -0.24, 0.30 | 0.71 | -0.03 | 0.86 | 0.01 | -0.30, 0.26 | 0.95 |
| TMS | 33 | -0.09 | 0.63 | -0.08 | -0.41, 0.28 | 0.62 | -0.07 | 0.72 | -0.05 | -0.38, 0.28 | 0.78 |
| SDMT | 33 | 0.14 | 0.43 | 0.13 | -0.21, 0.44 | 0.44 | 0.13 | 0.47 | 0.10 | -0.18, 0.39 | 0.47 |
| SCN | 33 | 0.28 | 0.12 | 0.28 | -0.08, 0.54 | 0.07 | 0.28 | 0.11 | 0.27 | -0.01, 0.53 | 0.06 |
| VFC | 33 | 0.18 | 0.32 | 0.18 | -0.15, 0.48 | 0.26 | 0.18 | 0.31 | 0.17 | -0.12, 0.46 | 0.25 |
| SWR | 33 | 0.19 | 0.29 | 0.21 | -0.22, 0.52 | 0.23 | 0.19 | 0.29 | 0.20 | -0.12, 0.53 | 0.23 |
| Whole brain | 32 | 0.02 | 0.90 | 0.04 | -0.24, 0.33 | 0.81 | -0.05 | 0.77 | -0.03 | -0.32, 0.28 | 0.87 |
| Caudate | 32 | 0.07 | 0.71 | 0.07 | -0.22, 0.37 | 0.64 | 0.05 | 0.80 | 0.03 | -0.36, 0.36 | 0.86 |
| White matter | 33 | 0.04 | 0.82 | -0.00 | -0.33, 0.34 | 1.00 | 0.04 | 0.84 | -0.01 | -0.34, 0.31 | 0.93 |
| Grey matter | 33 | 0.28 | 0.11 | 0.24 | -0.05, 0.53 | 0.11 | 0.29 | 0.11 | 0.23 | -0.16, 0.61 | 0.26 |
| CSF NfL (log) | 33 | -0.15 | 0.41 | -0.14 | -0.43, 0.21 | 0.41 | -0.15 | 0.41 | -0.10 | -0.42, 0.21 | 0.51 |
| CSF Tau (log) | 33 | -0.28 | 0.12 | -0.29 | -0.55, -0.03 | **0.04** | -0.28 | 0.12 | -0.29 | -0.53, -0.05 | **0.03** |
| CSF mHTT | 33 | -0.32 | 0.07 | -0.32 | -0.57, -0.03 | **0.02** | -0.39 | **0.02** | -0.38 | -0.59, -0.09 | **0.003** |
| Plasma NfL (log) | 33 | -0.01 | 0.95 | -0.07 | -0.36, 0.24 | 0.67 | 0.02 | 0.90 | -0.02 | -0.32, 0.37 | 0.92 |
| Plasma Tau (log) | 33 | 0.00 | 0.99 | 0.02 | -0.27, 0.34 | 0.88 | 0.00 | 0.99 | 0.02 | -0.28, 0.34 | 0.91 |

| Follow-Up | | Age-adjusted | | | | | Age and CAG-adjusted | | | | |
|---|---|---|---|---|---|---|---|---|---|---|---|
| GABA | | Inverse weighted | | Bootstrapped | | | Inverse weighted | | Bootstrapped | | |
| **Measures** | **n** | **r** | ***p* value** | **r** | **95 % CIs** | ***p* value** | **r** | ***p* value** | **r** | **95 % CIs** | ***p* value** |
| cUHDRS | 29 | 0.10 | 0.60 | 0.10 | -0.24, 0.34 | 0.48 | 0.11 | 0.59 | 0.08 | -0.43, 0.35 | 0.66 |
| DBS | 30 | -0.03 | 0.89 | -0.03 | -0.51, 0.42 | 0.89 | 0.06 | 0.74 | 0.04 | -0.43, 0.48 | 0.85 |
| TFC | 30 | 0.11 | 0.56 | 0.12 | -0.38, 0.40 | 0.51 | 0.08 | 0.69 | 0.08 | -0.35, 0.40 | 0.67 |
| TMS | 30 | -0.17 | 0.37 | -0.17 | -0.39, 0.11 | 0.18 | -0.16 | 0.41 | -0.14 | -0.39, 0.29 | 0.41 |
| SDMT | 29 | 0.10 | 0.59 | 0.05 | -0.21, 0.33 | 0.69 | 0.10 | 0.60 | 0.02 | -0.49, 0.34 | 0.92 |
| SCN | 29 | 0.18 | 0.35 | 0.16 | -0.17, 0.40 | 0.25 | 0.22 | 0.25 | 0.17 | -0.34, 0.41 | 0.33 |
| VFC | 29 | 0.09 | 0.64 | 0.07 | -0.27, 0.31 | 0.64 | 0.09 | 0.66 | 0.03 | -0.49, 0.31 | 0.85 |
| SWR | 29 | 0.06 | 0.76 | 0.08 | -0.29, 0.35 | 0.60 | 0.04 | 0.83 | 0.06 | -0.35, 0.35 | 0.75 |
| Whole brain | 28 | 0.03 | 0.89 | -0.03 | -0.32, 0.26 | 0.84 | -0.05 | 0.79 | -0.09 | -0.36, 0.22 | 0.53 |
| Caudate | 28 | 0.26 | 0.17 | 0.23 | -0.13, 0.48 | 0.11 | 0.19 | 0.32 | 0.19 | -0.18, 0.50 | 0.24 |
| White matter | 27 | -0.02 | 0.93 | -0.07 | -0.41, 0.21 | 0.63 | -0.09 | 0.67 | -0.12 | -0.48, 0.16 | 0.45 |
| Grey matter | 27 | 0.08 | 0.69 | 0.03 | -0.31, 0.27 | 0.81 | 0.01 | 0.96 | -0.02 | -0.27, 0.25 | 0.87 |
| CSF NfL (log) | 29 | -0.10 | 0.60 | -0.17 | -0.49, 0.19 | 0.33 | -0.07 | 0.71 | -0.16 | -0.40, 0.12 | 0.25 |
| CSF Tau (log) | 29 | -0.06 | 0.74 | -0.08 | -0.28, 0.22 | 0.52 | -0.03 | 0.88 | -0.05 | -0.28, 0.20 | 0.71 |
| CSF mHTT | 28 | -0.04 | 0.83 | -0.03 | -0.32, 0.31 | 0.83 | -0.11 | 0.59 | -0.13 | -0.37, 0.09 | 0.27 |
| Plasma NfL (log) | 30 | -0.12 | 0.52 | -0.17 | -0.44, 0.23 | 0.30 | -0.09 | 0.64 | -0.14 | -0.42, 0.18 | 0.35 |
| Plasma Tau (log) | 30 | -0.04 | 0.85 | 0.02 | -0.42, 0.41 | 0.94 | -0.03 | 0.88 | 0.03 | -0.42, 0.42 | 0.91 |

| Baseline | | Age-adjusted | | | | | Age and CAG-adjusted | | | | |
|---|---|---|---|---|---|---|---|---|---|---|---|
| **GLX** | | **Inverse weighted** | | **Bootstrapped** | | | **Inverse weighted** | | **Bootstrapped** | | |
| **Measures** | **n** | **r** | ***p* value** | **r** | **95 % CIs** | ***p* value** | **r** | ***p* value** | **r** | **95 % CIs** | ***p* value** |
| cUHDRS | 41 | 0.03 | 0.85 | 0.08 | -0.27, 0.38 | 0.64 | 0.07 | 0.67 | 0.08 | -0.19, 0.35 | 0.61 |
| DBS | 41 | 0.01 | 0.96 | -0.00 | -0.29, 0.25 | 0.98 | 0.03 | 0.84 | 0.02 | -0.27, 0.28 | 0.88 |
| TFC | 41 | -0.04 | 0.80 | -0.14 | -0.30, 0.30 | 0.93 | -0.03 | 0.86 | -0.04 | -0.31, 0.32 | 0.82 |
| TMS | 41 | 0.02 | 0.91 | -0.02 | -0.35, 0.37 | 0.91 | -0.00 | 0.99 | -0.00 | -0.33, 0.32 | 0.98 |
| SDMT | 41 | 0.10 | 0.54 | 0.14 | -0.21, 0.40 | 0.38 | 0.14 | 0.37 | 0.15 | -0.15, 0.41 | 0.30 |
| SCN | 41 | 0.09 | 0.58 | 0.12 | -0.14, 0.38 | 0.37 | 0.14 | 0.40 | 0.13 | -0.14, 0.40 | 0.36 |
| VFC | 41 | 0.15 | 0.36 | 0.19 | -0.12, 0.49 | 0.22 | 0.24 | 0.14 | 0.24 | -0.01, 0.49 | 0.06 |
| SWR | 41 | 0.05 | 0.76 | 0.10 | -0.17, 0.39 | 0.46 | 0.09 | 0.57 | 0.11 | -0.08, 0.36 | 0.31 |
| Whole brain | 40 | 0.02 | 0.90 | 0.05 | -0.30, 0.33 | 0.75 | 0.05 | 0.78 | 0.04 | -0.29, 0.33 | 0.78 |
| Caudate | 38 | 0.12 | 0.49 | 0.18 | -0.17, 0.47 | 0.16 | 0.12 | 0.49 | 0.14 | -0.18, 0.44 | 0.34 |
| White matter | 41 | 0.07 | 0.64 | 0.10 | -0.18, 0.34 | 0.47 | 0.08 | 0.61 | 0.10 | -0.20, 0.34 | 0.49 |
| Grey matter | 41 | -0.08 | 0.62 | -0.05 | -0.49, 0.26 | 0.77 | -0.08 | 0.62 | -0.08 | -0.47, 0.25 | 0.66 |
| CSF NfL (log) | 41 | 0.04 | 0.78 | 0.02 | -0.30, 0.31 | 0.91 | 0.03 | 0.87 | 0.05 | -0.19, 0.31 | 0.70 |
| CSF Tau (log) | 41 | -0.14 | 0.39 | -0.19 | -0.61, 0.23 | 0.37 | -0.14 | 0.36 | -0.19 | -0.58, 0.23 | 0.36 |
| CSF mHTT | 41 | -0.13 | 0.41 | -0.16 | -0.49, 0.16 | 0.35 | -0.20 | 0.21 | -0.18 | -0.43, 0.16 | 0.20 |
| Plasma NfL (log) | 41 | 0.00 | 0.98 | -0.05 | -0.39, 0.30 | 0.77 | -0.03 | 0.87 | -0.04 | -0.36, 0.23 | 0.77 |
| Plasma Tau (log) | 41 | -0.08 | 0.61 | -0.10 | -0.37, 0.16 | 0.45 | -0.09 | 0.59 | -0.10 | -0.36, 0.17 | 0.47 |

| Follow-Up | | Age-adjusted | | | | | Age and CAG-adjusted | | | | |
|---|---|---|---|---|---|---|---|---|---|---|---|
| **GLX** | | **Inverse weighted** | | **Bootstrapped** | | | **Inverse weighted** | | **Bootstrapped** | | |
| **Measures** | **n** | **r** | ***p* value** | **r** | **95 % CIs** | ***p* value** | **r** | ***p* value** | **r** | **95 % CIs** | ***p* value** |
| cUHDRS | 35 | 0.36 | **0.03** | 0.38 | 0.04, 0.60 | **0.006** | 0.38 | **0.02** | 0.41 | 0.16, 0.58 | **<0.001** |
| DBS | 36 | -0.21 | 0.23 | -0.18 | -0.46, 0.23 | 0.30 | -0.07 | 0.68 | -0.05 | -0.38, 0.36 | 0.80 |
| TFC | 36 | 0.30 | 0.08 | 0.31 | -0.06, 0.58 | 0.06 | 0.25 | 0.14 | 0.28 | 0.00, 0.50 | **0.03** |
| TMS | 36 | -0.45 | **0.006** | -0.45 | -0.64, -0.19 | **<0.001** | -0.47 | **0.004** | -0.47 | -0.63, -0.28 | **<0.001** |
| SDMT | 35 | 0.31 | 0.07 | 0.32 | 0.04, 0.52 | **0.01** | 0.29 | 0.09 | 0.31 | 0.08, 0.48 | **0.002** |
| SCN | 35 | 0.26 | 0.13 | 0.31 | -0.01, 0.57 | **0.04** | 0.24 | 0.16 | 0.31 | -0.04, 0.51 | **0.02** |
| VFC | 35 | 0.41 | **0.01** | 0.39 | 0.05, 0.67 | **0.02** | 0.43 | **0.01** | 0.40 | 0.03, 0.64 | **0.01** |
| SWR | 35 | 0.30 | 0.08 | 0.33 | 0.01, 0.60 | **0.03** | 0.29 | 0.10 | 0.34 | -0.06, 0.58 | **0.03** |
| Whole brain | 34 | 0.39 | **0.02** | 0.37 | 0.05, 0.61 | **0.01** | 0.37 | **0.03** | 0.36 | 0.04, 0.60 | **0.02** |
| Caudate | 34 | 0.31 | 0.07 | 0.32 | 0.03, 0.61 | **0.02** | 0.30 | 0.08 | 0.32 | 0.04, 0.56 | **0.02** |
| White matter | 33 | 0.48 | **0.004** | 0.45 | 0.09, 0.68 | **0.002** | 0.47 | **0.01** | 0.44 | 0.10, 0.67 | **0.002** |
| Grey matter | 33 | 0.36 | **0.04** | 0.38 | 0.01, 0.63 | **0.02** | 0.34 | 0.05 | 0.37 | -0.03, 0.63 | **0.03** |
| CSF NfL (log) | 35 | -0.16 | 0.37 | -0.15 | -0.46, 0.17 | 0.35 | -0.08 | 0.63 | -0.09 | -0.38, 0.19 | 0.53 |
| CSF Tau (log) | 35 | -0.21 | 0.22 | -0.25 | -0.56, 0.09 | 0.14 | -0.17 | 0.32 | -0.22 | -0.51, 0.11 | 0.16 |
| CSF mHTT | 34 | -0.47 | **0.004** | -0.48 | -0.68, -0.16 | **<0.001** | -0.53 | **0.001** | -0.53 | -0.70, -0.25 | **<0.001** |
| Plasma NfL (log) | 36 | -0.14 | 0.42 | -0.12 | -0.40, 0.18 | 0.41 | -0.02 | 0.91 | -0.00 | -0.20, 0.21 | 0.97 |
| Plasma Tau (log) | 36 | -0.03 | 0.87 | 0.00 | -0.36, 0.33 | 1.00 | 0.02 | 0.91 | 0.04 | -0.31, 0.37 | 0.83 |

*Relationships between MRS metabolites and clinical, cognitive, imaging and biofluid markers were assessed at baseline and follow-up using Pearson's partial correlation controlling for age, and age and CAG repeat length. Correlation coefficients and 95% confidence intervals were computed using bootstrap testing with 1000 repetitions. A weighted correlation was also conducted, applying an inverse weighting to %SD value. Results displayed are unadjusted for multiplicity. Bold text indicated significance p < 0.05. cUHDRS, composite Unified Huntington's Disease Rating Scale; DBS, Disease Burden Score; TFC, Total Functional Capacity; TMS, Total Motor Score; SDMT, Symbol Digit Modalities Test; SCN, Stroop Colour Naming; VFC, Verbal Fluency Categorical; SWR, Stroop Word Reading Test; NfL, Neurofilament Light Chain; mHTT, Mutant Huntingtin.*

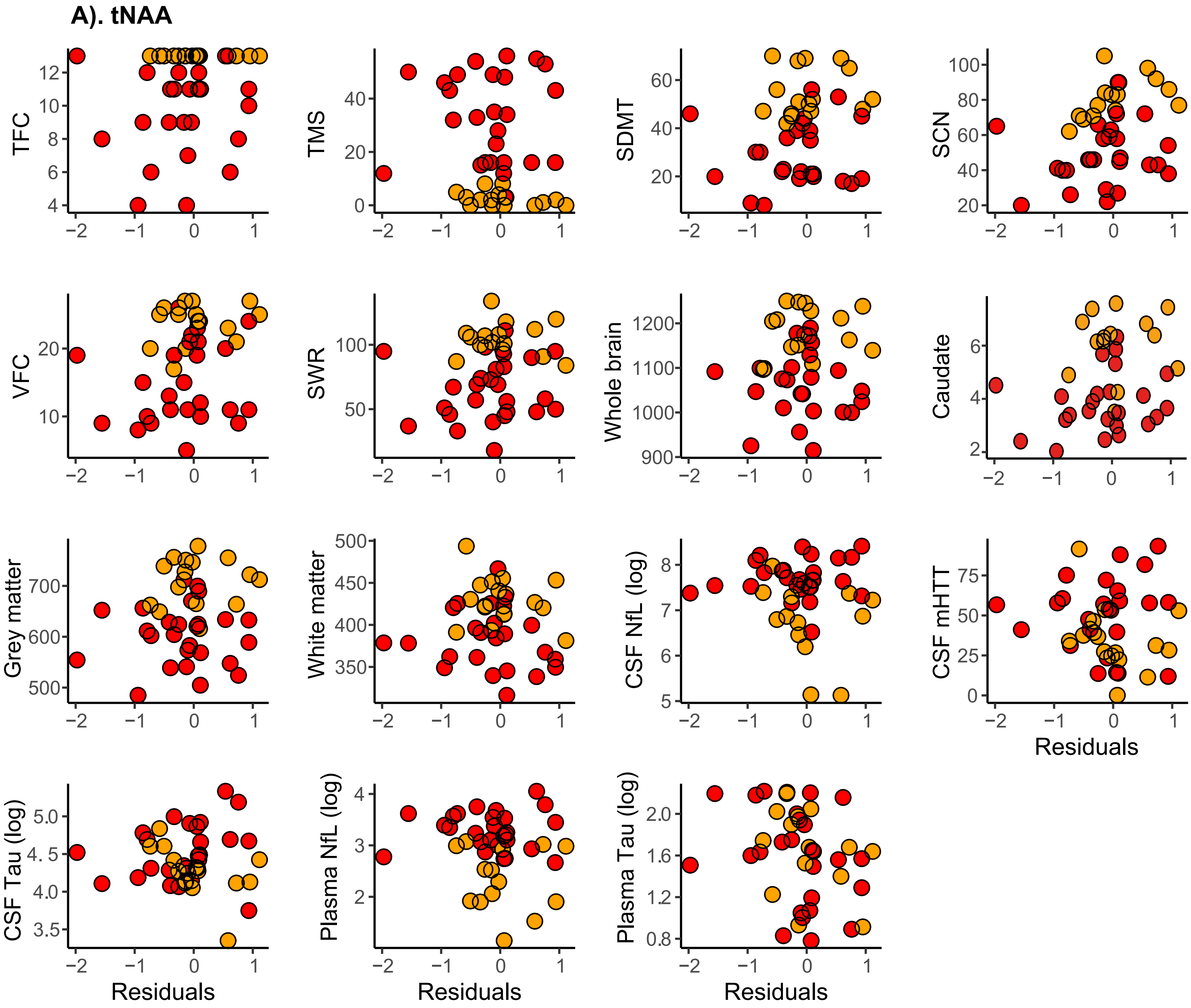
A). tNAA
TFC
TMS
SDMT
SCN
VFC
SWR
Whole brain
Caudate
Grey matter
White matter
CSF NfL (log)
CSF mHTT
CSF Tau (log)
Plasma NfL (log)
Plasma Tau (log)
Residuals

**B). tCre**

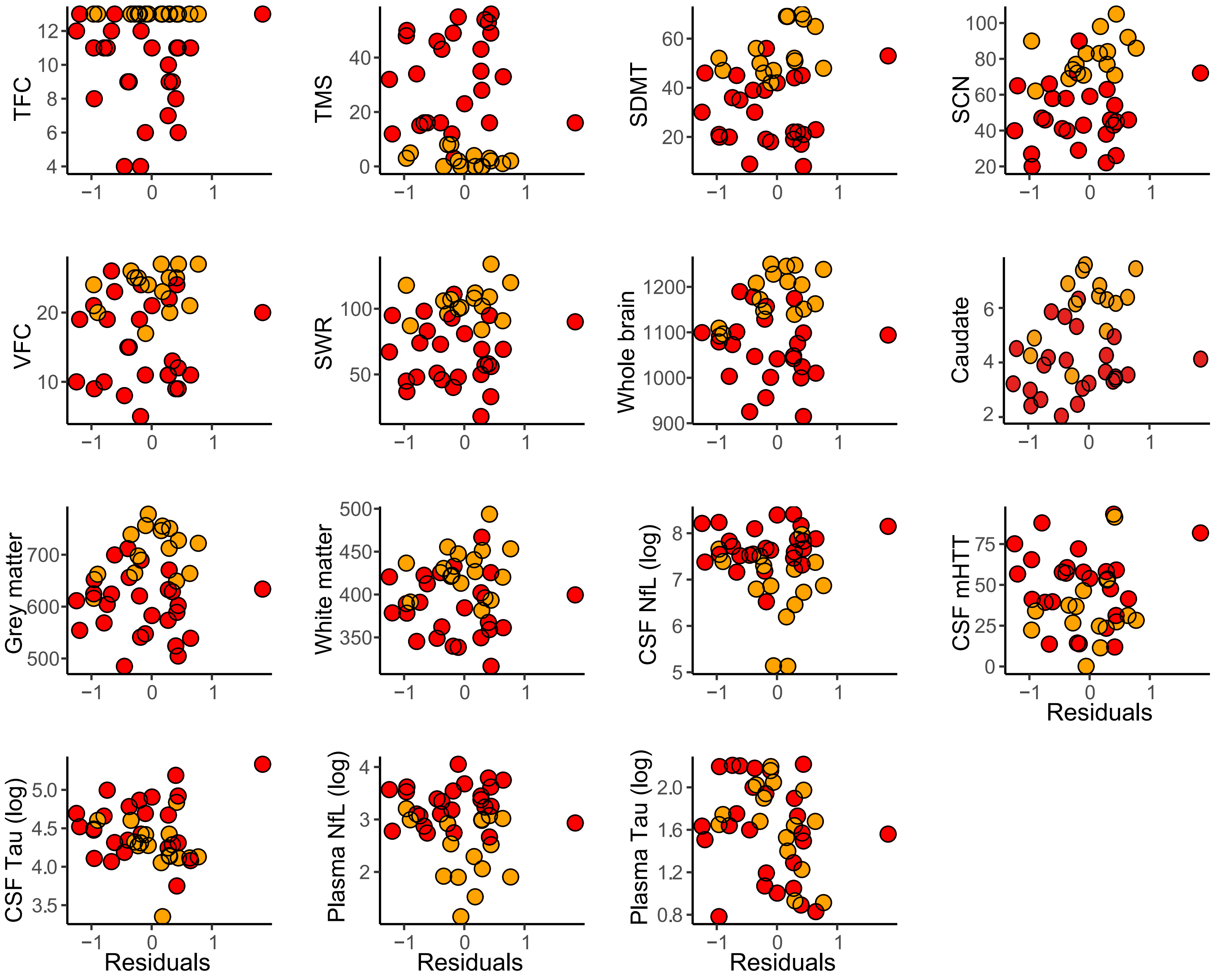

TFC
TMS
SDMT
SCN
VFC
SWR
Whole brain
Caudate
Grey matter
White matter
CSF NfL (log)
CSF mHTT
CSF Tau (log)
Plasma NfL (log)
Plasma Tau (log)
Residuals
Residuals
Residuals
Residuals

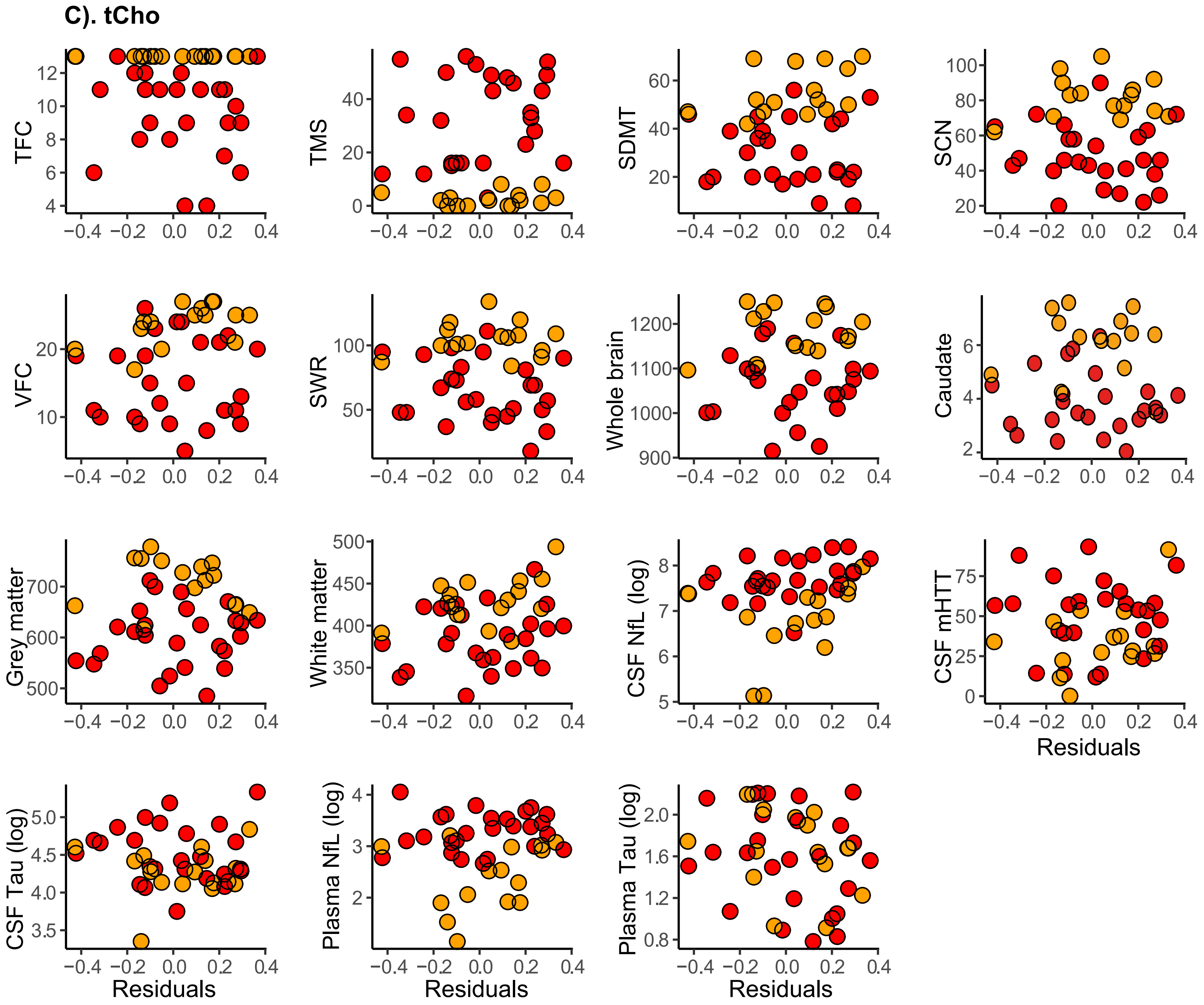

C). tCho
TFC
TMS
SDMT
SCN
VFC
SWR
Whole brain
Caudate
Grey matter
White matter
CSF NfL (log)
CSF mHTT
CSF Tau (log)
Plasma NfL (log)
Plasma Tau (log)
Residuals

**D). MI**

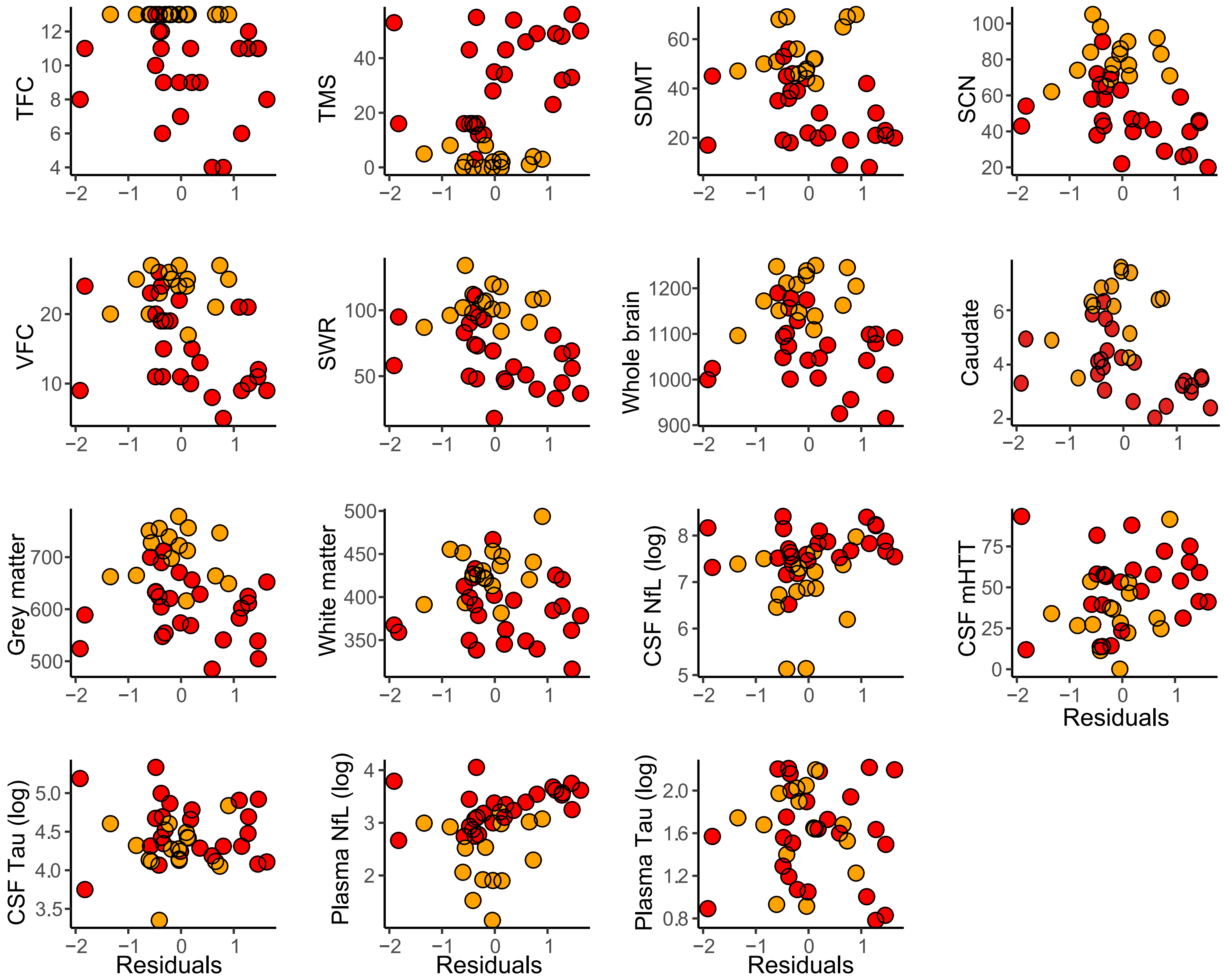
TFC
TMS
SDMT
SCN
VFC
SWR
Whole brain
Caudate
Grey matter
White matter
CSF NfL (log)
CSF mHTT
CSF Tau (log)
Plasma NfL (log)
Plasma Tau (log)
Residuals

**E). GSH**

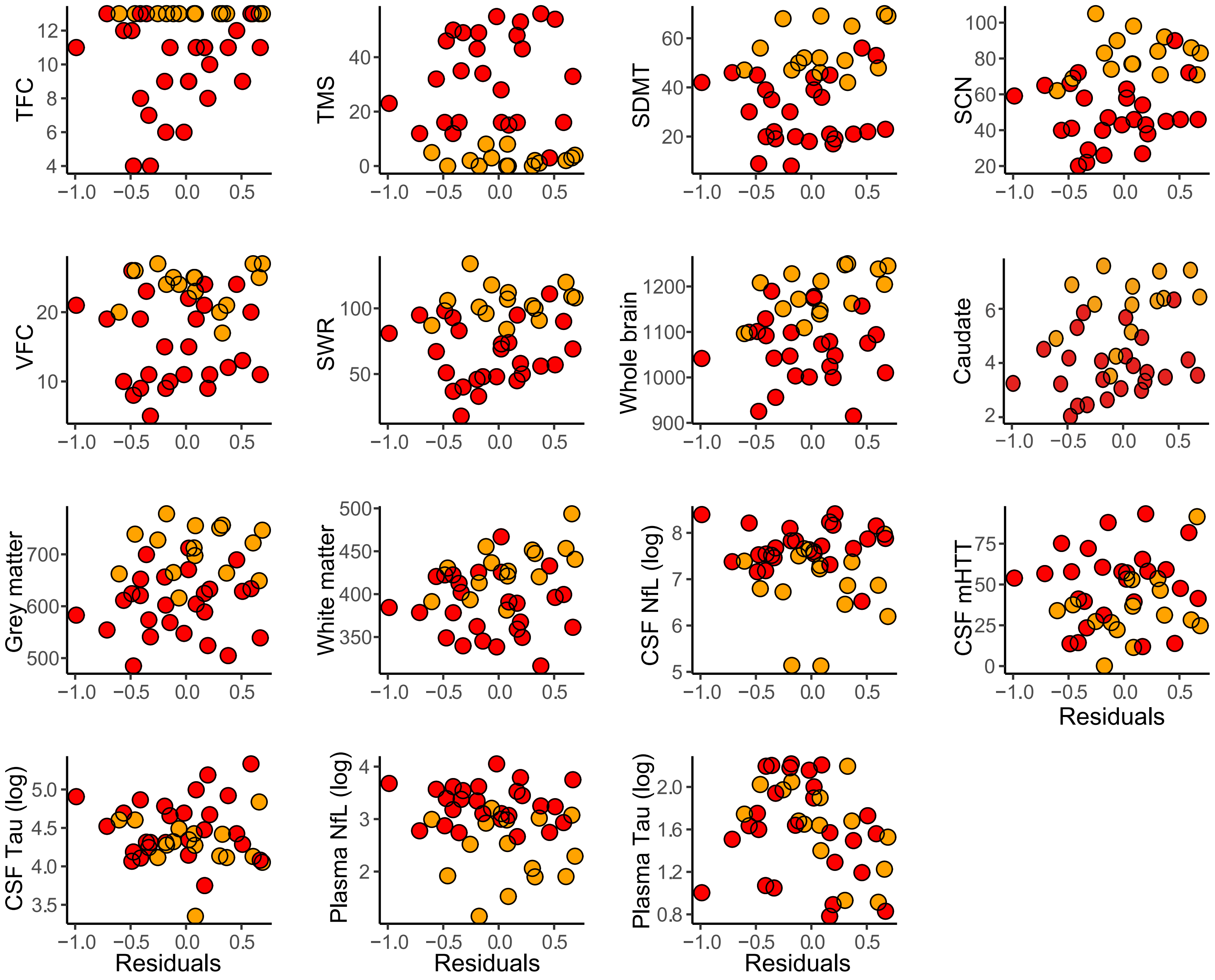

TFC
TMS
SDMT
SCN
VFC
SWR
Whole brain
Caudate
Grey matter
White matter
CSF NfL (log)
CSF mHTT
CSF Tau (log)
Plasma NfL (log)
Plasma Tau (log)
Residuals

**F). GABA**

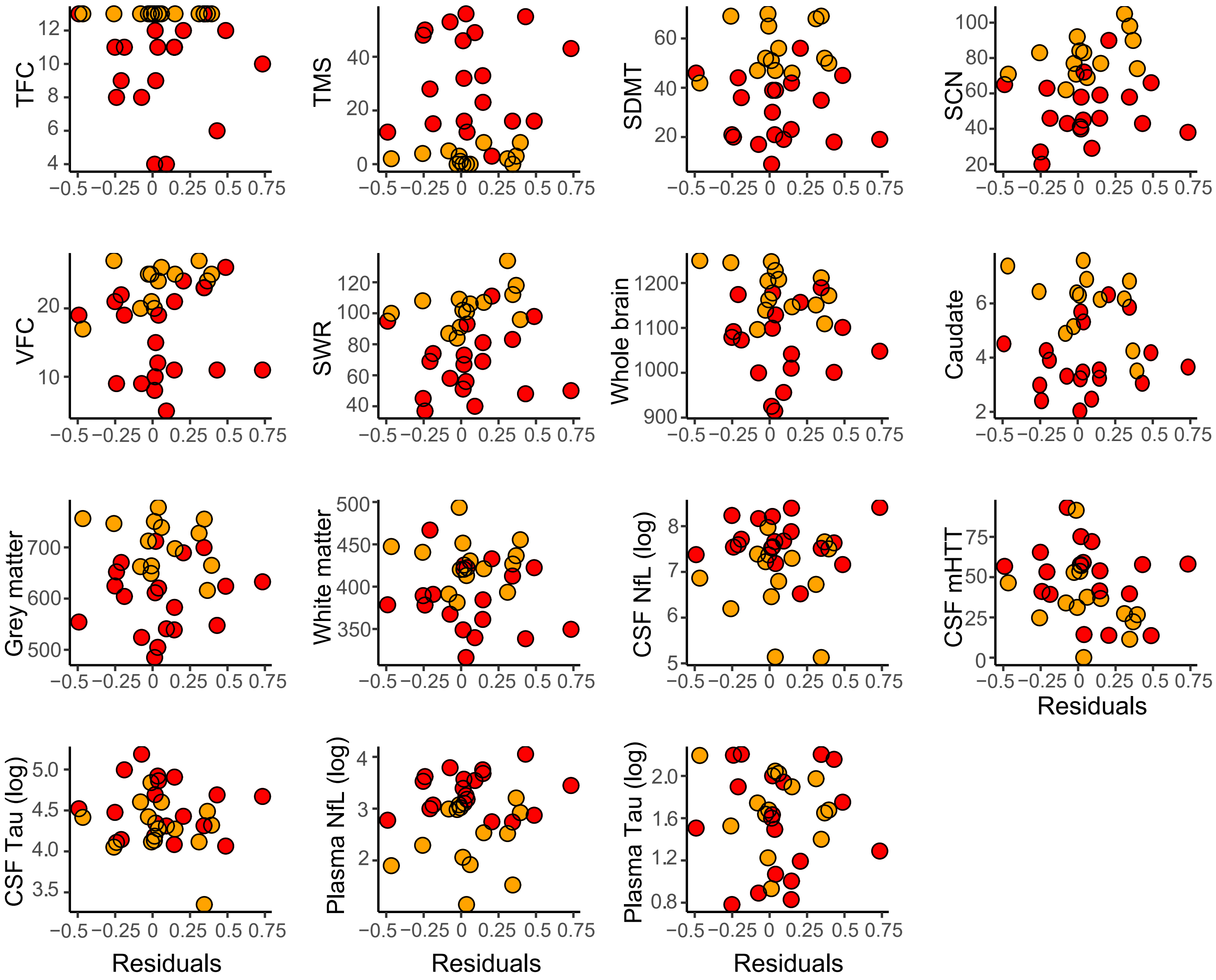
TFC
TMS
SDMT
SCN
VFC
SWR
Whole brain
Caudate
Grey matter
White matter
CSF NfL (log)
CSF mHTT
CSF Tau (log)
Plasma NfL (log)
Plasma Tau (log)
Residuals
−0.5 −0.25 0 0.25 0.5 0.75

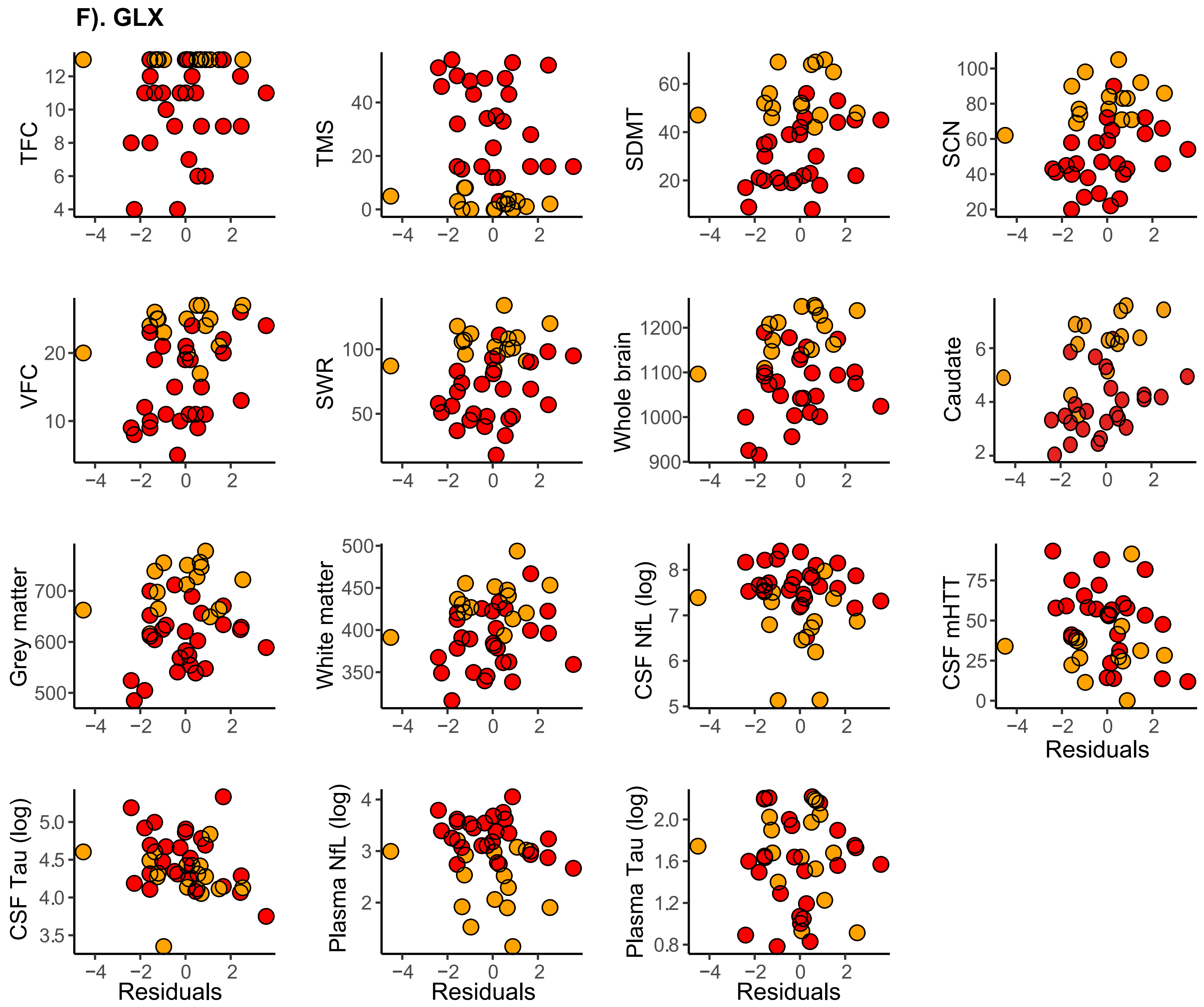


**Supplementary Fig. 3: Scatterplots of Correlations Between Metabolites and Established Measures (Baseline)**. Scatter plots displaying associations between tNAA (**A**), tCre (**B**), tCho (**C**), MI (**D**), GSH (**E**), GABA (**F**), GLX (**G**) and measures of clinical progression, cognitive decline, imaging and biofluid markers (Baseline cohort). Values displayed are controlled for CSF PVE only. Red and yellow datapoints indicate manifest and premanifest patients, respectively. TFC, Total Functional Capacity; TMS, Total Motor Score; SDMT, Symbol Digit Modalities Test; SCN, Stroop Colour Naming; VFC, Verbal Fluency Categorical; SWR, Stroop Word Reading Test; NfL, Neurofilament Light; mHTT, mutant Huntington.

**A). tNAA - Follow-Up Cohort**

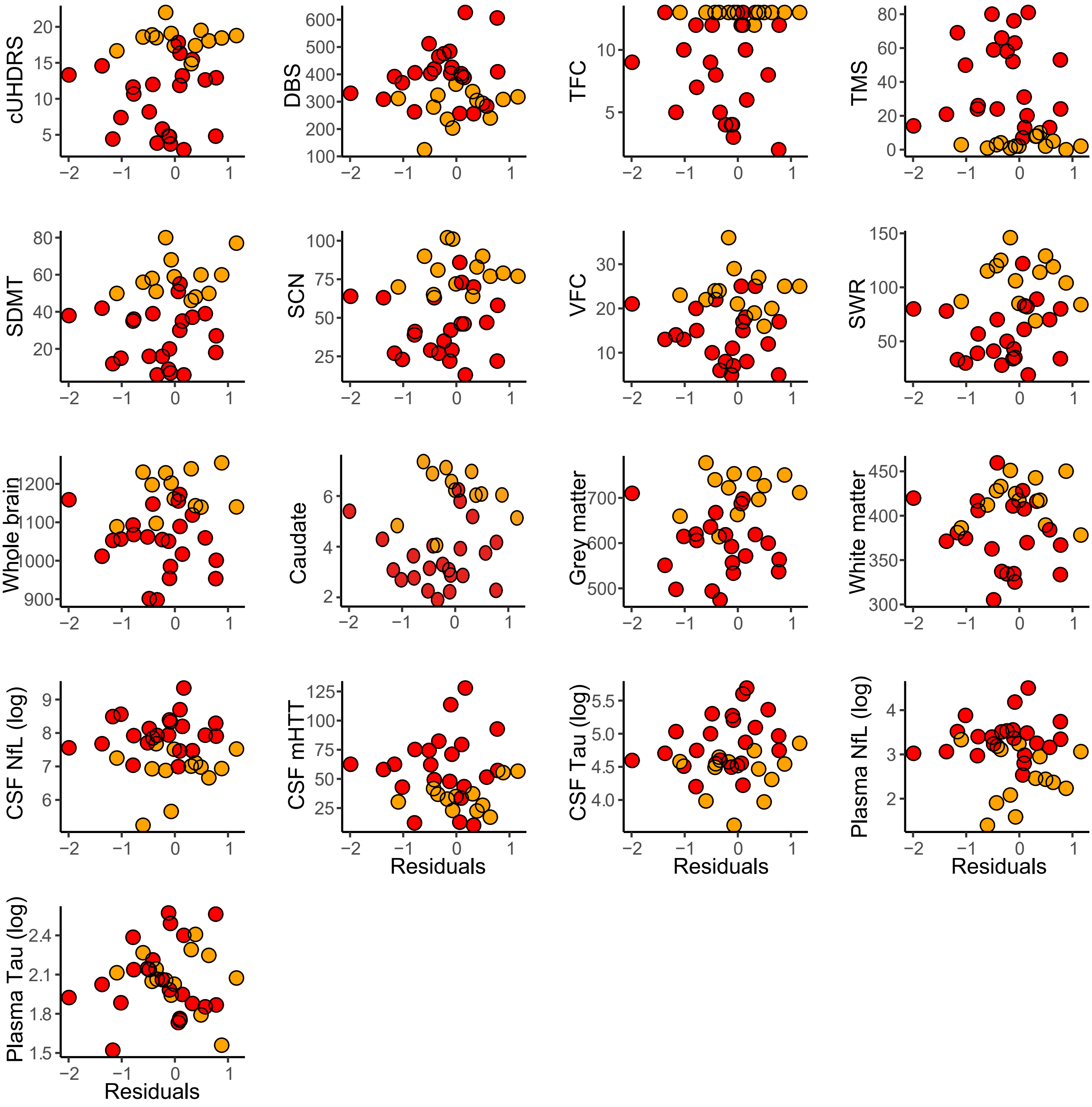

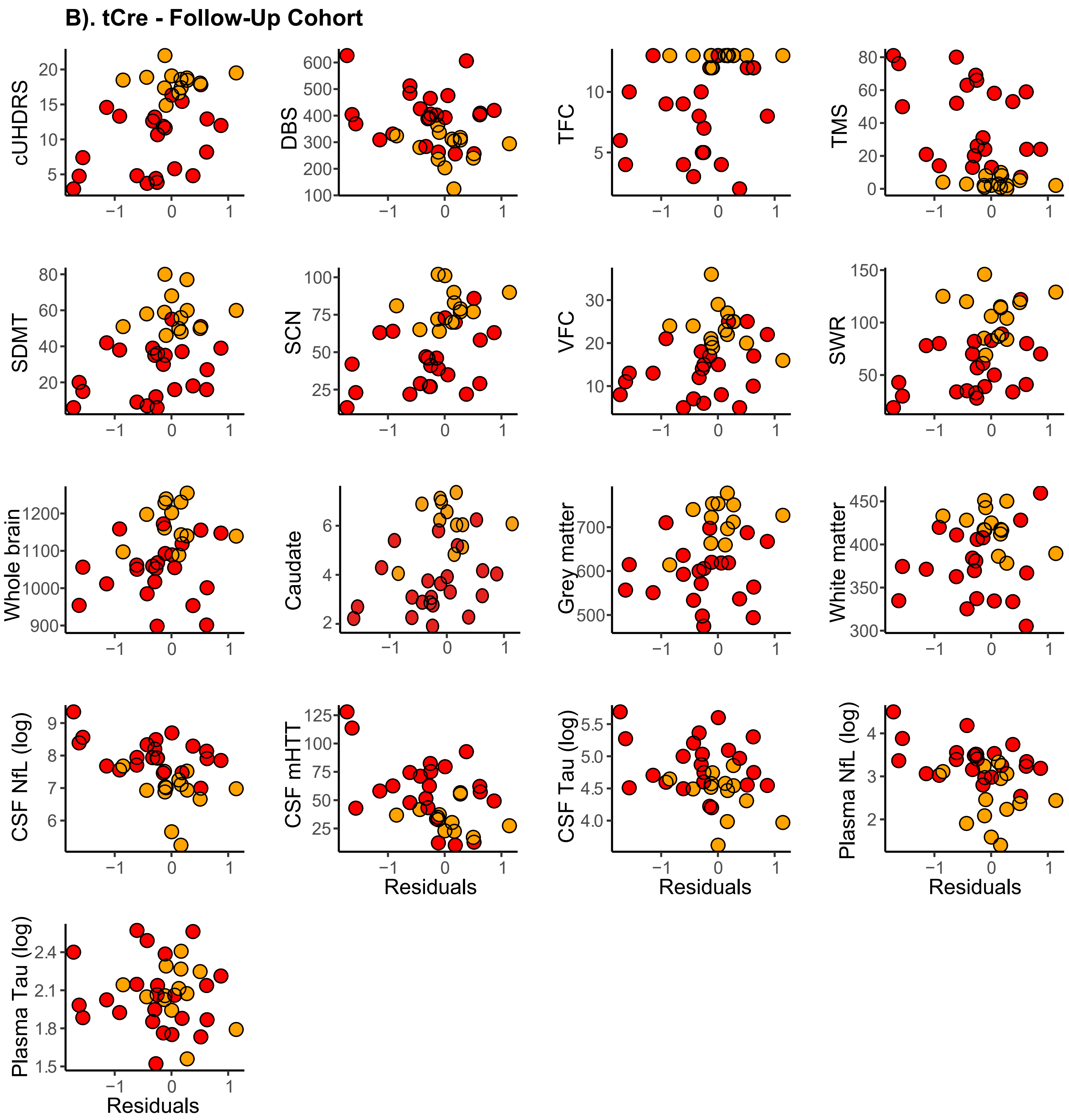
B). tCre - Follow-Up Cohort
cUHDRS
DBS
TFC
TMS
SDMT
SCN
VFC
SWR
Whole brain
Caudate
Grey matter
White matter
CSF NfL (log)
CSF mHTT
CSF Tau (log)
Plasma NfL (log)
Plasma Tau (log)
Residuals

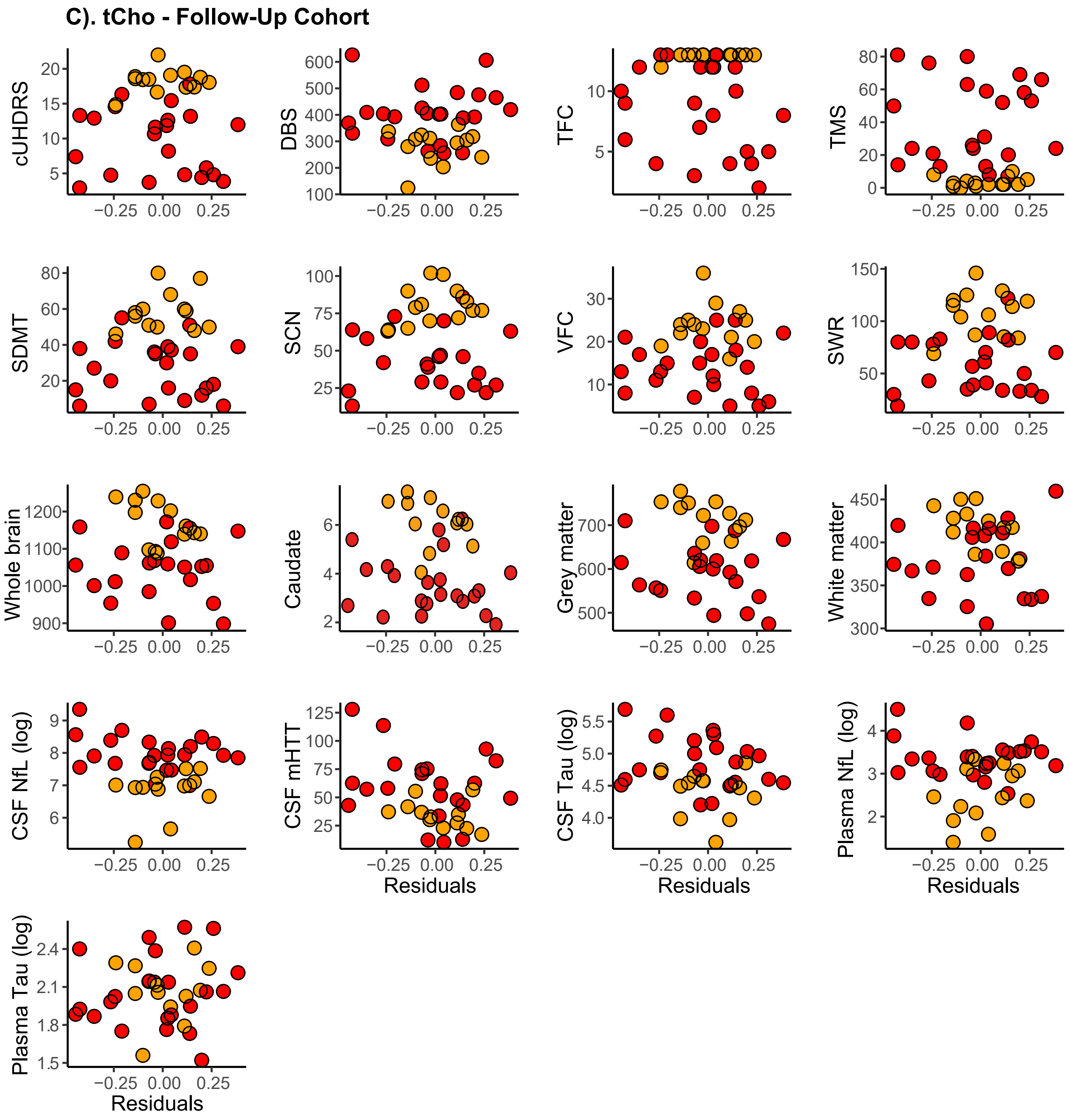
C). tCho - Follow-Up Cohort
cUHDRS
DBS
TFC
TMS
SDMT
SCN
VFC
SWR
Whole brain
Caudate
Grey matter
White matter
CSF NfL (log)
CSF mHTT
CSF Tau (log)
Plasma NfL (log)
Plasma Tau (log)
Residuals

**D). MI - Follow-Up Cohort**

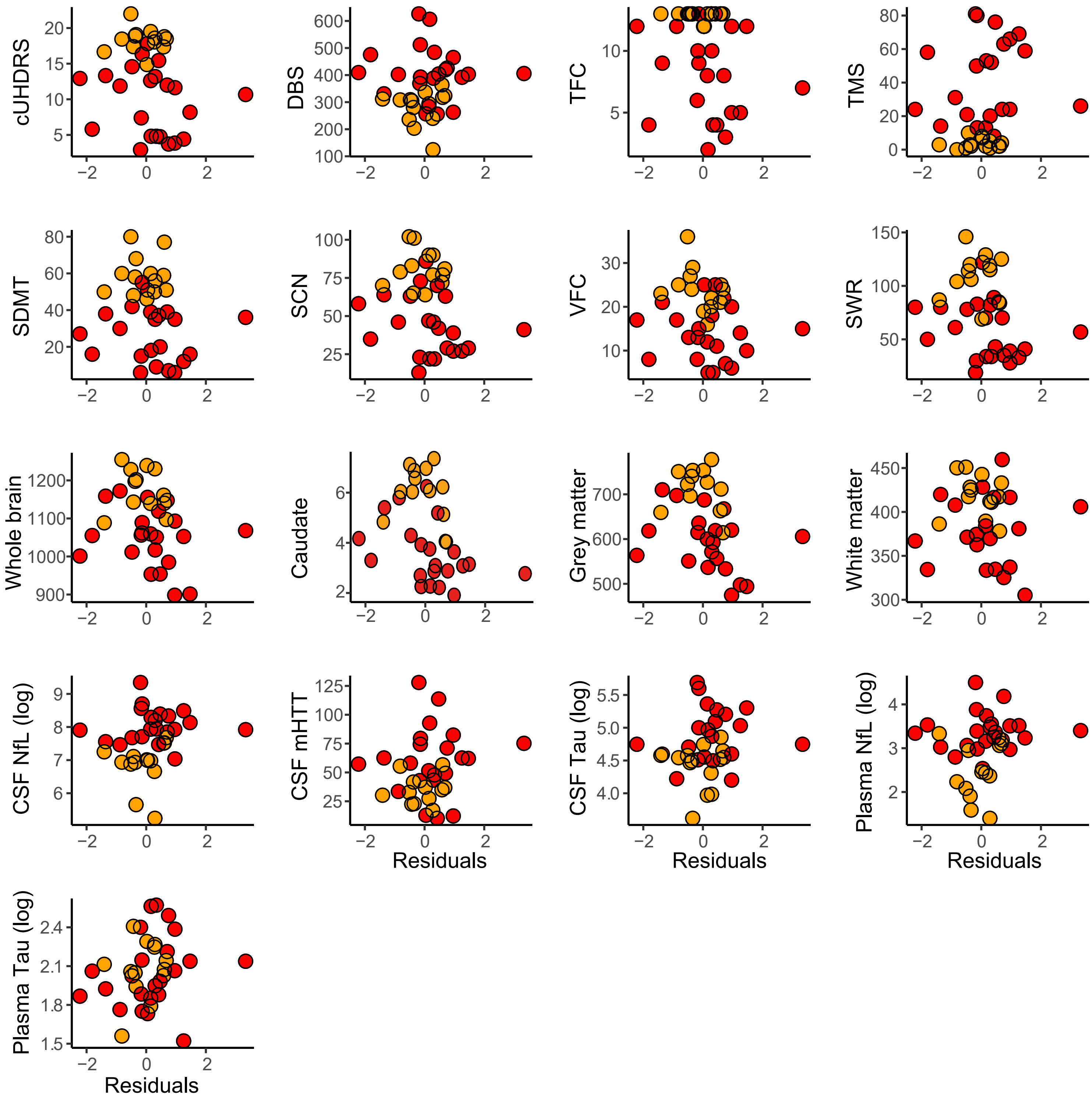

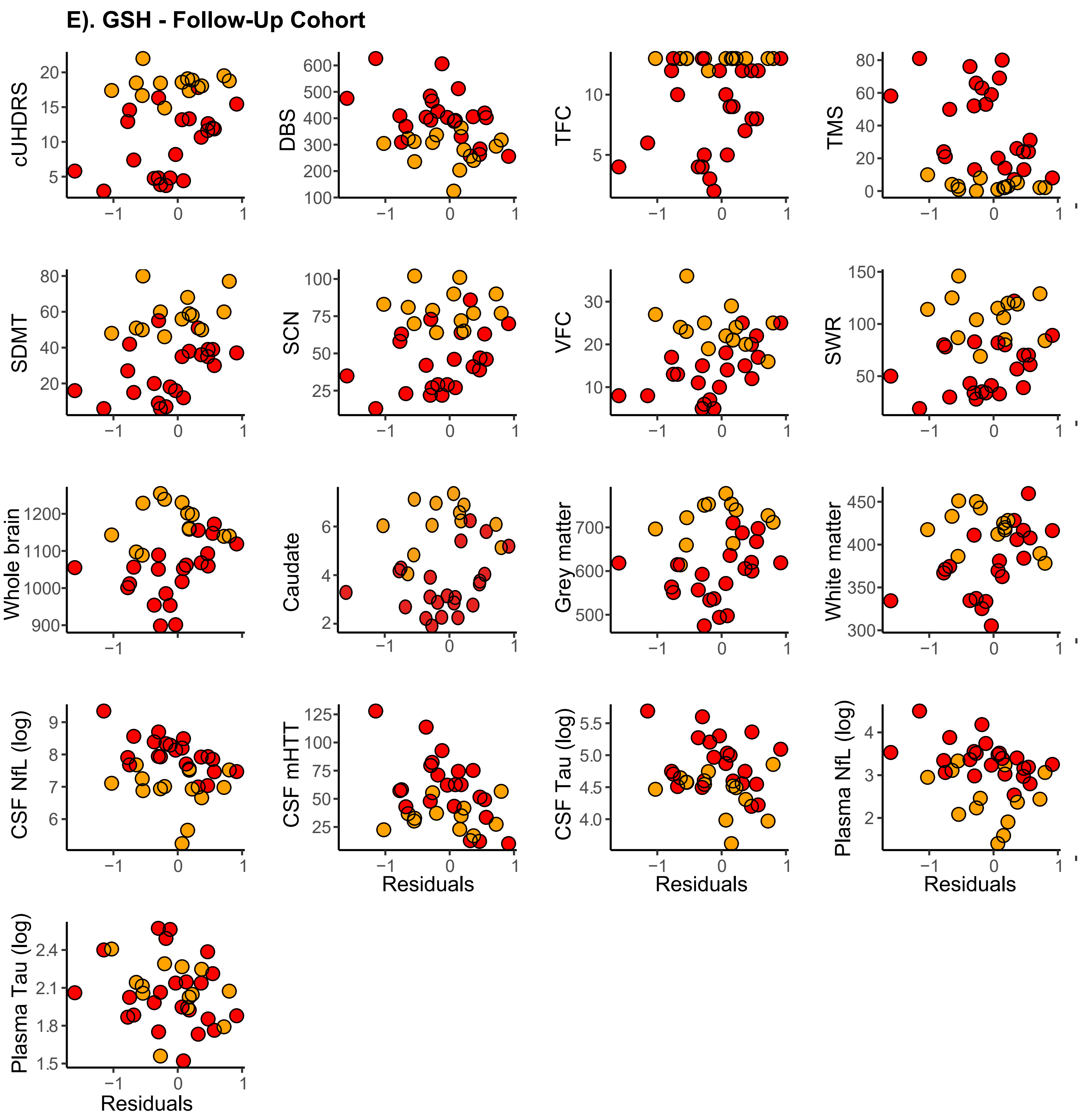
E). GSH - Follow-Up Cohort
cUHDRS
DBS
TFC
TMS
SDMT
SCN
VFC
SWR
Whole brain
Caudate
Grey matter
White matter
CSF NfL (log)
CSF mHTT
CSF Tau (log)
Plasma NfL (log)
Plasma Tau (log)
Residuals

### F). GABA - Follow-Up Cohort

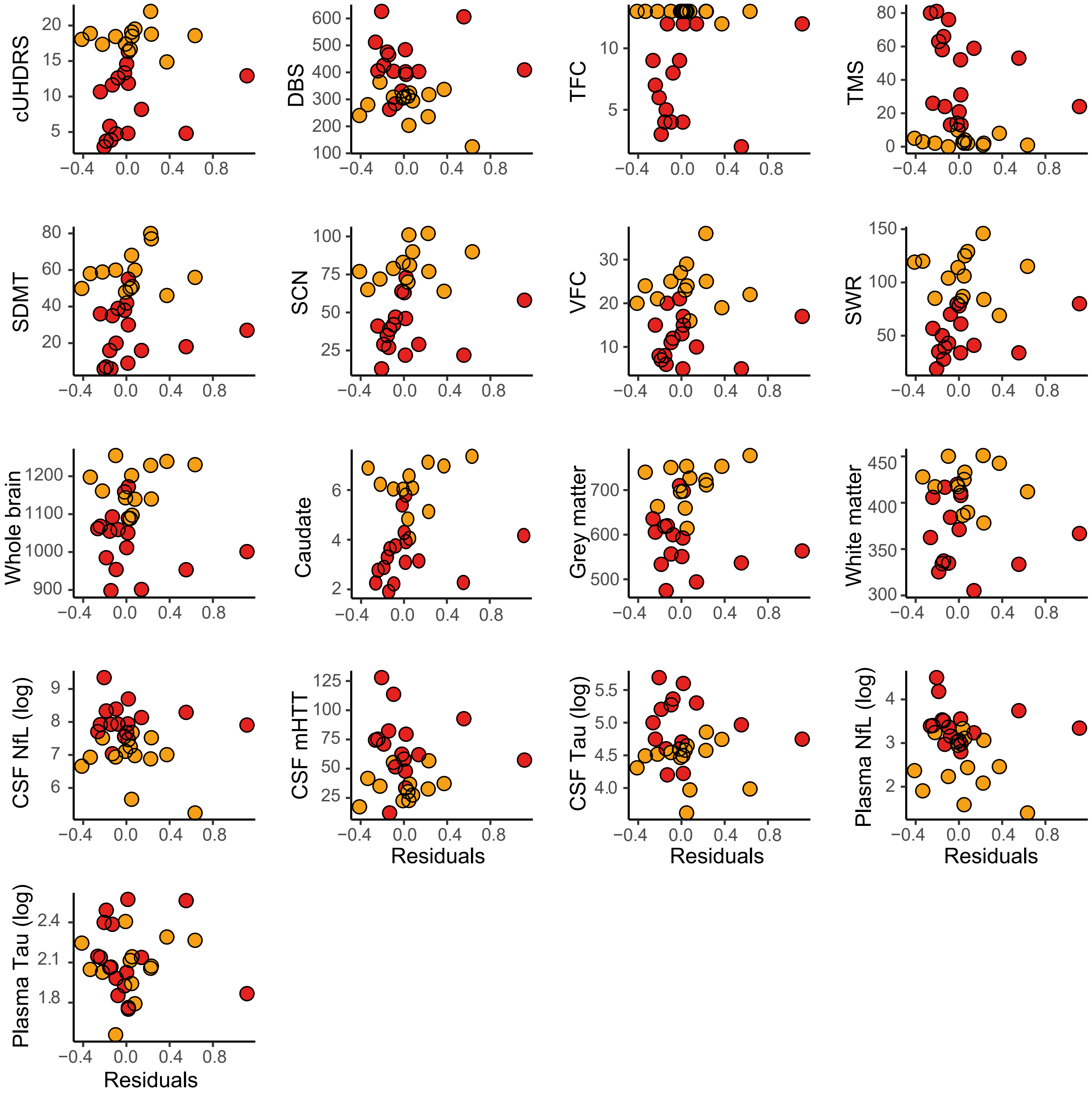

### G). GLX - Follow-Up Cohort

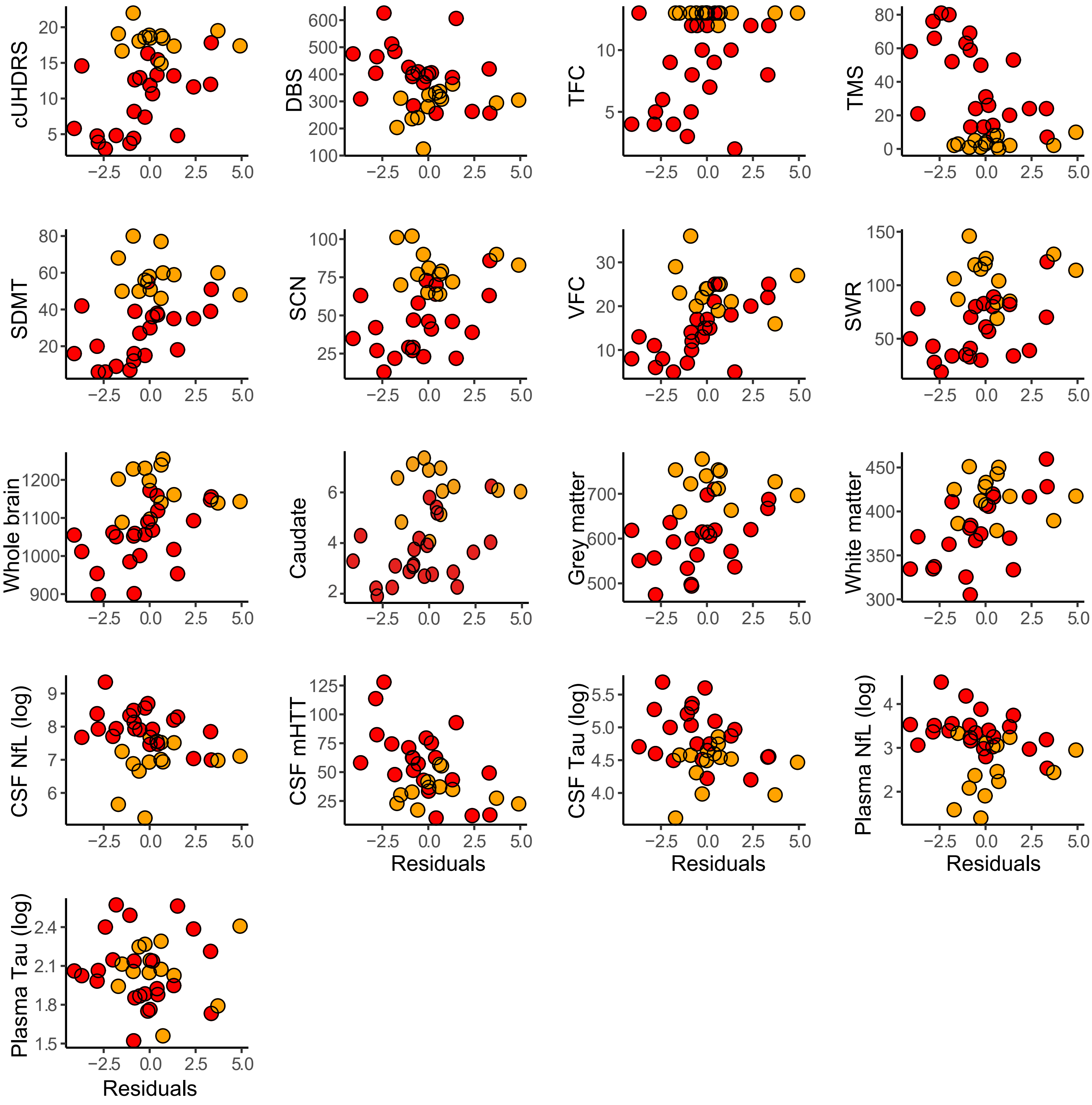


**Supplementary Fig. 4: Scatterplots of Correlations Between Metabolites and Established Measures (follow-up)**. Scatter plots displaying associations between tNAA (**A**), tCre (**B**), tCho (**C**), MI (**D**), GSH (**E**), GABA (**F**), GLX (**G**) and measures of clinical progression, cognitive decline, imaging and biofluid markers (follow-up cohort). Values displayed are controlled for CSF PVE only. Red and yellow datapoints indicate manifest and premanifest patients, respectively. cUHDRS, Composite Unified Huntingtons Disease Rating Scale; DBS, Disease Burden Score; TFC, Total Functional Capacity; TMS, Total Motor Score; SDMT, Symbol Digit Modalities Test; SCN, Stroop Colour Naming; VFC, Verbal Fluency Categorical; SWR, Stroop Word Reading Test; NfL, Neurofilament Light; mHTT, mutant Huntington.

**A) tNAA**

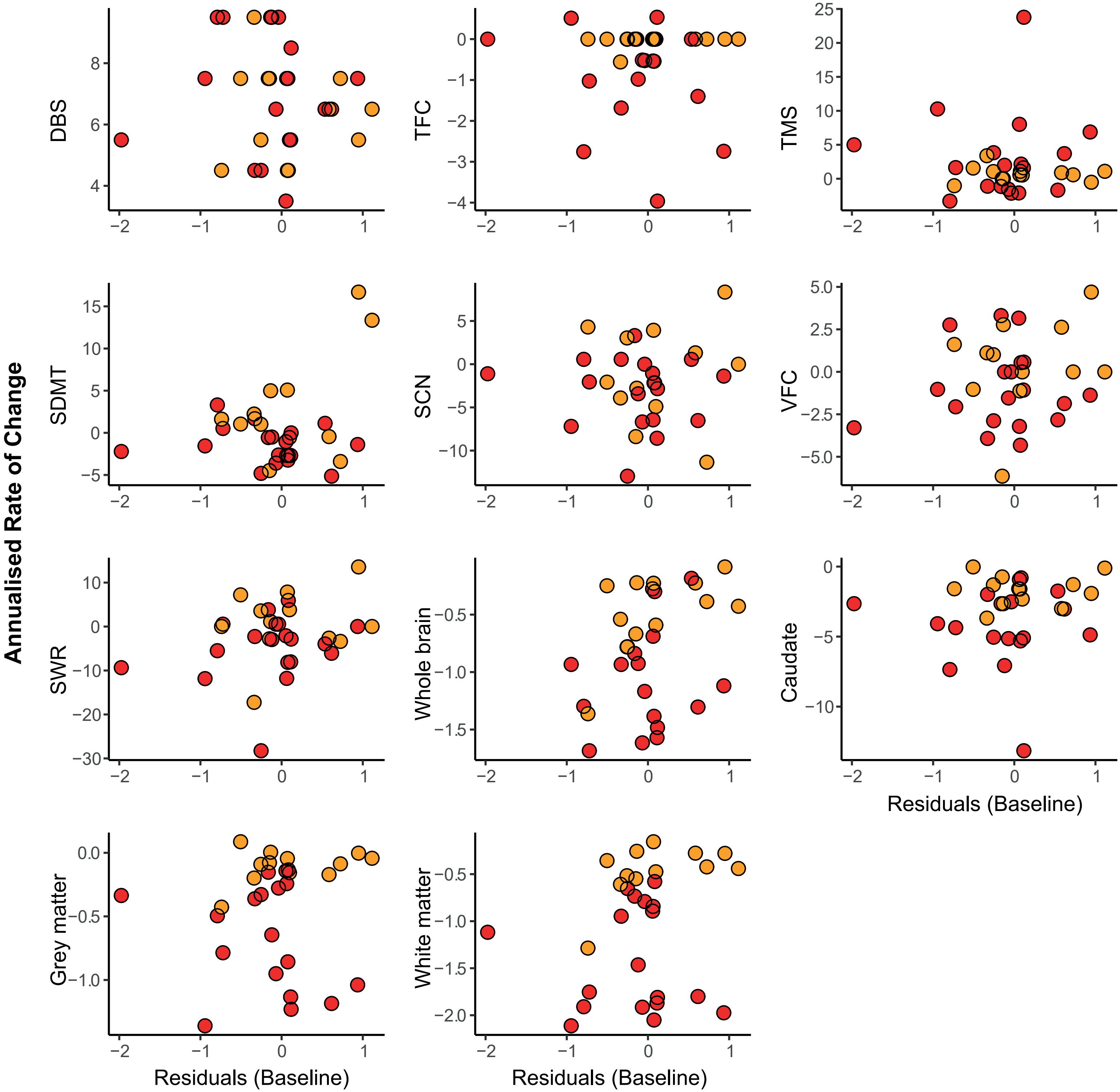

Annualised Rate of Change
DBS
TFC
TMS
SDMT
SCN
VFC
SWR
Whole brain
Caudate
Grey matter
White matter
Residuals (Baseline)
Residuals (Baseline)
Residuals (Baseline)

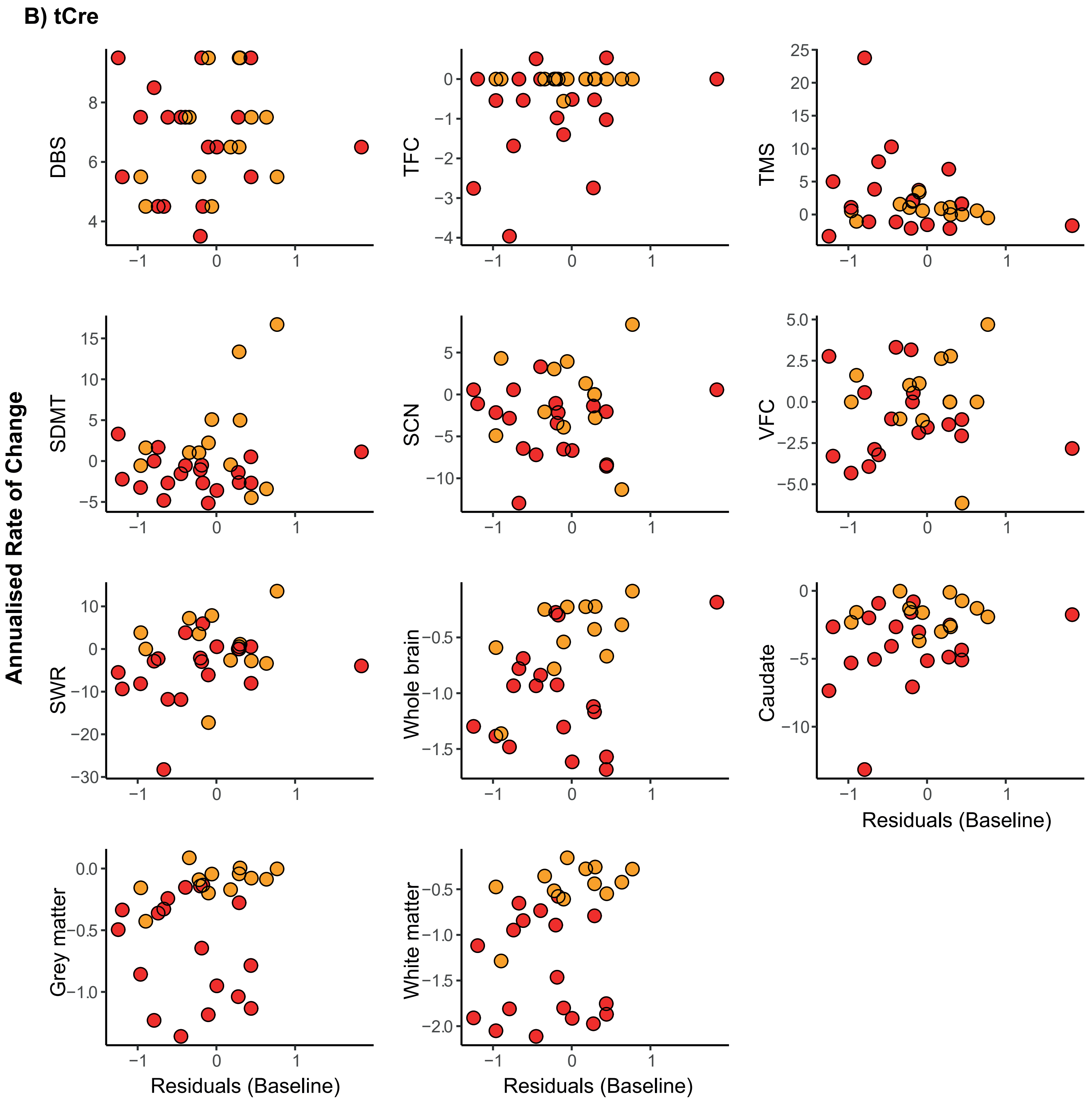
B) tCre
Annualised Rate of Change
DBS
TFC
TMS
SDMT
SCN
VFC
SWR
Whole brain
Caudate
Grey matter
White matter
Residuals (Baseline)
Residuals (Baseline)
Residuals (Baseline)

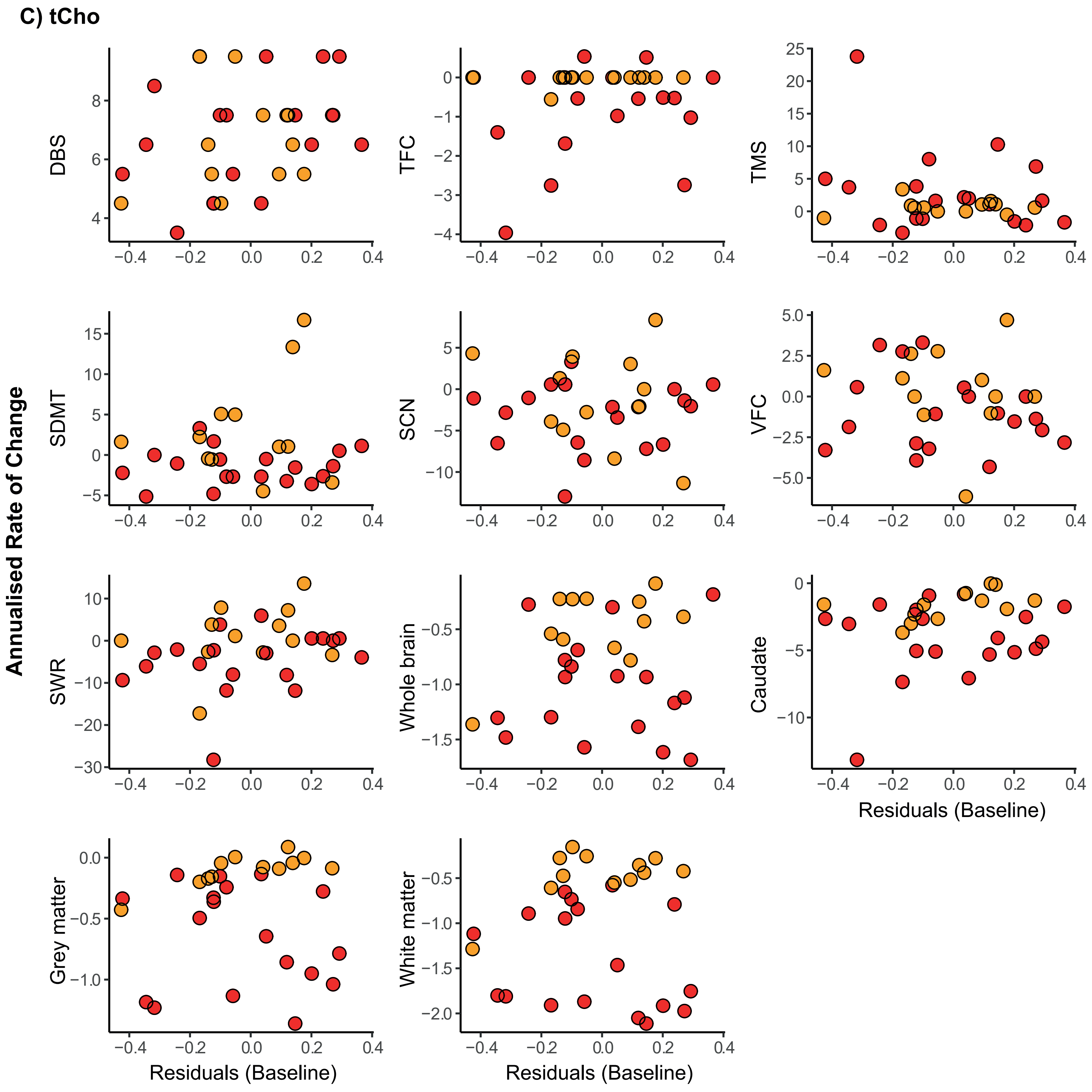

C) tCho
Annualised Rate of Change
DBS
TFC
TMS
SDMT
SCN
VFC
SWR
Whole brain
Caudate
Grey matter
White matter
Residuals (Baseline)
Residuals (Baseline)
Residuals (Baseline)

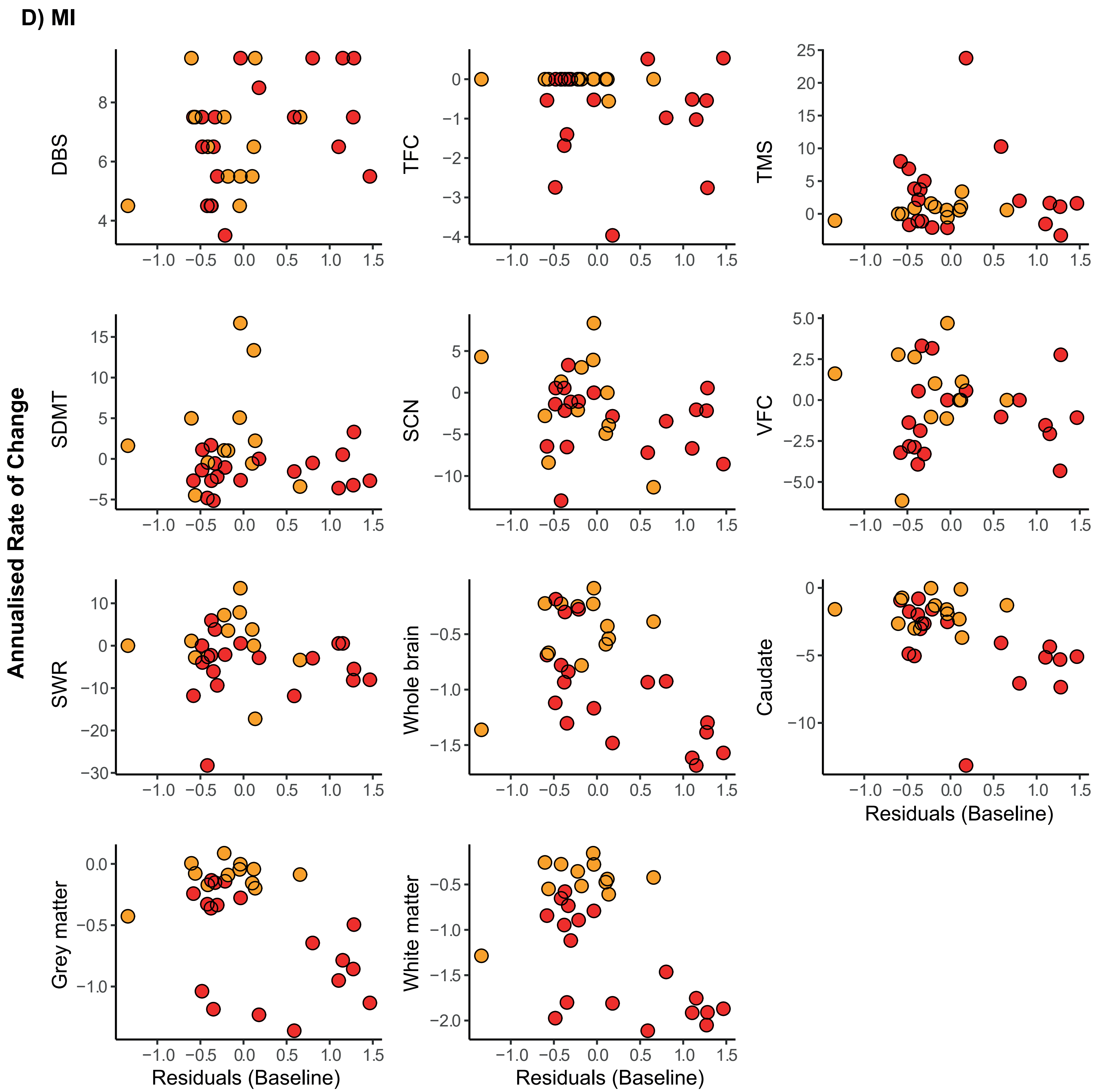
D) MI
Annualised Rate of Change
DBS
TFC
TMS
SDMT
SCN
VFC
SWR
Whole brain
Caudate
Grey matter
White matter
Residuals (Baseline)
Residuals (Baseline)
Residuals (Baseline)

**E) GSH**

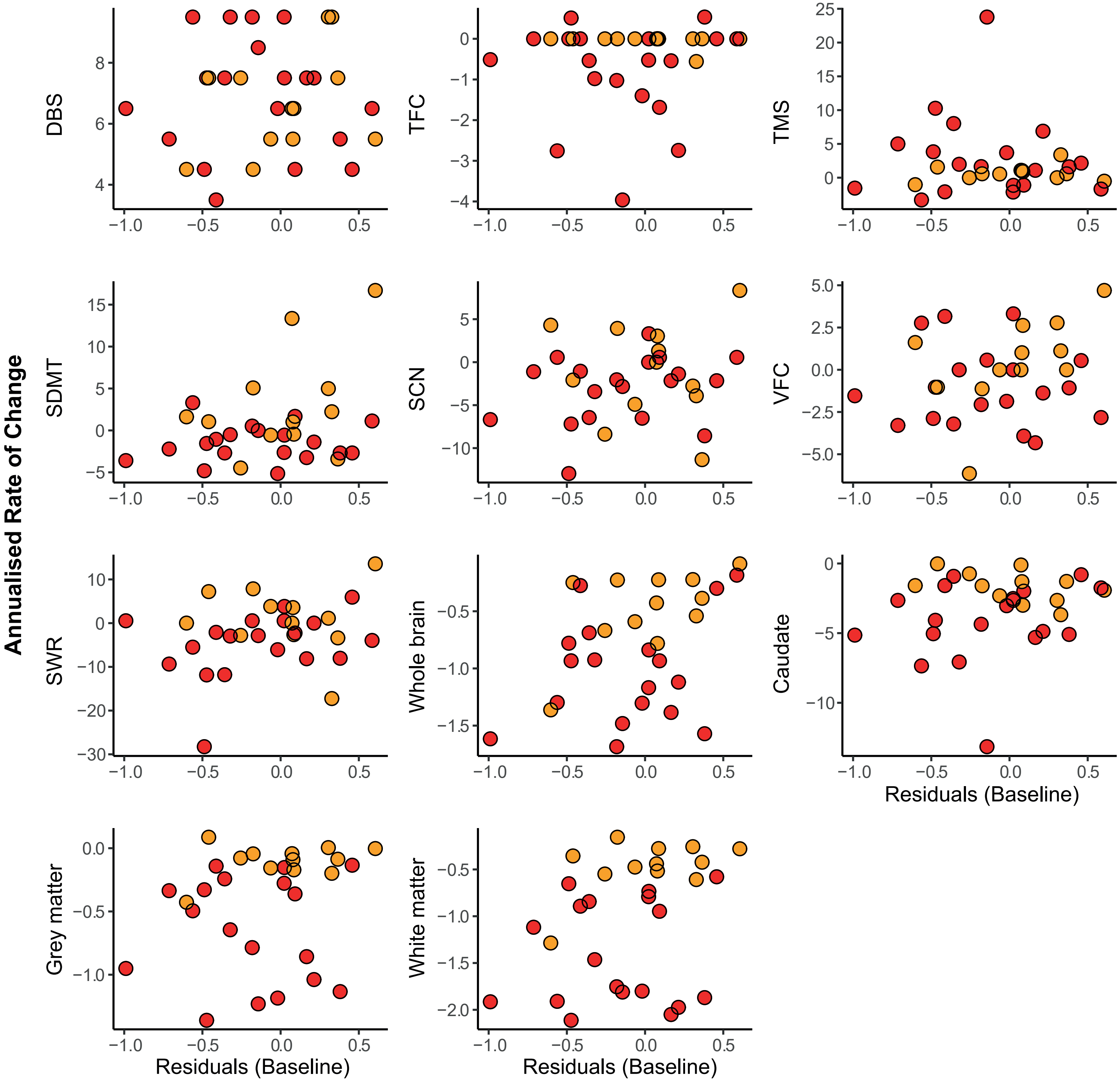

Annualised Rate of Change
DBS
TFC
TMS
SDMT
SCN
VFC
SWR
Whole brain
Caudate
Grey matter
White matter
Residuals (Baseline)
Residuals (Baseline)
Residuals (Baseline)

### F) GABA

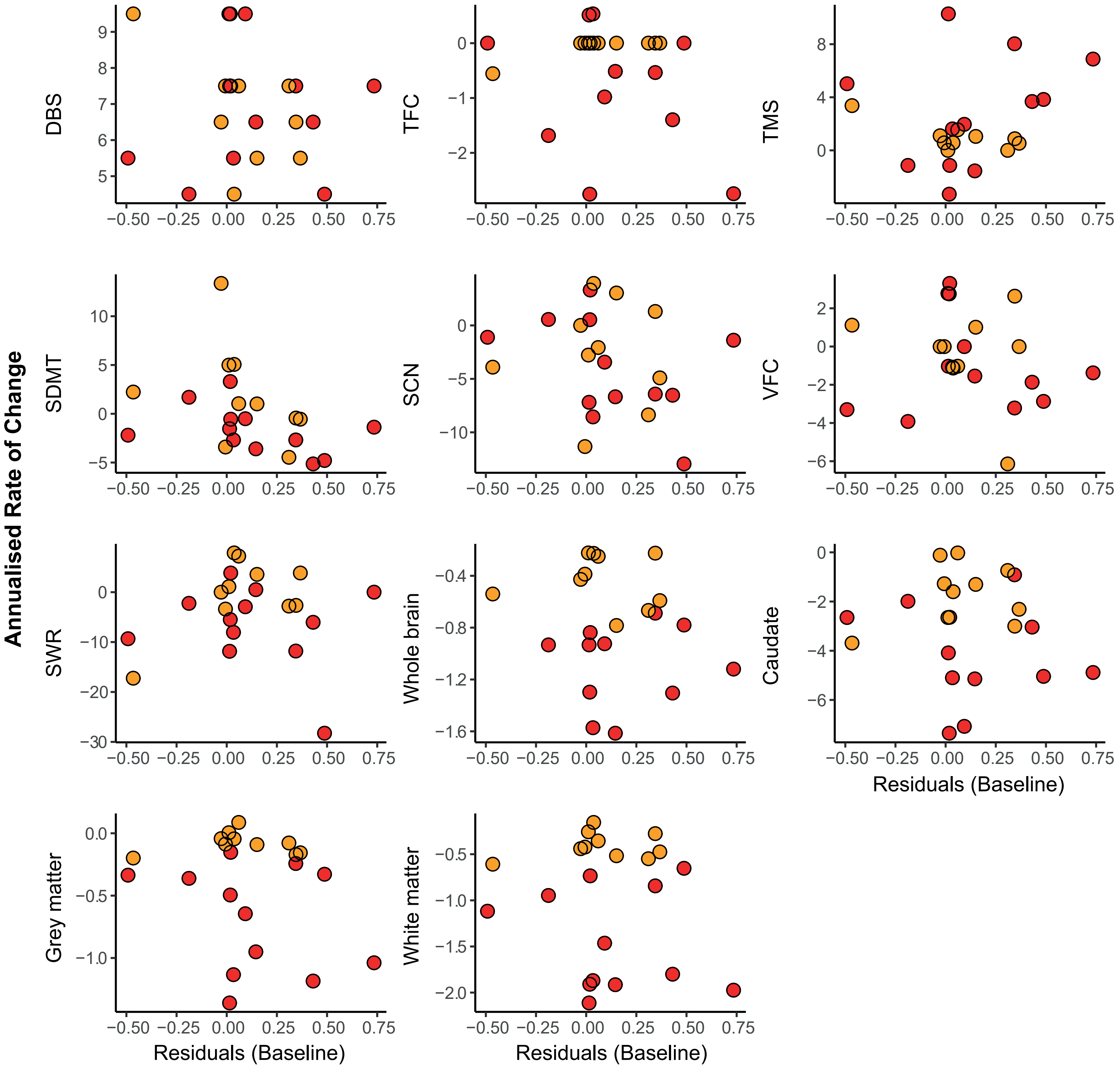

Annualised Rate of Change
DBS
TFC
TMS
SDMT
SCN
VFC
SWR
Whole brain
Caudate
Grey matter
White matter
Residuals (Baseline)

**G) GLX**

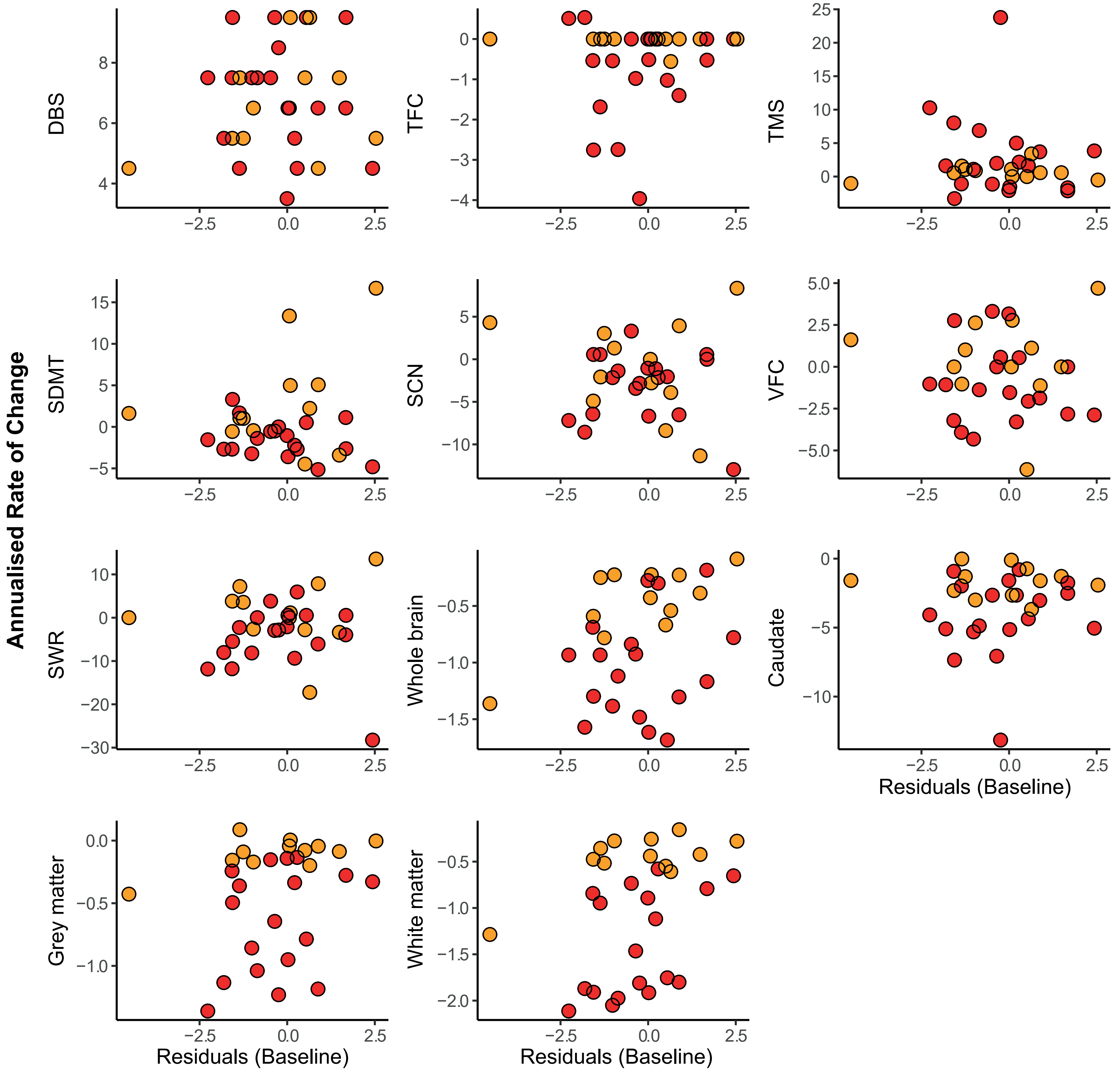


**Supplementary Fig. 5: Scatterplots of Correlations Between Baseline Metabolite Values and Annualised Rate of Change in Established Measures**. Scatter plots displaying associations between baseline tNAA (**A**), tCre (**B**), tCho (**C**), MI (**D**), GSH (**E**), GABA (**F**), GLX (**G**) and annualised rate of change in measures of clinical progression, cognitive decline, imaging markers. Values displayed are controlled for CSF PVE only. Red and yellow datapoints indicate manifest and premanifest patients, respectively. cUHDRS, Composite Unified Huntington's Disease Rating Scale; DBS, Disease Burden Score; TFC, Total Functional Capacity; TMS, Total Motor Score; SDMT, Symbol Digit Modalities Test; SCN, Stroop Colour Naming; VFC, Verbal Fluency Categorical; SWR, Stroop Word Reading Test.

**Supplementary Table 6 – Correlations between Baseline Metabolites and Annualised Rate of Change (Δ) in Clinical, Cognitive, and Imaging Measures.**

| | | **Age-adjusted** | | | | | **Age and CAG-adjusted** | | | | |
|---|---|---|---|---|---|---|---|---|---|---|---|
| **Baseline tNAA** | | **Inverse weighted** | | **Bootstrapped** | | | **Inverse weighted** | | **Bootstrapped** | | |
| **Measures** | **n** | **r** | ***p* value** | **r** | **95 % CIs** | ***p* value** | **r** | ***p* value** | **r** | **95 % CIs** | ***p* value** |
| Δ cUHDRS | 31 | 0.23 | 0.21 | 0.21 | -0.16, 0.53 | 0.25 | 0.23 | 0.21 | 0.18 | -0.18, 0.52 | 0.32 |
| Δ DBS | 31 | -0.03 | 0.87 | -0.10 | -0.39, 0.19 | 0.51 | 0.06 | 0.76 | 0.00 | -0.29, 0.25 | 1.00 |
| Δ TFC | 31 | -0.09 | 0.64 | -0.07 | -0.40, 0.31 | 0.71 | -0.13 | 0.50 | -0.13 | -0.45, 0.20 | 0.46 |
| Δ TMS | 31 | -0.08 | 0.67 | -0.04 | -0.41, 0.29 | 0.81 | -0.07 | 0.71 | -0.02 | -0.39, 0.29 | 0.92 |
| Δ SDMT | 31 | 0.30 | 0.11 | 0.28 | -0.18, 0.66 | 0.20 | 0.29 | 0.11 | 0.27 | -0.17, 0.63 | 0.20 |
| Δ SCN | 31 | 0.01 | 0.96 | 0.03 | -0.27, 0.42 | 0.88 | 0.00 | 0.99 | -0.00 | -0.29, 0.36 | 0.99 |
| Δ VFC | 31 | 0.25 | 0.17 | 0.18 | -0.16, 0.46 | 0.27 | 0.25 | 0.17 | 0.17 | -0.14, 0.46 | 0.27 |
| Δ SWR | 31 | 0.32 | 0.08 | 0.27 | 0.02, 0.51 | **0.03** | 0.32 | 0.08 | 0.25 | -0.01, 0.49 | 0.05 |
| Δ Whole brain | 30 | 0.39 | **0.03** | 0.37 | 0.09, 0.62 | **0.007** | 0.30 | 0.10 | 0.31 | 0.04, 0.56 | **0.02** |
| Δ Caudate | 31 | 0.07 | 0.70 | 0.10 | -0.16, 0.39 | 0.46 | 0.05 | 0.77 | 0.06 | -0.18, 0.33 | 0.64 |
| Δ White matter | 30 | 0.20 | 0.29 | 0.20 | -0.10, 0.55 | 0.23 | 0.21 | 0.27 | 0.17 | -0.09, 0.44 | 0.22 |
| Δ Grey matter | 30 | 0.04 | 0.85 | 0.03 | -0.29, 0.43 | 0.86 | 0.01 | 0.98 | -0.03 | -0.31, 0.32 | 0.85 |

| Baseline tCre | | Age-adjusted | | | | | Age and CAG-adjusted | | | | |
|---|---|---|---|---|---|---|---|---|---|---|---|
| | | **Inverse weighted** | | **Bootstrapped** | | | **Inverse weighted** | | **Bootstrapped** | | |
| **Measures** | **n** | **r** | ***p* value** | **r** | **95 % CIs** | ***p* value** | **r** | ***p* value** | **r** | **95 % CIs** | ***p* value** |
| Δ cUHDRS | 31 | 0.40 | **0.03** | 0.36 | 0.06, 0.57 | **0.004** | 0.46 | **0.01** | 0.43 | 0.15, 0.60 | **<0.001** |
| Δ DBS | 31 | 0.05 | 0.80 | 0.06 | -0.24, 0.35 | 0.68 | -0.01 | 0.94 | 0.00 | -0.31, 0.27 | 1.00 |
| Δ TFC | 31 | 0.28 | 0.13 | 0.27 | -0.10, 0.57 | 0.11 | 0.34 | 0.06 | 0.33 | -0.04, 0.57 | **0.03** |
| Δ TMS | 31 | -0.25 | 0.17 | -0.25 | -0.47, 0.15 | 0.10 | -0.27 | 0.14 | -0.27 | -0.49, 0.22 | 0.10 |
| Δ SDMT | 31 | 0.20 | 0.27 | 0.20 | -0.13, 0.52 | 0.24 | 0.22 | 0.24 | 0.22 | -0.13, 0.52 | 0.20 |
| Δ SCN | 31 | 0.01 | 0.97 | 0.00 | -0.37, 0.38 | 0.98 | 0.02 | 0.92 | 0.02 | -0.37, 0.40 | 0.91 |
| Δ VFC | 31 | 0.04 | 0.83 | 0.01 | -0.36, 0.43 | 0.95 | 0.04 | 0.84 | 0.02 | -0.36, 0.43 | 0.94 |
| Δ SWR | 31 | 0.29 | 0.12 | 0.24 | -0.03, 0.47 | 0.07 | 0.30 | 0.10 | 0.26 | -0.01, 0.48 | **0.04** |
| Δ Whole brain | 30 | 0.37 | **0.04** | 0.34 | -0.10, 0.67 | 0.09 | 0.45 | **0.01** | 0.42 | 0.06, 0.75 | **0.02** |
| Δ Caudate | 31 | 0.33 | 0.07 | 0.28 | -0.02, 0.52 | 0.05 | 0.41 | **0.02** | 0.36 | 0.00, 0.55 | **0.005** |
| Δ White matter | 30 | 0.25 | 0.17 | 0.19 | -0.23, 0.59 | 0.35 | 0.38 | **0.04** | 0.30 | -0.11, 0.60 | 0.09 |
| Δ Grey matter | 30 | 0.09 | 0.65 | 0.03 | -0.35, 0.37 | 0.85 | 0.14 | 0.45 | 0.08 | -0.24, 0.41 | 0.63 |

| | | Age-adjusted | | | | | Age and CAG-adjusted | | | | |
|---|---|---|---|---|---|---|---|---|---|---|---|
| **Baseline tCho** | | **Inverse weighted** | | **Bootstrapped** | | | **Inverse weighted** | | **Bootstrapped** | | |
| **Measures** | **n** | **r** | ***p* value** | **r** | **95 % CIs** | ***p* value** | **r** | ***p* value** | **r** | **95 % CIs** | ***p* value** |
| Δ cUHDRS | 31 | 0.32 | 0.08 | 0.26 | -0.12, 0.54 | 0.13 | 0.48 | **0.01** | 0.43 | 0.10, 0.68 | **0.005** |
| Δ DBS | 31 | 0.34 | 0.19 | 0.26 | -0.08, 0.54 | 0.10 | -0.01 | 0.98 | 0.00 | -0.33, 0.33 | 1.00 |
| Δ TFC | 31 | 0.20 | 0.28 | 0.15 | -0.31, 0.50 | 0.49 | 0.37 | **0.04** | 0.31 | -0.13, 0.63 | 0.13 |
| Δ TMS | 31 | -0.30 | 0.10 | -0.22 | -0.57, 0.21 | 0.32 | -0.39 | **0.03** | -0.30 | -0.66, 0.15 | 0.17 |
| Δ SDMT | 31 | 0.10 | 0.59 | 0.09 | -0.24, 0.37 | 0.54 | 0.15 | 0.41 | 0.16 | -0.16, 0.47 | 0.34 |
| Δ SCN | 31 | -0.08 | 0.67 | -0.08 | -0.43, 0.26 | 0.66 | -0.02 | 0.91 | -0.00 | -0.33, 0.34 | 0.98 |
| Δ VFC | 31 | -0.14 | 0.46 | -0.17 | -0.44, 0.13 | 0.25 | -0.14 | 0.46 | -0.17 | -0.45, 0.15 | 0.28 |
| Δ SWR | 31 | 0.25 | 0.18 | 0.24 | -0.03, 0.44 | **0.04** | 0.30 | 0.10 | 0.32 | 0.03, 0.54 | **0.01** |
| Δ Whole brain | 30 | 0.13 | 0.50 | 0.08 | -0.37, 0.50 | 0.72 | 0.30 | 0.10 | 0.26 | -0.17, 0.63 | 0.21 |
| Δ Caudate | 31 | 0.24 | 0.19 | 0.17 | -0.17, 0.59 | 0.37 | 0.44 | **0.01** | 0.37 | 0.06, 0.70 | **0.03** |
| Δ White matter | 30 | -0.05 | 0.80 | -0.10 | -0.44, 0.31 | 0.62 | 0.17 | 0.38 | 0.11 | -0.24, 0.46 | 0.55 |
| Δ Grey matter | 30 | -0.04 | 0.85 | -0.08 | -0.46, 0.39 | 0.72 | 0.14 | 0.47 | 0.09 | -0.27, 0.50 | 0.66 |

| | | **Age-adjusted** | | | | | **Age and CAG-adjusted** | | | | |
|---|---|---|---|---|---|---|---|---|---|---|---|
| **Baseline MI** | | **Inverse weighted** | | **Bootstrapped** | | | **Inverse weighted** | | **Bootstrapped** | | |
| **Measures** | **n** | **r** | ***p* value** | **r** | **95 % CIs** | ***p* value** | **r** | ***p* value** | **r** | **95 % CIs** | ***p* value** |
| Δ cUHDRS | 31 | -0.12 | 0.52 | -0.10 | -0.35, 0.15 | 0.41 | 0.05 | 0.78 | 0.08 | -0.19, 0.31 | 0.52 |
| Δ DBS | 31 | 0.41 | **0.02** | 0.39 | 0.02, 0.64 | **0.01** | 0.01 | 0.94 | 0.00 | -0.32, 0.38 | 1.00 |
| Δ TFC | 31 | -0.17 | 0.35 | -0.14 | -0.50, 0.20 | 0.44 | 0.01 | 0.95 | 0.05 | -0.30, 0.36 | 0.77 |
| Δ TMS | 31 | 0.03 | 0.86 | -0.01 | -0.35, 0.26 | 0.93 | -0.08 | 0.68 | -0.12 | -0.42, 0.14 | 0.39 |
| Δ SDMT | 31 | -0.05 | 0.79 | -0.04 | -0.31, 0.23 | 0.74 | 0.04 | 0.85 | 0.04 | -0.26, 0.32 | 0.76 |
| Δ SCN | 31 | -0.25 | 0.17 | -0.26 | -0.59, 0.10 | 0.15 | -0.17 | 0.37 | -0.17 | -0.52, 0.18 | 0.35 |
| Δ VFC | 31 | -0.06 | 0.72 | -0.04 | -0.39, 0.31 | 0.83 | -0.05 | 0.80 | -0.03 | -0.38, 0.34 | 0.89 |
| Δ SWR | 31 | -0.05 | 0.77 | -0.09 | -0.35, 0.21 | 0.54 | 0.01 | 0.94 | 0.00 | -0.24, 0.24 | 0.97 |
| Δ Whole brain | 30 | -0.46 | **0.01** | -0.49 | -0.72, -0.08 | **0.002** | -0.30 | 0.10 | -0.36 | -0.65, 0.12 | 0.07 |
| Δ Caudate | 31 | -0.44 | **0.01** | -0.47 | -0.71, 0.19 | **<0.001** | -0.31 | 0.09 | -0.34 | -0.63, -0.04 | **0.03** |
| Δ White matter | 30 | -0.58 | **<0.001** | -0.58 | -0.78, -0.24 | **<0.001** | -0.44 | **0.02** | -0.44 | -0.70, -0.03 | **0.008** |
| Δ Grey matter | 30 | -0.49 | **0.01** | -0.48 | -0.68, -0.17 | **<0.001** | -0.35 | 0.06 | -0.34 | -0.61, 0.03 | **0.03** |

| Baseline GSH | | Age-adjusted | | | | | Age and CAG-adjusted | | | | |
|---|---|---|---|---|---|---|---|---|---|---|---|
| | | Inverse weighted | | Bootstrapped | | | Inverse weighted | | Bootstrapped | | |
| **Measures** | **n** | **r** | ***p* value** | **r** | **95 % CIs** | ***p* value** | **r** | ***p* value** | **r** | **95 % CIs** | ***p* value** |
| Δ cUHDRS | 31 | 0.23 | 0.22 | 0.25 | -0.07, 0.54 | 0.11 | 0.24 | 0.20 | 0.26 | -0.04, 0.52 | 0.08 |
| Δ DBS | 31 | 0.02 | 0.92 | -0.03 | -0.38, 0.25 | 0.86 | 0.05 | 0.80 | 0.00 | -0.36, 0.27 | 1.00 |
| Δ TFC | 31 | 0.05 | 0.79 | 0.08 | -0.23, 0.39 | 0.60 | 0.05 | 0.80 | 0.08 | -0.21, 0.39 | 0.61 |
| Δ TMS | 31 | -0.04 | 0.85 | -0.08 | -0.33, 0.26 | 0.58 | -0.03 | 0.85 | -0.07 | -0.34, 0.30 | 0.62 |
| Δ SDMT | 31 | 0.26 | 0.15 | 0.27 | -0.02, 0.58 | 0.10 | 0.26 | 0.15 | 0.27 | -0.04, 0.58 | 0.10 |
| Δ SCN | 31 | 0.18 | 0.33 | 0.16 | -0.25, 0.51 | 0.43 | 0.18 | 0.32 | 0.16 | -0.24, 0.51 | 0.43 |
| Δ VFC | 31 | 0.14 | 0.45 | 0.17 | -0.15, 0.48 | 0.31 | 0.14 | 0.45 | 0.17 | -0.15, 0.48 | 0.31 |
| Δ SWR | 31 | 0.24 | 0.20 | 0.21 | -0.24, 0.51 | 0.27 | 0.24 | 0.19 | 0.20 | -0.24, 0.50 | 0.27 |
| Δ Whole brain | 30 | 0.38 | **0.04** | 0.36 | -0.04, 0.65 | **0.04** | 0.40 | **0.03** | 0.38 | -0.02, 0.68 | **0.04** |
| Δ Caudate | 31 | 0.18 | 0.32 | 0.17 | -0.15, 0.45 | 0.25 | 0.20 | 0.27 | 0.18 | -0.17, 0.43 | 0.22 |
| Δ White matter | 30 | 0.26 | 0.17 | 0.24 | -0.12, 0.57 | 0.17 | 0.32 | 0.09 | 0.28 | -0.07, 0.58 | 0.10 |
| Δ Grey matter | 30 | 0.10 | 0.59 | 0.09 | -0.22, 0.43 | 0.58 | 0.11 | 0.57 | 0.08 | -0.22, 0.41 | 0.59 |

| | | Age-adjusted | | | | | Age and CAG-adjusted | | | | |
|---|---|---|---|---|---|---|---|---|---|---|---|
| **Baseline GABA** | | **Inverse weighted** | | **Bootstrapped** | | | **Inverse weighted** | | **Bootstrapped** | | |
| **Measures** | **n** | **r** | ***p* value** | **r** | **95 % CIs** | ***p* value** | **r** | ***p* value** | **r** | **95 % CIs** | ***p* value** |
| Δ cUHDRS | 22 | -0.05 | 0.81 | -0.26 | -0.66, 0.37 | 0.27 | -0.05 | 0.81 | -0.28 | -0.67, 0.33 | 0.22 |
| Δ DBS | 22 | 0.02 | 0.94 | -0.03 | -0.51, 0.32 | 0.90 | 0.04 | 0.85 | 0.00 | -0.49, 0.35 | 1.00 |
| Δ TFC | 22 | -0.18 | 0.44 | -0.23 | -0.74, 0.19 | 0.34 | -0.18 | 0.43 | -0.26 | -0.73, 0.15 | 0.26 |
| Δ TMS | 22 | -0.01 | 0.95 | 0.17 | -0.25, 0.74 | 0.47 | -0.02 | 0.94 | 0.18 | -0.26, 0.70 | 0.43 |
| Δ SDMT | 22 | -0.21 | 0.35 | -0.33 | -0.57, 0.02 | **0.02** | -0.21 | 0.36 | -0.33 | -0.57, 0.04 | **0.03** |
| Δ SCN | 22 | -0.24 | 0.28 | -0.24 | -0.57, 0.14 | 0.19 | -0.25 | 0.26 | -0.26 | -0.59, 0.13 | 0.17 |
| Δ VFC | 22 | 0.02 | 0.92 | -0.12 | -0.49, 0.36 | 0.57 | 0.02 | 0.93 | -0.12 | -0.48, 0.39 | 0.59 |
| Δ SWR | 22 | 0.22 | 0.33 | 0.07 | -0.52, 0.59 | 0.82 | 0.22 | 0.32 | 0.06 | -0.55, 0.59 | 0.83 |
| Δ Whole brain | 21 | 0.11 | 0.65 | 0.10 | -0.35, 0.38 | 0.54 | 0.07 | 0.76 | 0.08 | -0.23, 0.35 | 0.61 |
| Δ Caudate | 22 | -0.04 | 0.86 | -0.07 | -0.38, 0.26 | 0.69 | -0.04 | 0.85 | -0.09 | -0.43, 0.25 | 0.58 |
| Δ White matter | 22 | 0.01 | 0.97 | -0.02 | -0.57, 0.32 | 0.93 | 0.02 | 0.93 | -0.04 | -0.42, 0.27 | 0.82 |
| Δ Grey matter | 22 | -0.16 | 0.48 | -0.17 | -0.57, 0.22 | 0.39 | -0.17 | 0.45 | -0.20 | -0.53, 0.16 | 0.26 |

| | | Age-adjusted | | | | | Age and CAG-adjusted | | | | |
|---|---|---|---|---|---|---|---|---|---|---|---|
| **Baseline GLX** | | **Inverse weighted** | | **Bootstrapped** | | | **Inverse weighted** | | **Bootstrapped** | | |
| **Measures** | **n** | **r** | ***p* value** | **r** | **95 % CIs** | ***p* value** | **r** | ***p* value** | **r** | **95 % CIs** | ***p* value** |
| Δ cUHDRS | 31 | -0.05 | 0.77 | 0.01 | -0.37, 0.47 | 0.96 | -0.06 | 0.74 | -0.01 | -0.45, 0.40 | 0.95 |
| Δ DBS | 31 | 0.03 | 0.86 | -0.05 | -0.37, 0.35 | 0.78 | 0.08 | 0.66 | 0.00 | -0.32, 0.40 | 1.00 |
| Δ TFC | 31 | 0.00 | 0.99 | 0.06 | -0.21, 0.37 | 0.70 | 0.00 | 0.99 | 0.04 | -0.20, 0.30 | 0.79 |
| Δ TMS | 31 | -0.03 | 0.88 | -0.08 | -0.35, 0.22 | 0.61 | -0.02 | 0.90 | -0.06 | -0.37, 0.22 | 0.66 |
| Δ SDMT | 31 | -0.07 | 0.70 | 0.02 | -0.39, 0.59 | 0.92 | -0.07 | 0.70 | 0.01 | -0.38, 0.58 | 0.95 |
| Δ SCN | 31 | -0.27 | 0.14 | -0.20 | -0.61, 0.38 | 0.41 | -0.28 | 0.12 | -0.23 | -0.63, 0.33 | 0.36 |
| Δ VFC | 31 | -0.20 | 0.28 | -0.11 | -0.43, 0.33 | 0.57 | -0.20 | 0.29 | -0.11 | -0.43, 0.31 | 0.55 |
| Δ SWR | 31 | -0.10 | 0.58 | -0.11 | -0.60, 0.36 | 0.66 | -0.11 | 0.57 | -0.13 | -0.61, 0.34 | 0.61 |
| Δ Whole brain | 30 | 0.24 | 0.21 | 0.25 | -0.09, 0.53 | 0.13 | 0.28 | 0.13 | 0.26 | -0.08, 0.55 | 0.11 |
| Δ Caudate | 31 | 0.00 | 0.99 | 0.03 | -0.25, 0.33 | 0.86 | -0.01 | 0.96 | 0.00 | -0.40, 0.31 | 0.99 |
| Δ White matter | 30 | 0.16 | 0.40 | 0.20 | -0.11, 0.51 | 0.20 | 0.20 | 0.30 | 0.20 | -0.08, 0.45 | 0.12 |
| Δ Grey matter | 30 | 0.04 | 0.85 | 0.09 | -0.27, 0.40 | 0.60 | 0.04 | 0.85 | 0.06 | -0.26, 0.35 | 0.67 |

*Relationships between baseline MRS metabolites and rate of change (Δ) in clinical, cognitive and imaging markers were assessed using Pearson's partial correlation controlling for age, and age and CAG repeat length. Correlation coefficients and 95% confidence intervals were computed using bootstrap testing with 1000 repetitions. A weighted correlation was also conducted, applying an inverse weighting to %SD value. Results displayed are unadjusted for multiplicity. Bold text indicated significance p < 0.05. cUHDRS, composite Unified Huntington's Disease Rating Scale; DBS, Disease Burden Score; TFC, Total Functional Capacity; TMS, Total Motor Score; SDMT, Symbol Digit Modalities Test; SCN, Stroop Colour Naming; VFC, Verbal Fluency Categorical; SWR, Stroop Word Reading Test; NfL, Neurofilament Light Chain; mHtt, Mutant Huntingtin.*

**A) tNAA**

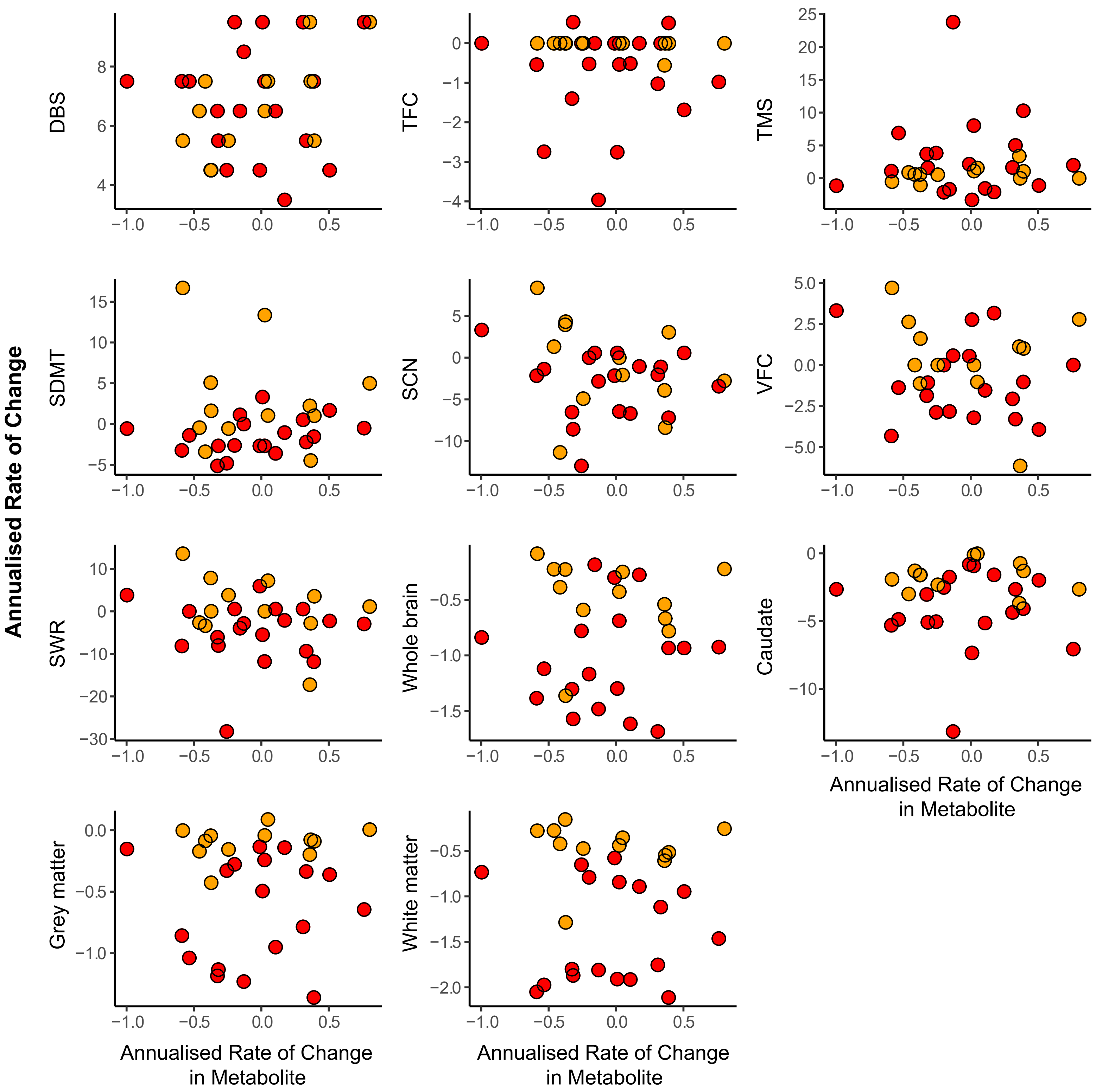
Annualised Rate of Change
DBS
TFC
TMS
SDMT
SCN
VFC
SWR
Whole brain
Caudate
Grey matter
White matter
Annualised Rate of Change in Metabolite

**B) tCre**

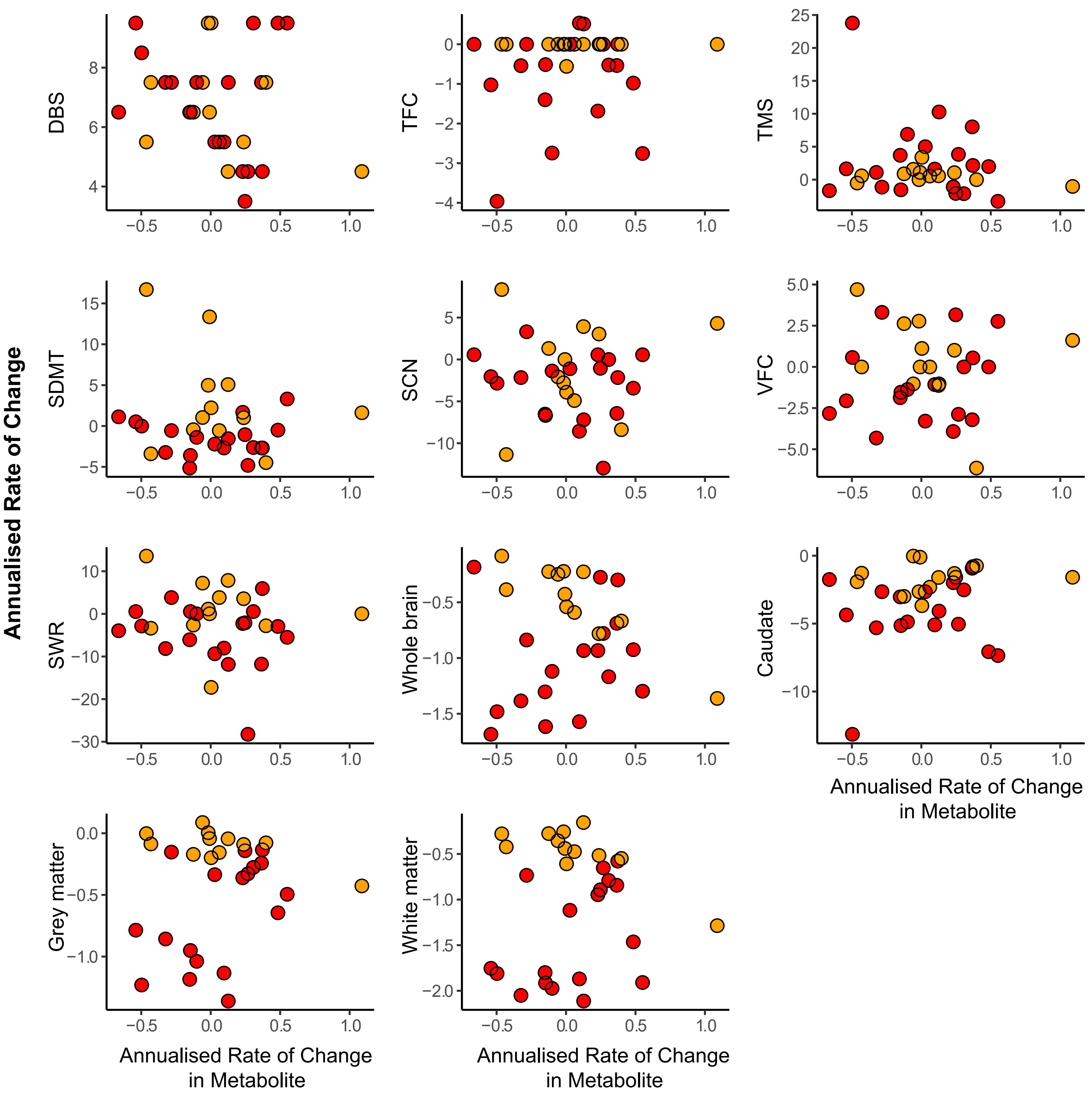

Annualised Rate of Change
DBS
TFC
TMS
SDMT
SCN
VFC
SWR
Whole brain
Caudate
Grey matter
White matter
Annualised Rate of Change in Metabolite

C) tCho

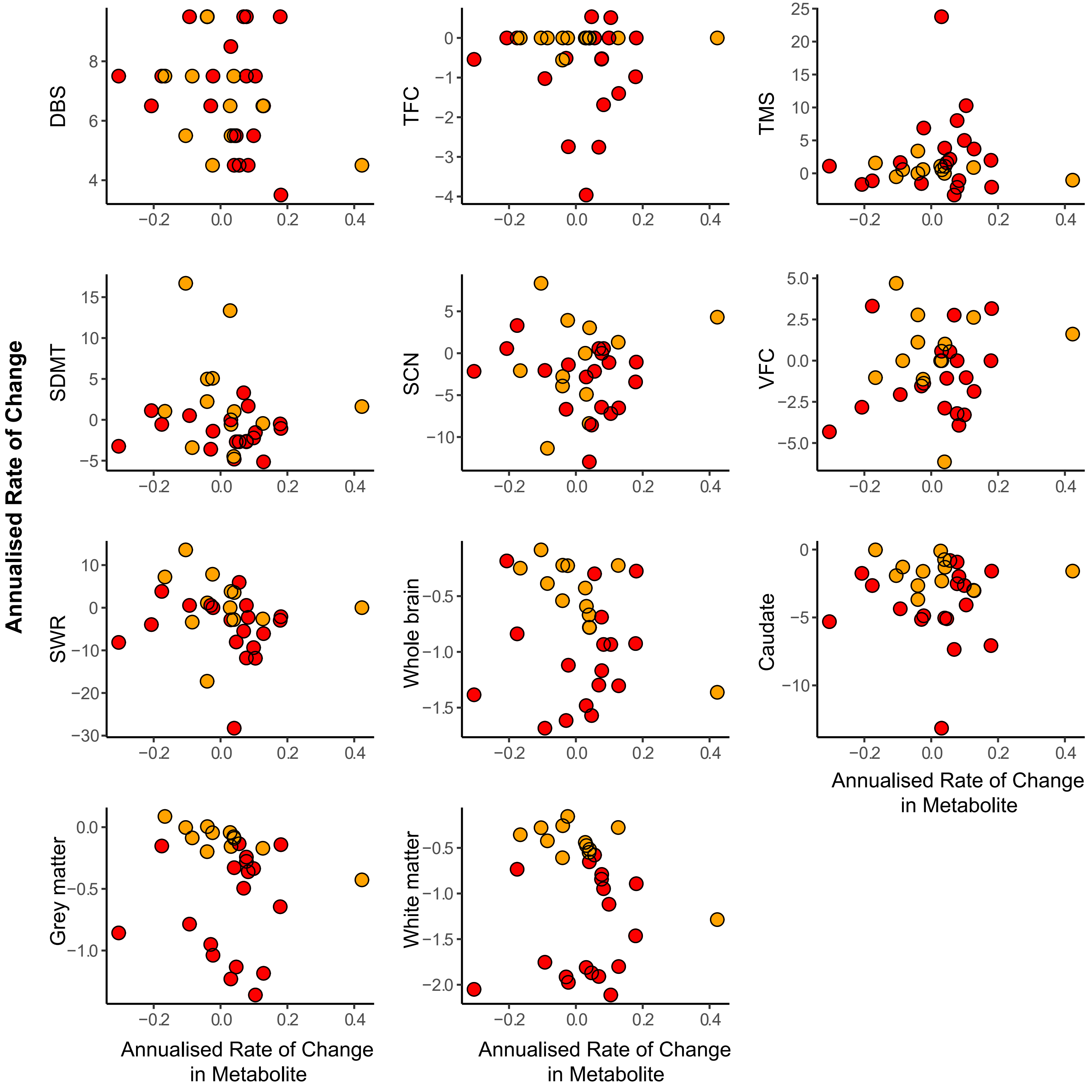

Annualised Rate of Change
DBS
TFC
TMS
SDMT
SCN
VFC
SWR
Whole brain
Caudate
Grey matter
White matter
Annualised Rate of Change in Metabolite

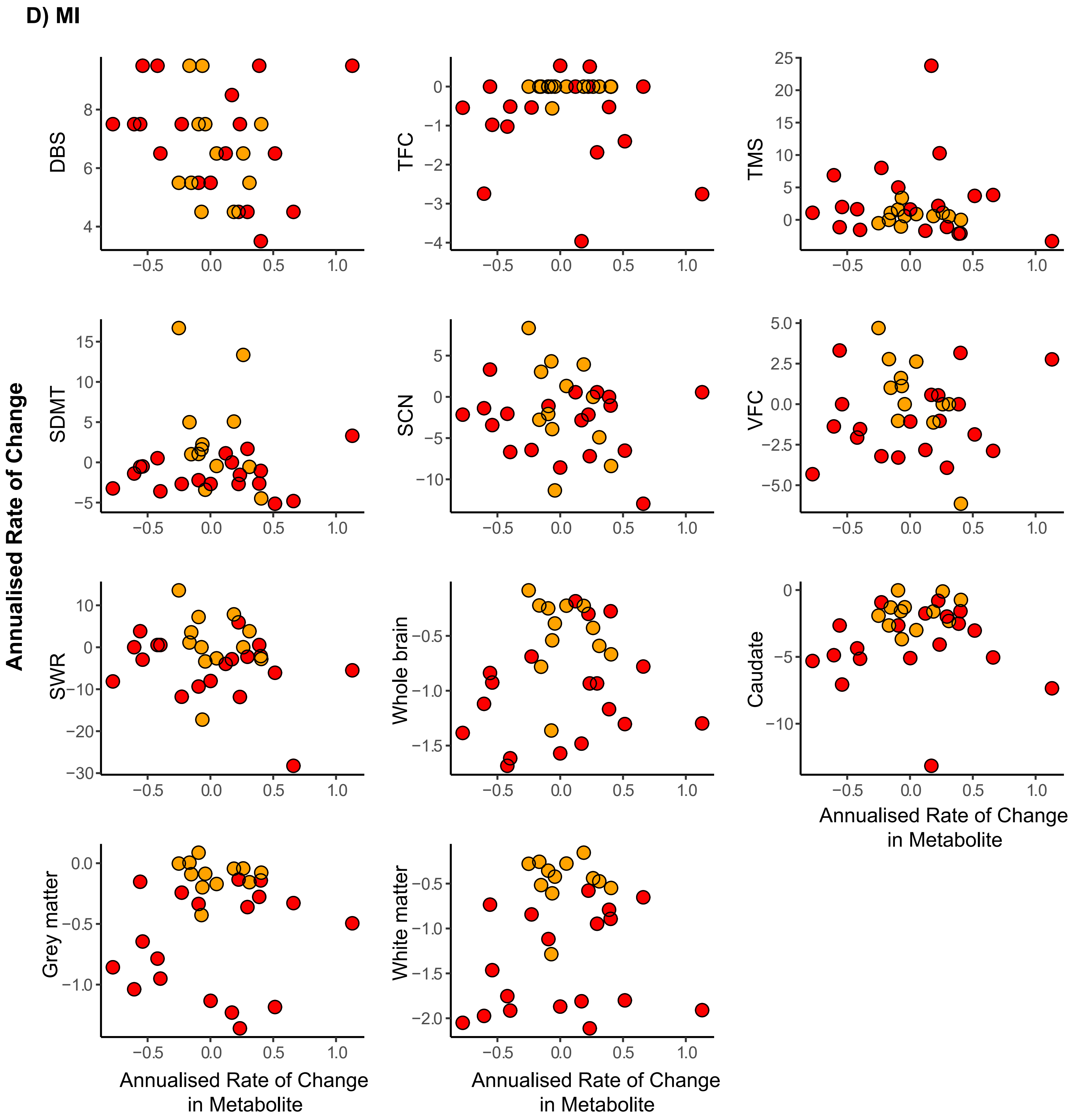
D) MI
Annualised Rate of Change
DBS
TFC
TMS
SDMT
SCN
VFC
SWR
Whole brain
Caudate
Grey matter
White matter
Annualised Rate of Change in Metabolite

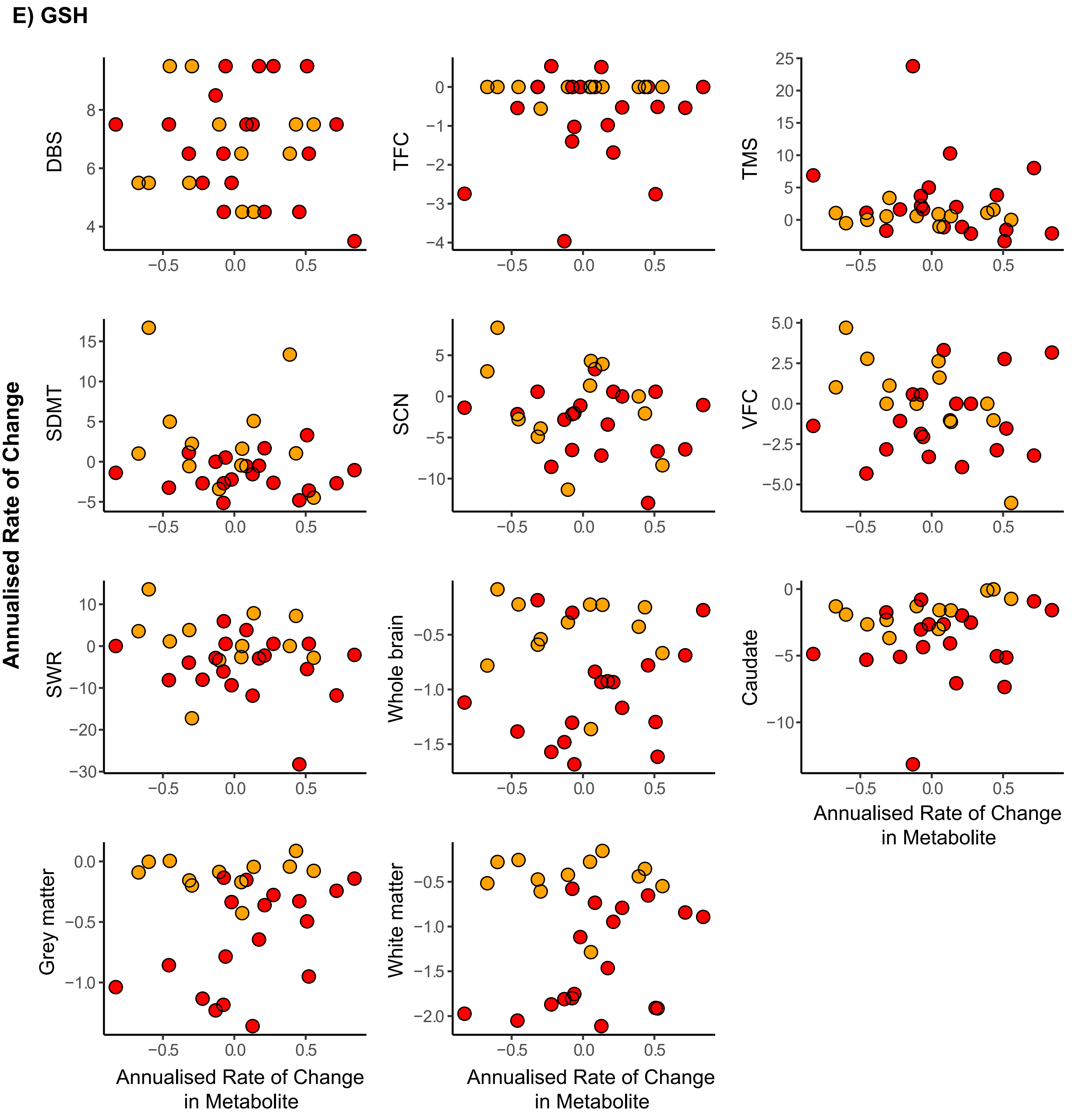
E) GSH
Annualised Rate of Change
DBS
TFC
TMS
SDMT
SCN
VFC
SWR
Whole brain
Caudate
Grey matter
White matter
Annualised Rate of Change in Metabolite
-0.5
0.0
0.5

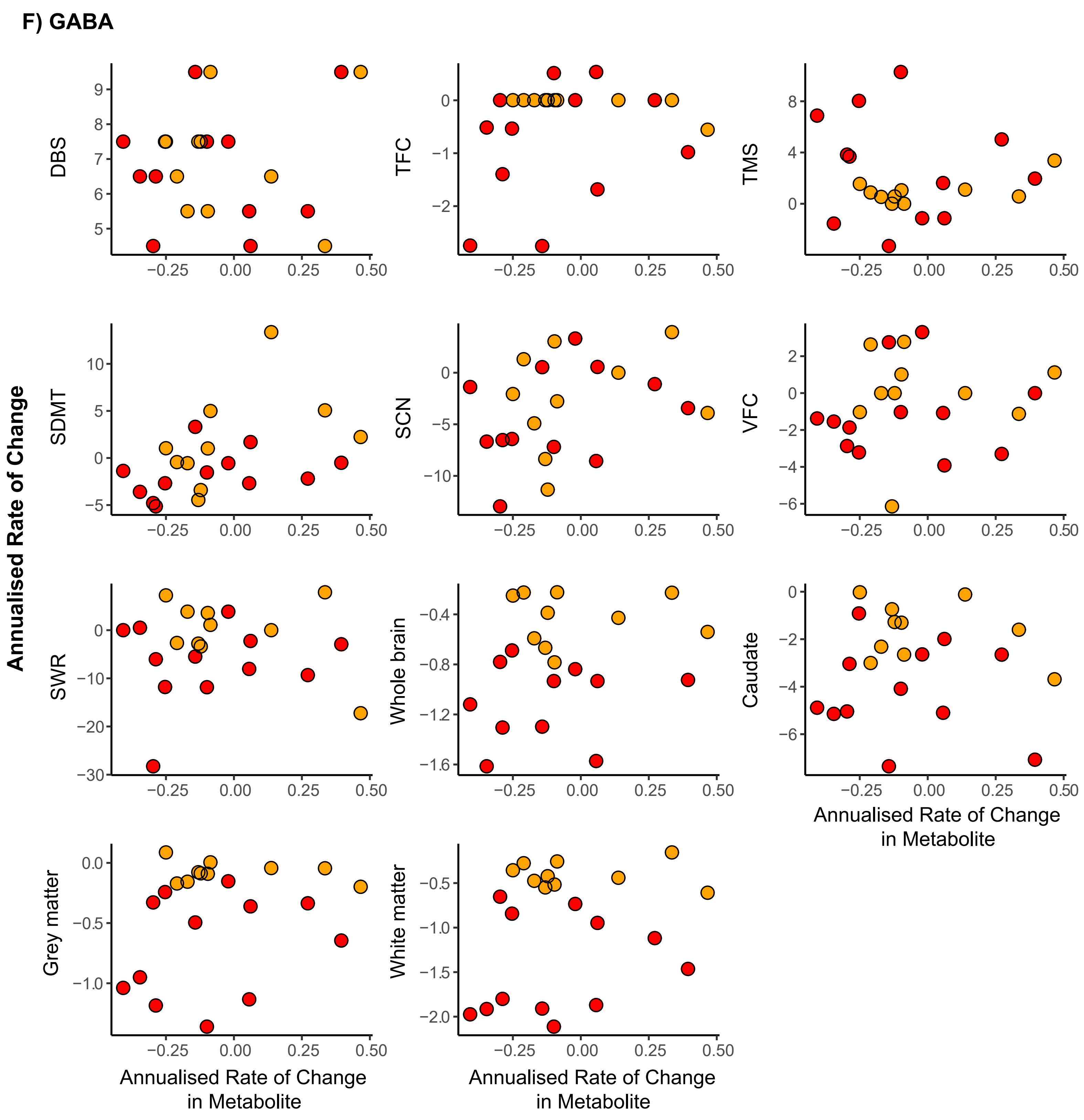
F) GABA
Annualised Rate of Change
DBS
TFC
TMS
SDMT
SCN
VFC
SWR
Whole brain
Caudate
Grey matter
White matter
Annualised Rate of Change in Metabolite
−0.25 0.00 0.25 0.50

### G) GLX

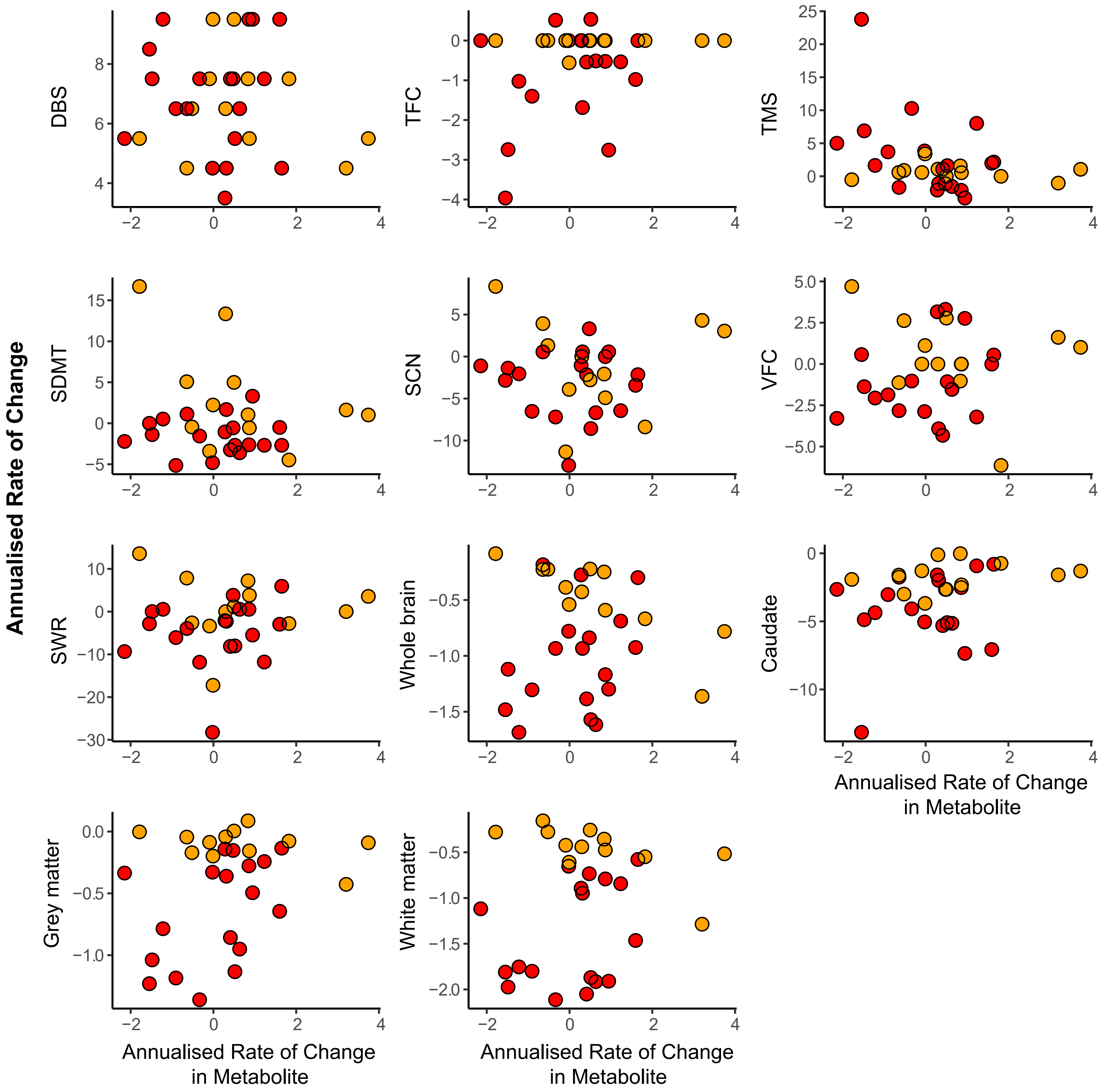


**Supplementary Fig. 6: Scatterplots of Correlations Between Annualised Rate of Change in Metabolites and Established Measures**. Scatter plots displaying associations between annualised rate of change in tNAA (**A**), tCre (**B**), tCho (**C**), MI (**D**), GSH (**E**), GABA (**F**), GLX (**G**) and measures of clinical progression, cognitive decline, imaging markers. Values displayed are controlled for CSF PVE only. Red and yellow datapoints indicate manifest and premanifest patients, respectively. cUHDRS, Composite Unified Huntingtons Disease Rating Scale; DBS, Disease Burden Score; TFC, Total Functional Capacity; TMS, Total Motor Score; SDMT, Symbol Digit Modalities Test; SCN, Stroop Colour Naming; VFC, Verbal Fluency Categorical; SWR, Stroop Word Reading Test.

**Supplementary Table 7 – Longitudinal correlations between annualised rate of change (Δ) of each metabolite and the Δ in clinical, cognitive and imaging measures.**

| | | Age-adjusted | | | | | Age and CAG-adjusted | | | | |
|---|---|---|---|---|---|---|---|---|---|---|---|
| Δ tNAA | | Inverse weighted | | Bootstrapped | | | Inverse weighted | | Bootstrapped | | |
| **Measures** | **n** | **r** | ***p* value** | **r** | **95 % CIs** | ***p* value** | **r** | ***p* value** | **r** | **95 % CIs** | ***p* value** |
| Δ cUHDRS | 31 | -0.05 | 0.79 | -0.07 | -0.41, 0.20 | 0.66 | 0.04 | 0.83 | 0.06 | -0.24, 0.34 | 0.69 |
| Δ DBS | 31 | 0.21 | 0.26 | 0.26 | -0.09, 0.58 | 0.13 | -0.05 | 0.78 | 0.00 | -0.33, 0.37 | 1.00 |
| Δ TFC | 31 | 0.01 | 0.97 | -0.00 | -0.29, 0.27 | 0.99 | 0.12 | 0.51 | 0.14 | -0.28, 0.42 | 0.43 |
| Δ TMS | 31 | 0.09 | 0.65 | 0.04 | -0.21, 0.32 | 0.75 | 0.05 | 0.80 | -0.02 | -0.27, 0.28 | 0.86 |
| Δ SDMT | 31 | 0.06 | 0.73 | 0.01 | -0.41, 0.33 | 0.94 | 0.11 | 0.57 | 0.08 | -0.32, 0.37 | 0.66 |
| Δ SCN | 31 | -0.14 | 0.45 | -0.19 | -0.47, 0.18 | 0.28 | -0.10 | 0.60 | -0.12 | -0.39, 0.27 | 0.48 |
| Δ VFC | 31 | -0.17 | 0.35 | -0.18 | -0.55, 0.25 | 0.39 | -0.18 | 0.33 | -0.17 | -0.54, 0.26 | 0.40 |
| Δ SWR | 31 | -0.12 | 0.53 | -0.15 | -0.44, 0.19 | 0.35 | -0.09 | 0.64 | -0.09 | -0.35, 0.18 | 0.51 |
| Δ Whole brain | 30 | 0.12 | 0.54 | 0.06 | -0.22, 0.33 | 0.65 | 0.30 | 0.10 | 0.28 | -0.03, 0.57 | 0.07 |
| Δ Caudate | 31 | 0.01 | 0.96 | -0.00 | -0.37, 0.22 | 0.99 | 0.14 | 0.44 | 0.17 | -0.10, 0.35 | 0.13 |
| Δ White matter | 30 | 0.05 | 0.79 | 0.04 | -0.22, 0.38 | 0.80 | 0.26 | 0.17 | 0.29 | -0.05, 0.58 | 0.07 |
| Δ Grey matter | 30 | 0.08 | 0.69 | 0.08 | -0.24, 0.36 | 0.58 | 0.24 | 0.19 | 0.28 | -0.06, 0.55 | 0.07 |

| | | Age-adjusted | | | | | Age and CAG-adjusted | | | | |
|---|---|---|---|---|---|---|---|---|---|---|---|
| Δ tCre | | **Inverse weighted** | | **Bootstrapped** | | | **Inverse weighted** | | **Bootstrapped** | | |
| **Measures** | **n** | **r** | ***p* value** | **r** | **95 % CIs** | ***p* value** | **r** | ***p* value** | **r** | **95 % CIs** | ***p* value** |
| Δ cUHDRS | 31 | 0.13 | 0.49 | 0.06 | -0.32, 0.50 | 0.78 | 0.08 | 0.65 | 0.01 | -0.34, 0.43 | 0.98 |
| Δ DBS | 31 | -0.15 | 0.42 | -0.12 | -0.50, 0.27 | 0.54 | -0.03 | 0.87 | 0.00 | -0.43, 0.37 | 1.00 |
| Δ TFC | 31 | 0.14 | 0.44 | 0.12 | -0.31, 0.47 | 0.56 | 0.09 | 0.62 | 0.07 | -0.28, 0.45 | 0.71 |
| Δ TMS | 31 | -0.28 | 0.12 | -0.23 | -0.59, 0.12 | 0.22 | -0.26 | 0.15 | -0.21 | -0.56, 0.18 | 0.28 |
| Δ SDMT | 31 | 0.01 | 0.97 | -0.07 | -0.42, 0.31 | 0.74 | -0.02 | 0.91 | -0.09 | -0.49, 0.27 | 0.63 |
| Δ SCN | 31 | 0.13 | 0.50 | 0.06 | -0.36, 0.50 | 0.78 | 0.10 | 0.60 | 0.03 | -0.41, 0.46 | 0.90 |
| Δ VFC | 31 | 0.13 | 0.49 | 0.07 | -0.37, 0.44 | 0.75 | 0.13 | 0.50 | 0.06 | -0.39, 0.44 | 0.77 |
| Δ SWR | 31 | -0.07 | 0.72 | -0.09 | -0.35, 0.18 | 0.51 | -0.09 | 0.63 | -0.13 | -0.39, 0.14 | 0.34 |
| Δ Whole brain | 30 | 0.11 | 0.58 | 0.01 | -0.47, 0.42 | 0.97 | 0.03 | 0.86 | -0.07 | -0.54, 0.33 | 0.76 |
| Δ Caudate | 31 | 0.23 | 0.21 | 0.20 | -0.23, 0.55 | 0.33 | 0.19 | 0.30 | 0.16 | -0.22, 0.51 | 0.42 |
| Δ White matter | 30 | 0.29 | 0.12 | 0.25 | -0.27, 0.58 | 0.23 | 0.26 | 0.16 | 0.22 | -0.19, 0.56 | 0.27 |
| Δ Grey matter | 30 | 0.43 | **0.02** | 0.39 | 0.05, 0.64 | **0.01** | 0.42 | **0.02** | 0.38 | 0.09, 0.63 | **0.006** |

| | | Age-adjusted | | | | | Age and CAG-adjusted | | | | |
|---|---|---|---|---|---|---|---|---|---|---|---|
| Δ tCho | | Inverse weighted | | Bootstrapped | | | Inverse weighted | | Bootstrapped | | |
| **Measures** | **n** | **r** | ***p* value** | **r** | **95 % CIs** | ***p* value** | **r** | ***p* value** | **r** | **95 % CIs** | ***p* value** |
| Δ cUHDRS | 31 | -0.00 | 0.99 | -0.04 | -0.29, 0.36 | 0.79 | -0.05 | 0.77 | -0.10 | -0.39, 0.23 | 0.51 |
| Δ DBS | 31 | -0.13 | 0.48 | -0.11 | -0.41, 0.20 | 0.47 | -0.02 | 0.90 | 0.00 | -0.34, 0.32 | 1.00 |
| Δ TFC | 31 | -0.04 | 0.82 | -0.03 | -0.28, 0.22 | 0.80 | -0.10 | 0.58 | -0.09 | -0.32, 0.13 | 0.41 |
| Δ TMS | 31 | 0.06 | 0.75 | 0.05 | -0.20, 0.29 | 0.69 | 0.09 | 0.65 | 0.08 | -0.18, 0.36 | 0.54 |
| Δ SDMT | 31 | 0.10 | 0.58 | 0.03 | -0.28, 0.40 | 0.86 | 0.08 | 0.66 | 0.00 | -0.34, 0.33 | 0.98 |
| Δ SCN | 31 | 0.10 | 0.58 | 0.11 | -0.21, 0.53 | 0.57 | 0.08 | 0.67 | 0.08 | -0.24, 0.50 | 0.68 |
| Δ VFC | 31 | 0.31 | 0.09 | 0.26 | -0.16, 0.60 | 0.18 | 0.31 | 0.09 | 0.26 | -0.15, 0.61 | 0.18 |
| Δ SWR | 31 | -0.02 | 0.93 | -0.05 | -0.31, 0.22 | 0.68 | -0.04 | 0.84 | -0.08 | -0.38, 0.17 | 0.55 |
| Δ Whole brain | 30 | 0.11 | 0.57 | 0.01 | -0.41, 0.39 | 0.97 | 0.06 | 0.74 | -0.06 | -0.51, 0.39 | 0.79 |
| Δ Caudate | 31 | 0.08 | 0.68 | 0.03 | -0.29, 0.34 | 0.85 | 0.02 | 0.90 | -0.04 | -0.30, 0.27 | 0.81 |
| Δ White matter | 30 | 0.30 | 0.10 | 0.24 | -0.10, 0.64 | 0.22 | 0.30 | 0.11 | 0.20 | -0.15, 0.69 | 0.34 |
| Δ Grey matter | 30 | 0.24 | 0.20 | 0.19 | -0.10, 0.51 | 0.21 | 0.21 | 0.26 | 0.15 | -0.15, 0.50 | 0.36 |

| | | Age-adjusted | | | | | Age and CAG-adjusted | | | | |
|---|---|---|---|---|---|---|---|---|---|---|---|
| Δ MI | | **Inverse weighted** | | **Bootstrapped** | | | **Inverse weighted** | | **Bootstrapped** | | |
| **Measures** | **n** | **r** | ***p* value** | **r** | **95 % CIs** | ***p* value** | **r** | ***p* value** | **r** | **95 % CIs** | ***p* value** |
| Δ cUHDRS | 31 | -0.05 | 0.77 | -0.13 | -0.37, 0.22 | 0.40 | -0.19 | 0.30 | -0.24 | -0.52, 0.08 | 0.11 |
| Δ DBS | 31 | -0.23 | 0.22 | -0.19 | -0.53, 0.30 | 0.35 | -0.04 | 0.83 | 0.00 | -0.35, 0.43 | 1.00 |
| Δ TFC | 31 | 0.08 | 0.66 | -0.10 | -0.57, 0.34 | 0.68 | -0.05 | 0.81 | -0.21 | -0.59, 0.17 | 0.30 |
| Δ TMS | 31 | -0.08 | 0.66 | -0.06 | -0.42, 0.22 | 0.69 | -0.02 | 0.92 | -0.02 | -0.49, 0.31 | 0.92 |
| Δ SDMT | 31 | -0.01 | 0.94 | -0.02 | -0.32, 0.31 | 0.90 | -0.08 | 0.68 | -0.07 | -0.45, 0.29 | 0.72 |
| Δ SCN | 31 | -0.23 | 0.20 | -0.18 | -0.53, 0.10 | 0.25 | -0.33 | 0.07 | -0.25 | -0.60, 0.14 | 0.18 |
| Δ VFC | 31 | 0.01 | 0.94 | 0.03 | -0.39, 0.44 | 0.90 | -0.01 | 0.96 | 0.02 | -0.42, 0.47 | 0.93 |
| Δ SWR | 31 | -0.27 | 0.15 | -0.24 | -0.53, 0.09 | 0.15 | -0.33 | 0.07 | -0.30 | -0.55, 0.03 | 0.05 |
| Δ Whole brain | 30 | 0.21 | 0.28 | 0.13 | -0.33, 0.49 | 0.54 | 0.07 | 0.69 | 0.02 | -0.33, 0.39 | 0.92 |
| Δ Caudate | 31 | 0.09 | 0.62 | 0.02 | -0.38, 0.48 | 0.93 | -0.05 | 0.80 | -0.10 | -0.41, 0.29 | 0.59 |
| Δ White matter | 30 | 0.29 | 0.12 | 0.15 | -0.47, 0.63 | 0.59 | 0.16 | 0.39 | 0.03 | -0.46, 0.49 | 0.92 |
| Δ Grey matter | 30 | 0.22 | 0.25 | 0.13 | -0.19, 0.52 | 0.49 | 0.09 | 0.64 | 0.02 | -0.33, 0.34 | 0.90 |

| Δ GSH | | Age-adjusted | | | | | Age and CAG-adjusted | | | | |
|---|---|---|---|---|---|---|---|---|---|---|---|
| | | **Inverse weighted** | | **Bootstrapped** | | | **Inverse weighted** | | **Bootstrapped** | | |
| **Measures** | **n** | **r** | ***p* value** | **r** | **95 % CIs** | ***p* value** | **r** | ***p* value** | **r** | **95 % CIs** | ***p* value** |
| Δ cUHDRS | 31 | -0.03 | 0.86 | -0.10 | -0.48, 0.30 | 0.63 | -0.07 | 0.72 | -0.14 | -0.48, 0.21 | 0.42 |
| Δ DBS | 31 | 0.01 | 0.98 | -0.07 | 0.42, 0.25 | 0.67 | 0.07 | 0.72 | 0.00 | -0.37, 0.31 | 1.00 |
| Δ TFC | 31 | 0.24 | 0.19 | 0.09 | -0.27, 0.48 | 0.65 | 0.24 | 0.19 | 0.06 | -0.25, 0.43 | 0.74 |
| Δ TMS | 31 | -0.19 | 0.30 | -0.13 | -0.39, 0.26 | 0.39 | -0.18 | 0.33 | -0.12 | -0.37, 0.22 | 0.41 |
| Δ SDMT | 31 | -0.19 | 0.30 | -0.20 | -0.54, 0.20 | 0.29 | -0.21 | 0.25 | -0.22 | -0.56, 0.17 | 0.25 |
| Δ SCN | 31 | -0.35 | 0.06 | -0.26 | -0.54, 0.08 | 0.11 | -0.37 | **0.04** | -0.29 | -0.56, 0.02 | 0.06 |
| Δ VFC | 31 | -0.11 | 0.53 | -0.16 | -0.51, 0.30 | 0.46 | -0.12 | 0.51 | -0.16 | -0.52, 0.29 | 0.44 |
| Δ SWR | 31 | 0.34 | 0.06 | -0.23 | -0.51, 0.20 | 0.21 | -0.35 | 0.05 | -0.25 | -0.50, 0.16 | 0.14 |
| Δ Whole brain | 30 | -0.05 | 0.81 | 0.01 | -0.34, 0.44 | 0.95 | -0.09 | 0.65 | -0.04 | -0.37, 0.31 | 0.84 |
| Δ Caudate | 31 | 0.06 | 0.73 | 0.12 | -0.22, 0.41 | 0.44 | 0.04 | 0.84 | 0.10 | -0.21, 0.40 | 0.52 |
| Δ White matter | 30 | 0.10 | 0.59 | 0.10 | -0.32, 0.49 | 0.64 | 0.08 | 0.67 | 0.07 | -0.32, 0.42 | 0.73 |
| Δ Grey matter | 30 | 0.19 | 0.32 | 0.21 | -0.16, 0.54 | 0.24 | 0.18 | 0.33 | 0.21 | -0.13, 0.49 | 0.20 |

| | | **Age-adjusted** | | | | | **Age and CAG-adjusted** | | | | |
|---|---|---|---|---|---|---|---|---|---|---|---|
| Δ GABA | | **Inverse weighted** | | **Bootstrapped** | | | **Inverse weighted** | | **Bootstrapped** | | |
| **Measures** | **n** | **r** | ***p* value** | **r** | **95 % CIs** | ***p* value** | **r** | ***p* value** | **r** | **95 % CIs** | ***p* value** |
| Δ cUHDRS | 22 | 0.37 | 0.09 | 0.30 | -0.35, 0.66 | 0.19 | 0.37 | 0.09 | 0.32 | -0.31, 0.63 | 0.14 |
| Δ DBS | 22 | -0.07 | 0.74 | 0.00 | -0.49, 0.51 | 0.99 | -0.08 | 0.74 | 0.00 | -0.50, 0.50 | 1.00 |
| Δ TFC | 22 | 0.25 | 0.27 | 0.20 | -0.25, 0.54 | 0.32 | 0.25 | 0.26 | 0.21 | -0.13, 0.57 | 0.22 |
| Δ TMS | 22 | -0.20 | 0.38 | -0.16 | -0.50, 0.30 | 0.42 | -0.19 | 0.39 | -0.16 | -0.50, 0.32 | 0.41 |
| Δ SDMT | 22 | 0.40 | 0.07 | 0.39 | 0.10, 0.59 | **0.001** | 0.40 | 0.07 | 0.39 | 0.09, 0.59 | **0.001** |
| Δ SCN | 22 | 0.30 | 0.17 | 0.30 | -0.05, 0.58 | 0.06 | 0.29 | 0.19 | 0.31 | -0.02, 0.57 | **0.04** |
| Δ VFC | 22 | 0.09 | 0.68 | 0.08 | -0.27, 0.39 | 0.62 | 0.11 | 0.62 | 0.09 | -0.26, 0.38 | 0.59 |
| Δ SWR | 22 | 0.02 | 0.91 | -0.00 | -0.63, 0.46 | 0.99 | 0.02 | 0.94 | -0.00 | -0.63, 0.46 | 0.99 |
| Δ Whole brain | 21 | -0.01 | 0.97 | -0.05 | -0.38, 0.30 | 0.79 | -0.00 | 1.00 | -0.04 | -0.41, 0.35 | 0.85 |
| Δ Caudate | 22 | -0.02 | 0.93 | -0.11 | -0.58, 0.32 | 0.65 | -0.05 | -0.82 | -0.12 | -0.54, 0.28 | 0.57 |
| Δ White matter | 22 | 0.12 | 0.61 | 0.06 | -0.34, 0.35 | 0.74 | 0.12 | 0.61 | 0.07 | -0.34, 0.41 | 0.71 |
| Δ Grey matter | 22 | 0.17 | 0.44 | 0.11 | -0.29, 0.42 | 0.52 | 0.17 | 0.44 | 0.12 | -0.31, 0.47 | 0.52 |

| | | Age-adjusted | | | | | Age and CAG-adjusted | | | | |
|---|---|---|---|---|---|---|---|---|---|---|---|
| Δ GLX | | Inverse weighted | | Bootstrapped | | | Inverse weighted | | Bootstrapped | | |
| **Measures** | **n** | **r** | ***p* value** | **r** | **95 % CIs** | ***p* value** | **r** | ***p* value** | **r** | **95 % CIs** | ***p* value** |
| Δ cUHDRS | 31 | 0.24 | 0.20 | 0.24 | -0.29, 0.56 | 0.26 | 0.25 | 0.17 | 0.24 | -0.24, 0.55 | 0.20 |
| Δ DBS | 31 | -0.01 | 0.97 | -0.03 | -0.37, 0.31 | 0.87 | 0.02 | 0.91 | 0.00 | -0.35, 0.34 | 1.00 |
| Δ TFC | 31 | 0.26 | 0.16 | 0.28 | -0.12, 0.58 | 0.11 | 0.28 | 0.13 | 0.29 | -0.01, 0.59 | 0.05 |
| Δ TMS | 31 | -0.39 | **0.03** | -0.36 | -0.56, -0.04 | **0.004** | -0.40 | **0.03** | -0.37 | -0.56, -0.03 | **0.002** |
| Δ SDMT | 31 | -0.10 | 0.58 | -0.12 | -0.60, 0.20 | 0.53 | -0.11 | 0.56 | -0.13 | -0.56, 0.19 | 0.48 |
| Δ SCN | 31 | -0.02 | 0.90 | 0.04 | -0.38, 0.47 | 0.86 | -0.03 | 0.87 | 0.03 | -0.38, 0.42 | 0.88 |
| Δ VFC | 31 | 0.14 | 0.45 | 0.08 | -0.37, 0.43 | 0.70 | 0.14 | 0.45 | 0.08 | -0.37, 0.43 | 0.71 |
| Δ SWR | 31 | 0.09 | 0.62 | 0.13 | -0.29, 0.38 | 0.45 | 0.09 | 0.62 | 0.12 | -0.29, 0.39 | 0.45 |
| Δ Whole brain | 30 | 0.05 | 0.78 | 0.03 | -0.31, 0.37 | 0.87 | -0.01 | 0.95 | -0.02 | -0.37, 0.31 | 0.93 |
| Δ Caudate | 31 | 0.27 | 0.14 | 0.31 | -0.05, 0.55 | 0.05 | 0.30 | 0.10 | 0.33 | 0.11, 0.56 | **0.005** |
| Δ White matter | 30 | 0.29 | 0.13 | 0.30 | -0.01, 0.57 | **0.04** | 0.36 | 0.05 | 0.36 | 0.11, 0.56 | **0.002** |
| Δ Grey matter | 30 | 0.39 | **0.04** | 0.43 | 0.13, 0.69 | **0.002** | 0.45 | **0.01** | 0.49 | 0.28, 0.68 | **<0.001** |

*Relationships between rate of change (Δ) in MRS metabolites and Δ in clinical, cognitive and imaging markers were assessed using Pearson's partial correlation controlling for age, and age and CAG repeat length. Correlation coefficients and 95% confidence intervals were computed using bootstrap testing with 1000 repetitions. A weighted correlation was also conducted, applying an inverse weighting to %SD value. Results displayed are unadjusted for multiplicity. Bold text indicated significance $p < 0.05$. cUHDRS, composite Unified Huntington's Disease Rating Scale; DBS, Disease Burden Score; TFC, Total Functional Capacity; TMS, Total Motor Score; SDMT, Symbol Digit Modalities Test; SCN, Stroop Colour Naming; VFC, Verbal Fluency Categorical; SWR, Stroop Word Reading Test; NfL, Neurofilament Light Chain; mHtt, Mutant Huntingtin.*
